# Enhancing patient stratification and interpretability through class-contrastive and feature attribution techniques

**DOI:** 10.1101/2024.03.25.24304824

**Authors:** Sharday Olowu, Neil Lawrence, Soumya Banerjee

## Abstract

A crucial component of the treatment of genetic disorders is identifying and characterising the genes and gene modules that drive disease processes. Recent advances in Next-Generation Sequencing (NGS) improve the prospects for achieving this goal. However, many machine learning techniques are not explainable and fail to account for gene correlations. In this work, we develop a comprehensive set of explainable machine learning techniques to perform patient stratification for inflammatory bowel disease. We focus on Crohn’s disease (CD) and its subtypes: CD with deep ulcer, CD without deep ulcer and IBD-controls. We produce an interpretable probabilistic model over disease subtypes using Gaussian Mixture Modelling. We then apply class-contrastive and feature-attribution techniques to identify potential target genes and modules. We modify the widely used kernelSHAP (Shapley Additive Explanations) algorithm to account for gene correlations. We obtain relevant gene modules for each disease subtype. We develop a class-contrastive technique to visually explain why a particular patient is predicted to have a particular subtype of the disease. We show that our results are relevant to the disease through Gene Ontology enrichment analysis and a review of the literature. We also uncover some novel findings, including currently uncharacterised genes. These approaches maybe beneficial, in personalised medicine, to inform decision-making regarding the diagnosis and treatment of genetic disorders. Our approach is model-agnostic and can potentially be applied to other diseases and domains where explainability and feature correlations are important.

## 1 Introduction

In recent years, vast amounts of genomic data have become publicly available, driven by the development of next-generation sequencing technologies such as RNA-Seq [1]. These allow us to study patterns of gene expression within tissue samples, which can be used to analyse the genetic component of various diseases.

Genetic diseases can be characterised by the activity of certain genes, which often work in concert as modules, driving specific biological processes. The objective of this work is to develop explainable techniques for the identification of genes and gene modules associated with disease. Advanced machine learning techniques have greater ability to capture nuanced relationships between genes and, despite high dimensionality and noise, enable clustering or classification of disease subtype. However, many of these machine learning models are difficult to explain.

An active area of research is in developing methods to explain machine learning model predictions: Explainable AI. This is increasingly important in sensitive domains like healthcare, recruitment and adjudication. For example, in clinical settings, any predictions drawn must be grounded in sound reasoning, given the risks involved.

In this work, we develop a technique for explainability, to identify genes and gene modules associated with disease. We focus on improving the explainability of patient stratification from genomic data. We develop explainability techniques to account for key characteristics of gene expression such as gene distributions, correlations, and dependencies.

Inflammatory Bowel Disease (IBD) can be categorised into Crohn’s Disease (CD) and Ulcerative Colitis. IBD is a chronic digestive disorder characterised by inflammation of the gastrointestinal tract, causing symptoms such as abdominal pain, diarrhoea, and weight loss. There is known to be a strong genetic component, but the risk factors are not fully understood [2].

We use our methods to analyse bulk RNA-Seq data and explore the genetic basis of CD subtypes. We use our techniques to analyse transcriptomic profiles and identify genes associated with subtypes of CD. Patient stratification can enable targeted treatment. Identifying therapeutic gene targets may inform the development of more effective treatments.

### 1.1 Overview of approach

Figure 1 gives an overview of our approach. Our contributions are summarised below:

- We predict Crohn’s disease (CD) subtype based on gene expression data. We use a machine learning model called a Gaussian Mixture Model which performs soft clustering. The disease subtypes predicted are Crohn’s disease with deep ulcer (a severe form of the disease), Crohn’s disease with no ulcer, and IBD-control.
- We adapt an explainable AI technique called kernelSHAP to account for gene correlations. We couple kernelSHAP to a probabilistic model we derive from the Gaussian Mixture Model, to quantify the influence of genes for each Crohn’s disease subtype.
- We then use consensus clustering [3] to identify potential gene modules by Crohn’s disease subtype.
- For these gene modules, we determine the type and relative magnitude of influence on disease subtype. These are confirmed using Gene Ontology enrichment analysis.
- Finally, we develop a class-contrastive technique to visually explain the impact of gene modules on the disease subtype for an individual patient.

**Figure 1:**
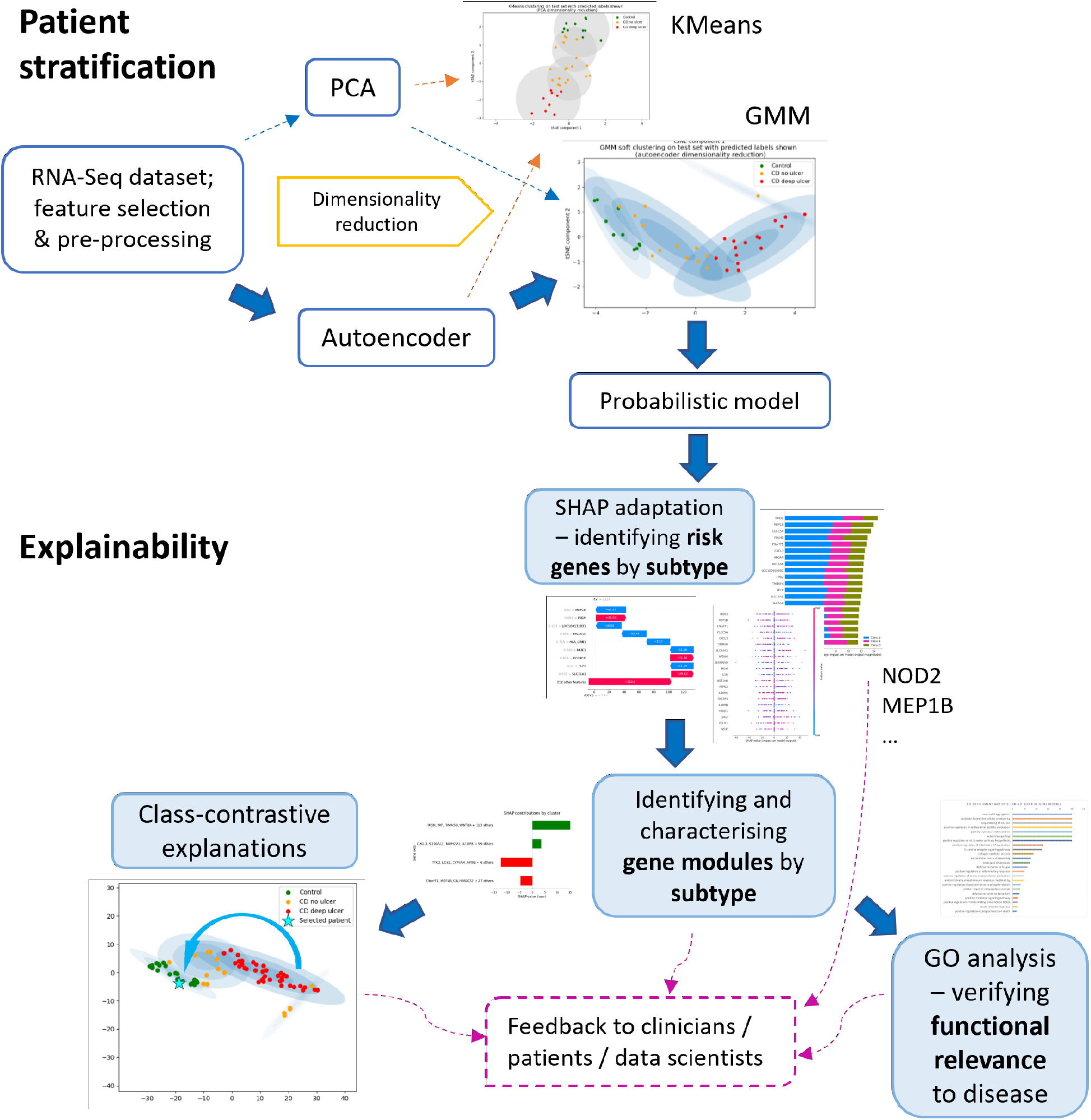
Overview of our computational framework. We identify genes and gene modules implicated in Crohn’s disease (CD) subtypes. Starting with RNA-Seq data, we perform feature selection and reduce dimensionality using PCA and an autoencoder. We then use a probabilistic model (Gaussian Mixture Model [GMM]) to cluster patients into disease subtypes (CD with deep ulcer, CD without deep ulcer and control). In order to explain our model we develop: 1. an extension of Shapley Additive Explanations (SHAP) to account for gene correlations, and 2. a class-contrastive technique to visually demonstrate the effect of changing gene expression on individual patients. We confirm these findings by referencing a variety of peer-reviewed studies and conducting a Gene Ontology (GO) enrichment analysis.

We identify known IBD risk genes such as NOD2, IRGM, JAK2 and IL10. Furthermore, our analysis suggests a role for uncharacterised genes such as LOC100505851 and LOC100132831. We show using Gene Ontology enrichment analysis that the identified gene modules are relevant to IBD. We visually demonstrate the impact of these genes on each patient using a class-contrastive technique.

Our techniques may aid in the diagnosis and treatment of genetic diseases. Our approach may also be broadly applicable in other domains where explainability and feature correlations are important.

## 2 Background

We highlight key theoretical background to our work here; please see the Supplementary Material for a comprehensive review of all techniques.

### 2.1 Gaussian Mixture Models (GMMs)

Gaussian Mixture Models (GMMs) can be used to perform soft clustering of data. They help organise data into groups, which is useful for finding disease subtypes based on our transcriptomic dataset.

GMMs work by using a mix of bell-shaped curves (Gaussian distributions) to represent the groups. These groups are called mixture components and have associated parameters such as mean *μ*_*k*_ and covariance Σ_*k*_. The mix of these groups and how important they are in the model is determined by mixing coefficients (*π*_*k*_). These coefficients show how much each group contributes to understanding the overall data.

To make these groups fit the data best, we adjust them iteratively using a process called maximum-likelihood estimation. This process fine-tunes the groups’ characteristics to match the observed data distribution.

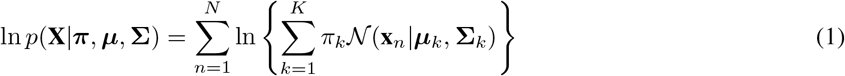

This equation helps us evaluate how well our groups fit our observed data. We aim to adjust our groups to maximise this measurement, ensuring our model accurately represents the data.

### 2.2 SHAPley Additive exPlanations (SHAP)

SHAPley Additive exPlanations (SHAP) [4] is a method to explain the predictions of machine learning models. It aims to find the contribution of each feature to the model output by using a game-theoretic approach.

KernelSHAP provides an efficient approximation to SHAP values using weighted linear regression based on sampling. Rather than retraining a new model for each coalition as with classic SHAP, we marginalise the missing features out of the model. In Eq 2, we define a fidelity function *L* that measures how unfaithful is a surrogate model *g* in approximating the model *f*, in the feature subspace defined by *z*^*′*^, across all models. Here, we use *z*^*′*^ ∈ {0, 1}^*M*^ to define the coalition of features, where *M* is the number of input features.

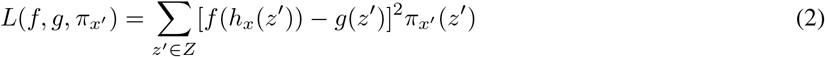

We generate synthetic samples for each model, where each baseline sample *z* is drawn from the same probability distribution as the input features. We can compute the model output *f*(*h*_*x*_(*z*′)) as 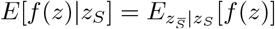. However, kernelSHAP assumes feature independence so 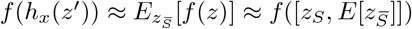. This means we simulate the subset of missing features 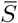 using expectation values, to show that these features carry no information. We use *z*^*′*^ to represent a perturbed version of the sample *z*, where the included features *S* take their value from the input instance we are analysing. The *h*_*x*_ function is used to map the samples to a potentially higher-dimensional space. A kernel weighting function is also used to emphasise the independent and global effects of features.

We then perform linear regression to minimise the fidelity function *L*, which gives rise to Eq 3. *ϕ*_0_ is the expected model output when no features are present and the remaining coefficients *ϕ*_1*−M*_ are the SHAP values of the corresponding features.

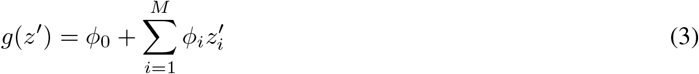

We aim to find the SHAP value of each feature, as this represents the type and extent of influence that a feature has on the model prediction. This explainability technique can be applied to estimate the influence of particular genes on patient outcomes. Further algorithmic details are available in the Supplementary Material.

## 3 Related work

### 3.1 Explainability

#### 3.1.1 Class-contrastive techniques

Class-contrastive techniques can be used to find the impact of particular features on the model output. This can be very useful for improving the transparency of a model. For example, Banerjee et al. generate explainability for mortality predictions of patients in [5], as predicted by deep learning models. By setting the presence or absence of binary features such as “suffering with depression” or “lack of family support”, they were able to find the isolated effect of each feature on the risk of patient mortality (as predicted by a black-box model). This technique is usually used with categorical features. The class-contrastive approach has been used for self-supervised clustering of RNA-Seq data [6], but not for generating explanations of disease subtype based on genomic data. In this work, we extend the class-contrastive approach to demonstrate the impact of gene modules on disease subtype, taking into account the underlying gene expression distributions.

#### 3.1.2 Applications of SHAP

SHapley Additive exPlanations (SHAP) [4] is another state-of-the-art explainability technique. Fast approximations of SHAP have been applied to analyse gene expression data [7, 8, 9, 10, 11], such as kernelExplainer, treeExplainer and gradientExplainer [12]. Yu et al. use a deep autoencoder [9] to learn gene expression representations, applying treeExplainer SHAP to measure the contributions of genes to each of the latent variables.

Although SHAP has shown success in this line of work, one significant problem is that it assumes feature independence. This means that when applying SHAP to find the contribution of genes, it is assumed that there are no correlations between genes. This is unrealistic because genes are often correlated and/or regulated by other genes; this is governed by complex gene regulatory networks [13].

There have been some attempts to include feature dependence, for example in the linearExplainer and treeExplainer [14] SHAP variants. However, linear models are not suitable for modelling complex gene-gene and genotype-phenotype relationships. The treeExplainer is also limited to tree-based ensemble methods, which can be difficult to visualise and interpret. We aim to address this by incorporating inter-feature dependence for kernelSHAP (kernelExplainer), which is model-agnostic and therefore applicable in many more contexts. By incorporating inter-feature dependence, we can more accurately identify potential genes, gene modules, and associated biological pathways implicated in disease processes.

Aas et al. propose to achieve inter-feature dependence for kernelSHAP in [15]. Here, a multivariate Gaussian distribution is constructed using a sample mean vector and covariance matrix, calculated from the training data. For each input instance (corresponding to a coalition of features), this model is updated under a Bayesian framework and used to generate synthetic samples for the calculation of conditional expectations required by the kernelSHAP algorithm (Section 2b). In the context of RNA-Seq analysis, this method is not appropriate because the model of relationships between genes can differ significantly between input instances. More specifically, the Bayesian framework leads to the modification of feature correlations and the means of marginalised features to varying degrees.

In this work, we use an alternative approach to address the need to take account of consistent relationships between features. We construct a multivariate Gaussian distribution to model these relationships. However, when updating this model between input instances, we only modify the mean and variance of those features present in the current model’s coalition, leaving feature correlations intact. In this way, we preserve our knowledge of the underlying dataset, including the relationships between genes originally captured in the training data. This results in a more representative multivariate Gaussian distribution. Since we are aiming to draw insights about consistent relationships between genes, this promotes more realistic SHAP values and therefore cluster explanations.

### 3.2 Cluster analysis and gene module identification

Identifying gene modules is a crucial step in characterising the genetic component of disease. Current techniques tend to include a clustering aspect and/or network construction [16, 17, 18] to organise genes, such as Weighted Gene Co-expression Network Analysis [19, 20]. However, they can be sensitive to noise, with high computational complexity that can limit scalability to larger datasets. Our approach also uses clustering, but reduces complexity and the impact of noise by implicitly capturing gene and sample relationships. We achieve this by using Gaussian Mixture Modelling and a deep autoencoder that can infer both linear and non-linear relationships. We adapt our mixture-based clustering model for classifying disease subtype based on RNA-Seq data.

Our approach explicitly accounts for inter-feature dependence by analysing the underlying data distributions and correlations between genes, using data from real patients.

## 4 Data and Methods

Our work is comprised of two main stages. The first stage involved using dimensionality reduction and clustering techniques for patient stratification. Our data included patients with Crohn’s disease (CD) and controls. The subjects were classified into subtypes within the dataset: CD with deep ulcer (the most severe form of the disease), CD without deep ulcer and controls. The goal of patient stratification was to discover these subtype groups.

The second stage involved adapting state-of-the-art methods for explainability, and identifying and characterising genes and gene modules associated with each disease subtype.

### Patient stratification

1. Sampling from a publicly available genomic dataset
2. Dimensionality reduction using an autoencoder and PCA
3. Clustering and classification with GMMs and Kmeans

### Explainability

4. Adapting kernelSHAP to identify genes that are involved in disease
5. Identification of potential gene modules by disease subtype
6. Class-contrastive technique for patient-specific explainability

### 4.1 Patient stratification

#### 4.1.1 Sampling from a public RNA-Seq dataset

We performed our analyses on a publicly available transcriptomic dataset called RISK [21]. This contains RNA-Seq data from ileal tissue samples. The samples were taken from children: non-IBD controls as well as patients diagnosed with IBD who had not yet undergone treatment. The RNA was sequenced as described in [21], resulting in normalised counts in RPKM (Reads Per Kilobase of transcript per Million mapped reads). We removed data on Ulcerative Colitis to focus on Crohn’s disease (CD) subtypes. We used data from 260 individuals who were classified into three groups: CD with deep ulcer, CD without deep ulcer and non-IBD controls. Non-IBD controls were those with “suspected IBD, but with no microscopic or macroscopic inflammation and normal radiographic, endoscopic, and histologic findings”. The goal of patient stratification was to develop a robust method for recovering these subtypes.

The first task was to sample a relevant selection of genes from the RISK dataset [21, 22]. Many genes in the dataset did not vary much in expression across the sample of patients. We decided to analyse genes with a variance of at least 0.01 (normalised) RPKM across the sample, to help us identify those that could reasonably affect disease subtype.

In addition to the RISK dataset, a supplementary dataset from [21, 22] contained 1,281 genes that were found to be differentially expressed with a fold change of at least 1.5, between two independent CD and control groups. For each independent group, we identified the top 60 most upregulated genes and top 60 most downregulated genes. This resulted in 240 genes, from which we identified 130 matches in the RISK dataset. Further exploration of the literature identified more genes associated with IBD, such as JAK2, NOD2 and LTA. 41 of these genes were discovered in the RISK dataset and added to the sample. To promote more diversity, we also added a random sample of 50 genes from the RISK dataset not linked to IBD. This was done to provide potential models with a broader range of gene types that could be used to differentiate between disease subtypes. However, this also requires a model to be robust to random noise. Our selection process resulted in a total of 221 genes.

#### 4.1.2 Dimensionality reduction approach

An autoencoder model (adapted from [23]) was used to reduce dimensionality, using the given sample of 221 genes (and their associated expression values) in the RISK dataset. The dataset was randomly shuffled and split 70:30 into training and test sets. We then performed feature scaling [24]. The model was implemented using the Keras Functional API [25]. We experimented with different layers, layer sizes and activation functions to achieve a good performance. The architecture for the encoder and decoder sections is detailed in Table 1 of the Supplementary Material. 32 neurons were used in the bottleneck layer to produce a 32-dimensional latent representation.

**Table 1:**
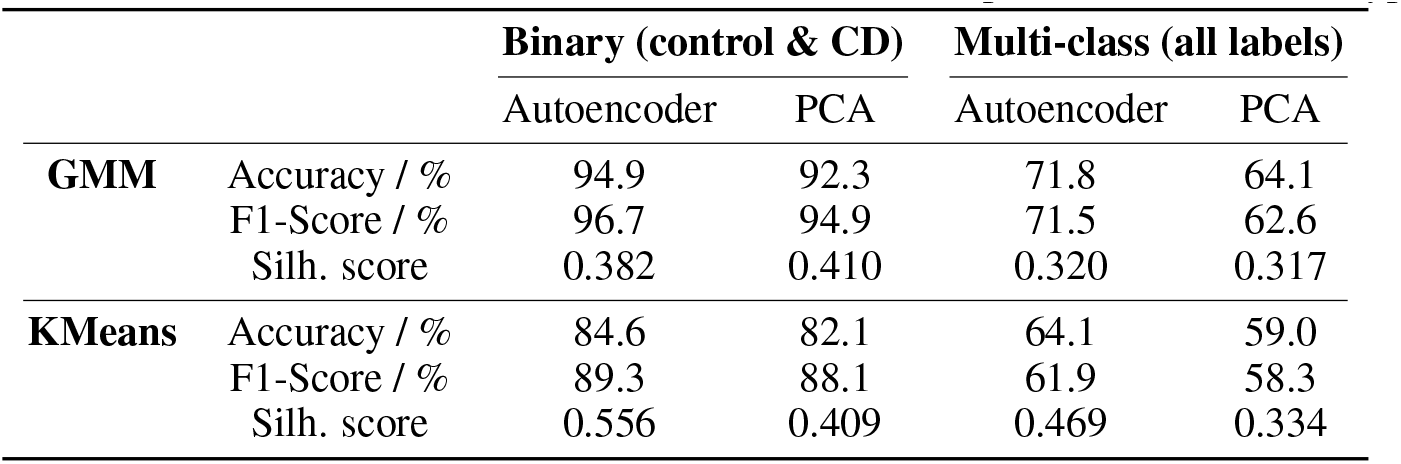
Clustering and classification evaluation results for final Gaussian Mixture Model (GMM) and KMeans models, using autoencoder and PCA dimensionality reduction methods. Results shown for binary classification (controls and all CD patients) and multi-class classification (control, CD no ulcer and CD deep ulcer) of disease subtype.

To tune the model, we used the Keras ‘GridSearchCV’ function for a 5-fold cross-validated grid search over the hyperparameters: epochs, batch size, optimiser, learning rate and weight initialisation function. The final model had the following hyperparameters: batch size of 32, 150 epochs, Adam optimiser, 0.001 learning rate and Xavier normal initialisation function. We used 80% of the training data for this training, keeping the remaining 20% for validation. Mean Squared Error (MSE) was used as a performance metric.

In this work, we applied both PCA and an autoencoder for dimensionality reduction. In both cases, after reducing to 32 latent variables, t-SNE (t-distributed Stochastic Neighbour Embedding) was used for visualisation.

#### 4.1.3 Gaussian Mixture Modelling (GMM) and KMeans clustering

We explored both GMM and KMeans clustering algorithms for patient stratification. We first divided the gene expression dataset into training, validation and test sets using a 70/15/15 split. After reducing dimensionality using the PCA-tSNE and autoencoder-tSNE methods explained previously, the GMM and KMeans algorithms were trained on the training set. We used 4 clusters over 3 disease subtype classes to allow for the discovery of more potential subtypes. Each component in the GMM was set to have an individual covariance matrix, to fit the model closely to the data. As the perplexity used for t-SNE can have a large impact on performance, we tuned this hyperparameter on the validation set, using accuracy, F1-score and silhouette analysis (explained in Supplementary Material Section 1.4.5).

For classification, we designed a post-processing algorithm to assign each of the four clusters to a disease subtype (CD with deep ulcer, CD with no ulcer, or control). This is summarised in Algorithm 1. In essence, each cluster was assigned to the class with the greatest density of its datapoints present. For GMMs, the mixture component probability density functions were used in this estimation. This was replaced with a simple datapoint count for KMeans. We then managed possible duplicate assignments in *handle*_*duplicates*(). More specifically, where two classes were initially assigned to the same cluster, the class with the greatest density present took precedence. The remaining class was then assigned to one of the remaining clusters with the greatest density of its points present.

The next step was to explore methods for the classification of disease subtype. Realising the power of GMMs, we implemented a function to generate a probability distribution across the disease subtype classes for any new data sample, given our trained clustering model and class assignments generated by Algorithm 1. We first evaluated the probability density of each mixture component with respect to the given point. For the class with two clusters assigned, we took the highest probability density of the two components, discounting the other component. For each datapoint, we normalised the resulting values by dividing each by the total sum, to ensure that the probabilities added up to one. Therefore, the probability of a datapoint *x* being classified as *a*, the class of interest, would be 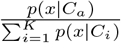 where *C* represents a disease subtype class and *K* = 3, the number of disease subtypes. Therefore, we adapted the GMM into a probabilistic model for the classification of CD subtype. KMeans subtype predictions for the test set were based on the closest cluster center and its given subtype assignment, since no probability density function was available. The models were visualised [26] and evaluated on the test set using accuracy, F1-score and silhouette analysis [27, 28]. This allowed us to assess clustering quality and classification ability.

### 4.2 Clustering explainability

#### 4.2.1 Modifying kernelSHAP to identify risk genes

As explained in Section 2(b), SHAPley Additive exPlanations (SHAP) is a state-of-the-art method for machine learning model explainability. KernelSHAP is a model-agnostic fast approximation of SHAP. This is commonly applied to regression or classification models. In this work, we developed a post-processing technique for Gaussian Mixture Models (GMMs). We produced a probability distribution over the disease subtypes for each patient. Therefore, we could couple our mixture models to kernelSHAP for patient-specific and global cluster explainability.

However, a major limitation of kernelSHAP in this context is that it assumes feature independence i.e. gene independence. In reality, gene expression in biological systems can be highly correlated between genes, and many genes are regulated by other genes within a complex network. Therefore, we extended kernelSHAP to incorporate inter-feature dependence, to enable more accurate cluster explanations.

The original kernelSHAP implementation is explained in Section 2b and Supp. Material 1.5.2 [4]. During the calculation of SHAP values, we perform linear regression which involves calculating model output expectations for each coalition of features. If we denote *S* as the subset of features being included in the coalition for a model *f*, and *S* as the set of missing features, the expectation *E* for the model output can be calculated as follows:

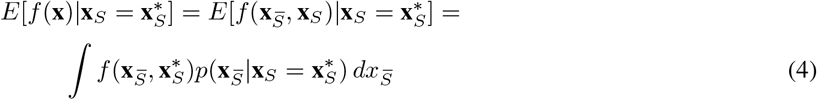

as explained in [29], where 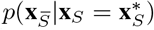 is the conditional distribution over the missing feature values, given a set of known values **x**_*S*_ for the subset of features *S* included in the given coalition, obtained from the current input of interest **x\*** (see Section 2b for full algorithmic details). To simplify the process, the original kernelSHAP implementation assumes feature independence and instead draws from the marginal distribution [4]. Building upon work by Aas et al. in [29], we propose an adaptation to approximate the conditional distribution, resulting in more representative synthetic samples and therefore more realistic SHAP values.

We can approximate the conditional distribution by modelling the underlying data distributions of the training set using a multivariate Gaussian distribution. We calculate a mean vector and sample covariance across the training data to construct this distribution. For each new input instance, we modify the distribution according to the features present in the coalition, before drawing synthetic samples for the expectation calculation.

Because we aim to uncover the relationships between genes, we preserve the relationships captured by the sample covariance and expectation on the training data. We reduce the variance of a particular feature to zero if it is included in the coalition. The corresponding values in the mean vector are also updated to be equal to values given in the input instance. In this way, we maintain the important underlying distributions and correlations between features (genes).

We then adjust the matrix to make it positive definite by adding a small multiple of the identity matrix: *C* = *C* + *ϵI*_*p*_, where *I* is a *p* x *p* identity matrix and *ϵ* = *abs*(*λ*_*min*_) + *b*, where *λ*_*min*_ is the smallest eigenvalue of *C* and *b* = 1.5. This ensures that we obtain a valid probability density function for sampling. The constant *b* can be tuned to affect *ϵ*; a larger *ϵ* value will result in greater overall strength of covariance between features. However, a value too large may distort the distribution. From the resulting distribution, we generate samples which are used to evaluate the expectations in Eq. 4. We must rescale each sample to account for the variances modified when making the covariance matrix positive definite, by applying the transformation *x* = ***σ***(*x −* ***μ***) + ***μ***. We also clip the values between 0 and 1 as the data is in normalised form.

Taking gene relationships into account results in synthetic samples with realistic background expression values for marginalised genes. We can therefore obtain more realistic SHAP values to explain the phenotypes predicted by the Gaussian Mixture Model. SHAP values are calculated on the test set, following the calculation of expected values using the training set. These calculations use the probabilistic model over Crohn’s disease subtypes, derived from the Gaussian Mixture Model. We then generate individualised patient-specific plots and summary plots to explain the clusters.

#### 4.2.2 Identification and characterisation of potential gene modules

We propose a method for identifying potential gene modules that explain disease subtype. The method relies on our kernelSHAP feature dependence adaptation and involves the following processes:

- Integration of SHAP values and gene expression data
- Consensus clustering
- Characterisation of gene modules
- Verification of gene modules using Gene Ontology enrichment analysis

Firstly, we prepared the data by combining different sources of information in a useful way. For each gene, we found how much it affects the model’s prediction for a specific disease subtype. This was done by averaging how important the gene is across all patients with that disease subtype. Then, we multiplied this average importance by the gene’s activity level across all patients.

The process is shown in Equation 5. This equation calculates representative values for each patient and gene in a specific disease type. Here, *x* represents a gene’s activity level, and *s* represents its importance according to the model (SHAP value). We add up the importance values (using *c*_*i*_ to identify patients in a particular disease subtype) and divide by the total number of patients (*n*) to find the average importance for each gene.

The resulting dataset shows how active each gene is and how much it influences the model’s prediction. This prediction represents how confident we are about assigning a particular disease subtype to a patient.

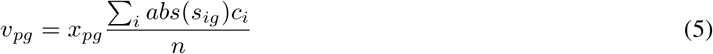

We then performed consensus clustering on the integrated data, using Weighted Ensemble Consensus of Random (WECR) K-Means, proposed by Yongxuan et al. in [3]. To summarise the WECR algorithm, we first run the K-means algorithm several times on random samples of the data using random subspaces of features, to generate an ensemble of base partitions. The value of k is also randomised for each run. In this work, we run K-Means 100 times, where each single run draws 80% of the sample and 50% of the features. To obtain the final clustering, we evaluate each base partition and form a co-association matrix. We then apply cluster-based similarity partitioning and spectral clustering [30]. Please see [3] for more details about the WECR algorithm.

As the number of gene modules in the data is unknown and dimensionality is high, applying this consensus clustering is suitable to obtain more stable clusters. For the same reason, we integrated different types of data and used four different validation metrics to select the optimal number of clusters *k* from 2 to 9. These were the Bayesian Information Criterion (BIC), Davies-Bouldin (DB) Index, Silhouette Score (SIL) and Calinski-Harabasz (CH) Index, where BIC and DB should be minimised, and SIL and CH should be maximised. In this way, we obtained representative modules informed by similarities in expression pattern and influence on disease subtype. We then displayed aggregate bar plots to show the relative contributions made by each gene module in predicting a particular disease subtype. This is represented using a sum of the mean SHAP values across all genes in the module (where the mean is calculated across all patients of the given disease subtype).

Incorporating inter-feature dependence provides valuable insights into sets of genes which could be working in concert: this includes the type and magnitude of their influence on the model’s prediction of disease subtype.

Finally, we confirm the functional relevance of the gene modules using Gene Ontology enrichment analysis.

#### 4.2.3 Class-contrastive technique for patient-specific explainability

In this work, we develop a class-contrastive technique for explaining clusters specific to a patient. Class-contrastive reasoning generates an explanation by providing a contrast to another class. An example of a class-contrastive explanation is: “The selected patient is predicted to be in a severe disease subtype (CD with deep ulcer) because all genes in a particular module were overexpressed. If these genes had abundance similar to genes in the control group, then the patient would be predicted to be in a less severe disease category (CD without deep ulcer).”

The expression of each gene approximately follows a normal distribution. Some genes were upregulated or downregulated in patients with CD (Crohn’s disease) compared to controls. Therefore, for a specific patient with CD, we can generate a class-contrastive explanation in the following way: we can modify the expression of genes in a given module so that they are more similar to controls. We did this by assigning a new expression value *v* for each chosen gene, as the mean value for the expression of this gene across all control individuals, as shown in Eq 6.

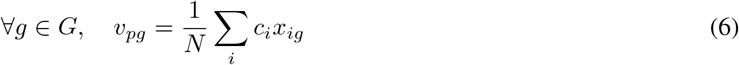

where *x* is an expression value, *p* is the selected patient, *g* is the selected gene and *G* is the full set of genes in the module. We sum over all control individuals *i* in finding the mean, where *c*_*i*_ is the indicator variable for the control group and *N* is the total number of control individuals.

We can reduce the dimensionality, as described in Section 4(a)(iii), and generate the GMM clustering for the dataset before and after this correction. If the patient with CD moves into a different CD cluster after correction, we can infer how the genes in the module may be affecting the disease. In Section 5(e) we adapt this technique for intuitive patient-specific explainability, showing how gene modules can contribute to CD subtype. As the identification of gene modules relies on the feature dependence extension proposed in Section 4(b)(i), this also takes gene correlations into account across all patients. This method combines the strengths of feature attribution, consensus clustering and class-contrastive reasoning.

### 4.3 Software

Data manipulation, machine learning and visualisation were implemented in Python using the following libraries: Numpy [31], Pandas [32], SciPy [33], Seaborn [34], Scikit-learn [27], Tensorflow [35], Keras [25], Matplotlib [36], Shap [4] and Pyckmeans [3].

All code is available from the following repository: https://zenodo.org/doi/10.5281/zenodo.10278383

## 5 Results and Discussion

In this section, we evaluate and discuss the significance of our results. Please see the Supplementary Material (Section 3) for further results and technical details.

### 5.1 Gaussian Mixture Model (GMM) and KMeans clustering

KMeans and Gaussian Mixture Models were implemented to cluster the samples into 4 groups. We then added a post-processing step to transform the models into classifiers of disease phenotype for Crohn’s disease (CD). The three classes/disease subtypes were “CD with deep ulcer”, “CD no ulcer” and “control”. We used both PCA and an autoencoder for dimensionality reduction of the data, each alongside tSNE for visualisation in two dimensions. We then apply the clustering techniques. The steps are as follows:

1. PCA → tSNE → KMeans
2. Autoencoder → tSNE → KMeans
3. PCA → tSNE → GMM
4. Autoencoder → tSNE → GMM

Our autoencoder model shows good performance, achieving a MSE of 0.0143 on the test set, despite its simplicity compared to the state-of-the-art [37, 38]. This suggests good capabilities in reducing dimensionality of the RNA-Seq data, while retaining important information.

The final results for GMM clustering on the test set are shown in Figure 2, using dimensionality reduction by the autoencoder (left) and PCA (right). The final evaluation results are summarised in Table 1. Please see the Supplementary Material for KMeans visualisations and results from the training process.

**Figure 2:**
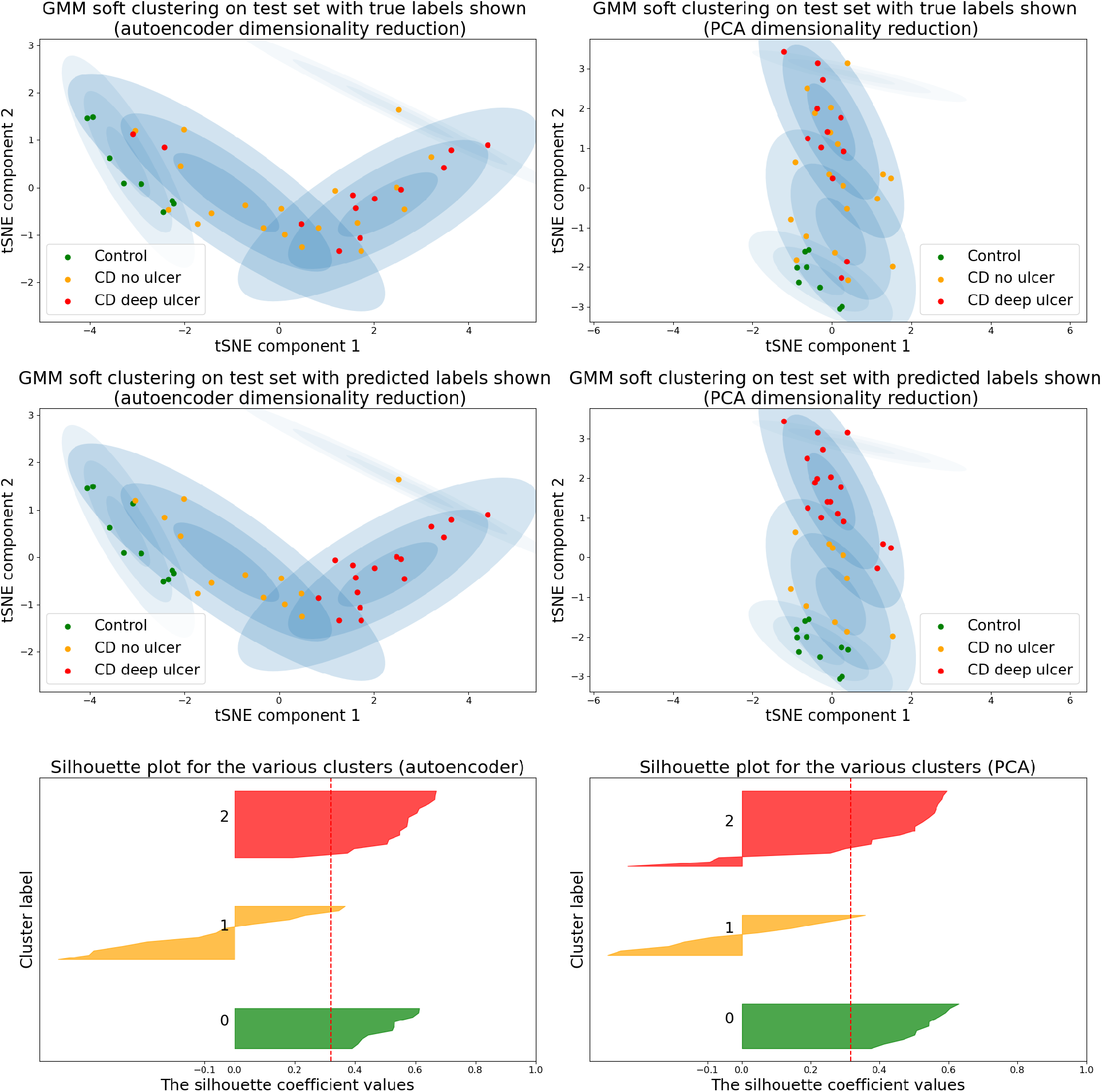
Gaussian Mixture Model (GMM) clustering model results after applying dimensionality reduction using autoencoder and tSNE (perplexity=130) (left) and PCA and tSNE (perplexity=150) (right). Deployed on the test set with true labels shown (top third) and predicted labels shown (middle third). Silhouette plots are shown for GMM clusters after applying autoencoder-tSNE (left) and PCA-tSNE (right) methods, with clusters 0, 1 and 2 corresponding to “control”, “CD no ulcer” and “CD deep ulcer” respectively. These are the disease subtypes and CD deep ulcer is the most severe form of the disease.

The results show a good overall performance (Table 1). Clustering provides an informative visual representation of relationships between patients in terms of disease subtype. In particular, the GMM provides an effective density estimation which is useful for inferring an accurate disease subtype during post-processing. Our autoencoder also performs better than PCA, particularly in the context of multi-class classification of disease subtype. Here, accuracy and F1-score were higher by 7.7% and 8.9% respectively when using our autoencoder compared to PCA (Table 1) [when using GMMs]. In addition, by using a greater number of clusters than disease subtypes, we may discover additional substructures which could correspond to potentially new subtypes of CD. Please see Sections 3.5 and 3.6 of the Supplementary Material for a detailed evaluation and comparison of the clustering methods and dimensionality reduction techniques.

### 5.2 Cluster explanation using kernelSHAP adapted for feature dependence

As explained in Section 4(b)(i), we can couple our GMMs to kernelSHAP [4] to generate explainability for each cluster, including visualisations [39]. As each feature corresponds to a gene and each cluster class corresponds to a disease subtype, the resulting SHAP values represent the importance of each gene in predicting a particular disease subtype for a given patient. Additionally, we modify the original kernelSHAP method to incorporate feature dependence. This more accurately models the living system, as many genes are highly correlated and/or regulated by other genes. Using this method, we can therefore identify the genes that are the most influential in predicting given disease subtypes. Please see the Supplementary Material for additional results, such as force plots and beeswarm plots.

#### 5.2.1 Waterfall plot

A waterfall plot can be used to analyse a single patient, as shown in Figure 3. This explains the model output for the CD deep ulcer cluster class. It shows a quantification of the contributions made from the top genes identified, alongside that of the remaining genes. In this way we can see how the model output has been shifted from the expected value *E*[*f* (*z*)], to the actual output, *f* (*x*). In the former, the model *f* is provided with a baseline sample *z* and no information about the features, whereas in the latter we provide our actual data sample *x* as input to the model. These values are given in the log-odds space.

**Figure 3:**
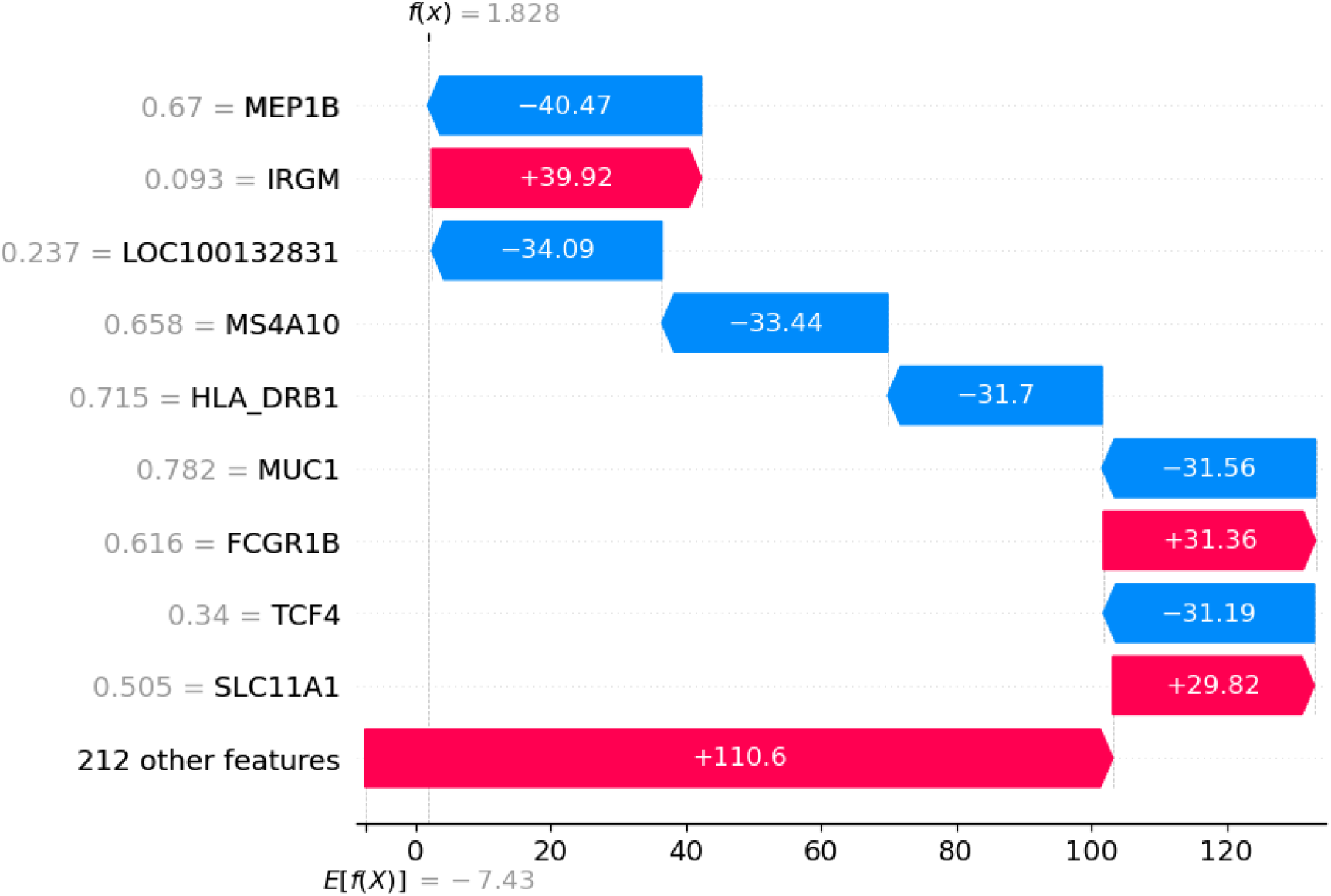
Waterfall plot for analysing model output for a single patient (Patient 260) for the “CD deep ulcer” disease subtype (using dependent features). Shown are contributions made from the top genes identified, alongside that of the remaining genes. The plot shows how the model output has hifted from the expected value *E*[*f* (*z*)], where the model has no information about the features, to the actual output, *f* (*x*) (values are in log-odds space). Shown are some highly relevant genes, such as IRGM which is a negative regulator of IL1B as it suppresses NLRP3 inflammasome activation. It has a protective effect against gut inflammation in CD. IRGM has a relatively low normalised expression level of 0.093 for this patient, which would have little protective effect. This leads the model to predict a greater probability of CD with deep ulcer.

The genes shown are highly relevant. For example, IRGM is a negative regulator of IL1B as it suppresses NLRP3 inflammasome activation by hindering its assembly. In this way, it has a protective effect against inflammatory cell death and gut inflammation in Crohn’s disease [40]. In the plot we can see that IRGM has a relatively low normalised expression level of 0.093 for this patient, which would have little protective effect. This rightfully leads the model to predict a greater probability of CD with deep ulcer. In comparison to the plot resulting from feature independence (Supp. Material Fig. 20), we obtain more genes specifically related to IBD for the same patient (who is diagnosed with CD with a deep ulcer). For example, in addition to IRGM, we also obtain HLA_DRB1, MEP1B, MUC1 and SLC11A1, which all have established links to IBD [41, 42, 43, 44].

#### 5.2.2 Summary plot

Although SHAP values are specific to a data instance, they can be combined to provide global explanations. Figure 4 shows a summary graph of the 20 genes that were the most influential in the overall predictions of the model. The blue, pink and green bars depict the magnitude of influence of a gene on the “CD deep ulcer”, “CD no ulcer” and “control” classes respectively.

**Figure 4:**
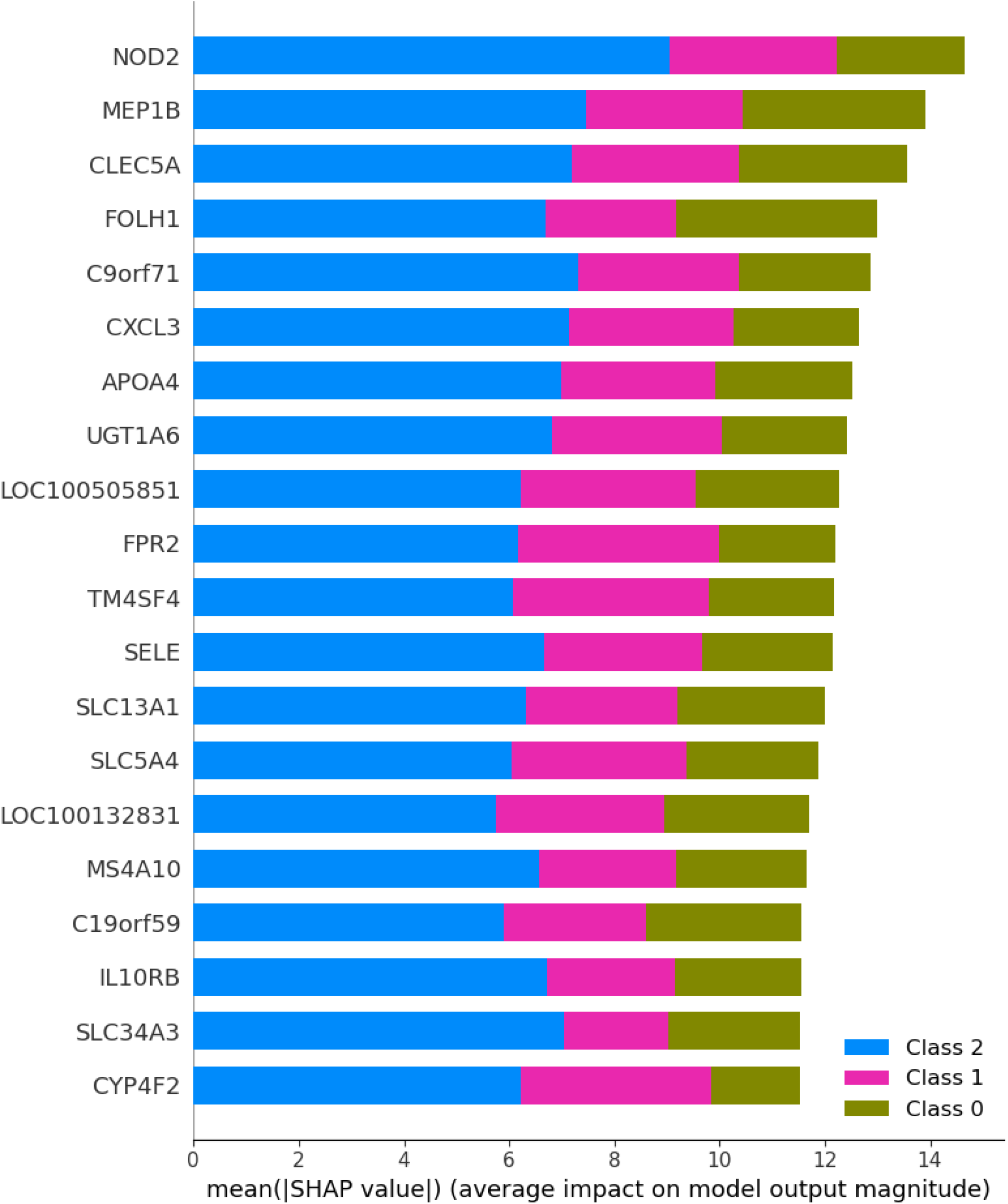
Summary plot showing top 20 genes in terms of their average impact on class predictions across all patients. This is for a model which accounts for feature dependence. The blue, pink and green bars depict the magnitude of influence of a gene on the “CD deep ulcer”, “CD no ulcer” and “control” classes respectively. The greatest proportion of influence of genes is attributed to the “CD deep ulcer” cluster. Genes like NOD2, MEP1B and FOLH1 are within the top 5 and known as susceptibility genes for IBD. The most significant gene identified overall is NOD2, which is known to be strongly associated with IBD.

We can see that each gene contributes to the predictions of each class/disease subtype to varying degrees. For all genes shown, the greatest proportion of their influence is attributed to the “CD deep ulcer” cluster, which is indicative of their significance. Compared to the plot generated without feature dependence (Supp. Material Fig. 21), the genes acknowledged as most influential in the literature are ranked near the top of the list here. For example, NOD2, MEP1B and FOLH1 are within the top 5 and are known susceptibility genes for IBD. By contrast, the top 5 genes generated without feature dependence (Supp. Material Fig. 21) have more tentative links to IBD and C19orf59 has no links. This suggests that the attribution of feature importance may be more accurate when feature dependence is incorporated.

Our analysis suggests that NOD2 is an important gene. This is a “nucleotide-binding oligomerisation domain” responsible for sensing bacteria and is known to be associated with IBD. It does this by recognising a bioactive fragment of peptidoglycan on the bacterial cell envelope, called muramyl dipeptide (MDP). After binding to MDP, NOD2 oligomerises and binds to a serine or threonine kinase RICK. RICK then oligomerises [45], activating the NF-*κ*B signalling pathway. This results in the accumulation of pro-inflammatory cytokines [46], causing inflammation and tissue damage [47].

In addition to NOD2, MEP1B and FOLH1, we obtain other important genes which were not detected using feature independence, such as IL10RB, CXCL3, APOA4, SLC13A1 and SLC5A4: these are all implicated in IBD. Chemokines such as CXCL3 and cytokines such as IL10 are associated with inflammatory processes in IBD [48]. In both summary plots (Fig. 4 and Supp. Material Fig. 21), we obtain LOC100505851 and LOC100132831. These “LOC” genes are currently considered uncharacterised [49] and may represent novel findings in IBD.

Our method is not without limitations. For example, our method found genes, such as SELE, that do not have strong links to IBD.

The ability to generate local explanations, for example, using waterfall plots, may be useful for applications in personalised medicine. Compared to state-of-the-art work, which tends to apply SHAP to black-box neural networks [7, 8, 9, 10], we apply SHAP using a probabilistic model derived from a GMM that captures disease subtype relationships. This further improves the interpretability of SHAP values.

We have demonstrated the ability to generate global explanations by combining SHAP values: this is useful for finding common themes across the data. Most of the top genes identified have well-established links to IBD in the literature. This alignment is more pronounced when feature dependence is incorporated. Other genes may represent novel findings, particularly the uncharacterised “LOC” genes. The next section builds on this work to detect potential gene modules.

### 5.3 Identification and characterisation of potential gene modules

In Section 4(b)(ii), we proposed a novel method to identify potential gene modules. This integrates SHAP values with gene expression values, before performing Weighted Ensemble Consensus of Random consensus clustering [3]. We utilise SHAP values calculated using our kernelSHAP adaptation. This incorporates dependence between genes. We then use the SHAP values to characterise each gene module, verifying our findings with Gene Ontology enrichment analysis.

We first applied our technique to identify gene modules associated with the most severe form of the disease: Crohn’s disease with a deep ulcer. This was achieved by integrating the CD deep ulcer SHAP values with the expression values across all patients. We then perform Weighted Ensemble Consensus of Random (WECR) KMeans clustering. In the supplementary material we show the results of the final clustering on the co-association matrix using various numbers of clusters *k*. The evaluation results are visualised in Supplementary Material. We select *k* = 4 as it achieves a good balance in terms of the validation scores. Using a variety of metrics in the selection of *k* helps us achieve more stable clusters.

Figure 5 shows a bar plot in which each final gene module is characterised in terms of its influence on model predictions. Each bar represents the sum of mean SHAP values associated with the “CD deep ulcer” cluster, across all genes in the module. We show the top 4 most influential genes of each module. More positive values (green) indicate that the given module increases our confidence in a “CD deep ulcer” prediction and more negative values (red) reduce our confidence in a “CD deep ulcer” prediction. “CD no ulcer” gene modules were also identified, following the same process with CD no ulcer SHAP values. The evaluation results and final bar plot can be found in the Supplementary Material. Full gene module memberships for both classes are also given in the Supplementary Material.

**Figure 5:**
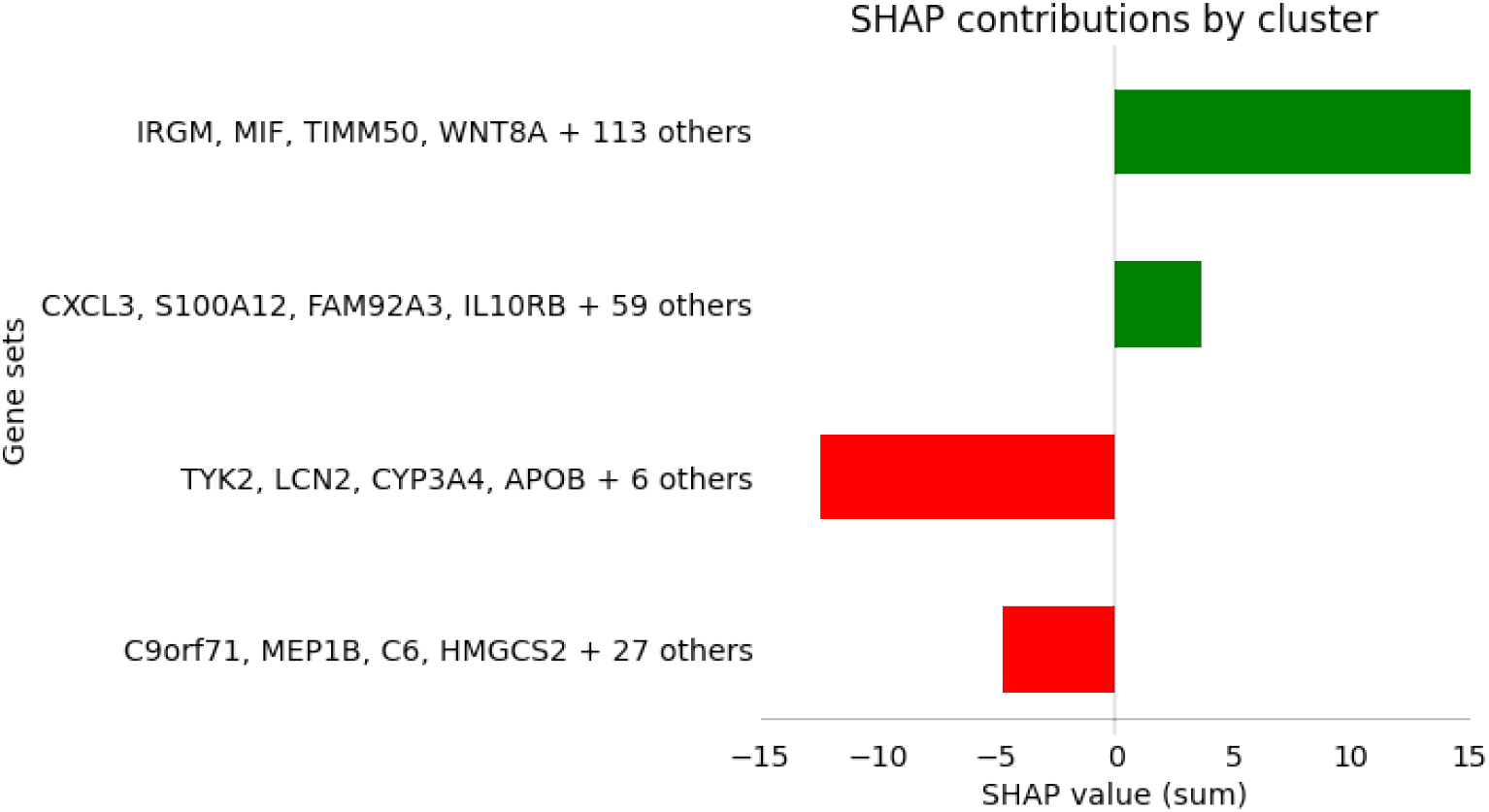
Final gene modules identified as being associated with severe disease (CD deep ulcer), alongside relative contributions determined using SHAP values. Shown is a bar plot in which each gene module is characterised in terms of its influence on the model predicting the “CD deep ulcer” cluster. Each bar represents the sum of mean SHAP values associated with the “CD deep ulcer” cluster, across all genes in the module. We show the top 4 most influential genes of each module. More positive values (green) indicate greater confidence for that module predicting “CD deep ulcer”. More negative values (red) reduce our confidence in a “CD deep ulcer” prediction. Consistent with the literature on IBD, we find genes like IRGM, CXCL3 and IL10RB are present in the modules and have an effect on disease severity.

We again find genes like IRGM, CXCL3 and IL10RB are present in the modules and have an effect on disease severity. This is consistent with the literature on IBD. Using this method, we can determine the type and relative magnitude of influence of each gene module on the model predictions (for each disease subtype).

#### 5.3.1 Gene Ontology (GO) enrichment analysis

We also verify the biological relevance of the identified gene modules using Gene Ontology (GO) enrichment analysis [50, 51, 52] (explained in greater detail in Supp. Material Section 1.1.4, and Tables 3 and 5). For example, many relevant biological processes are enriched in the CD deep ulcer 117-gene module; this is the module that made the greatest positive contribution to predicting CD deep ulcer (topmost bar in Figure 5: a full list of genes in this module is given in Supp. Material Table 3). We show the GO enrichment results for this module in Figure 6 and Supp. Material Table 5. GO analysis of other modules is available in Supp. Material Tables 4 and 6.

**Figure 6:**
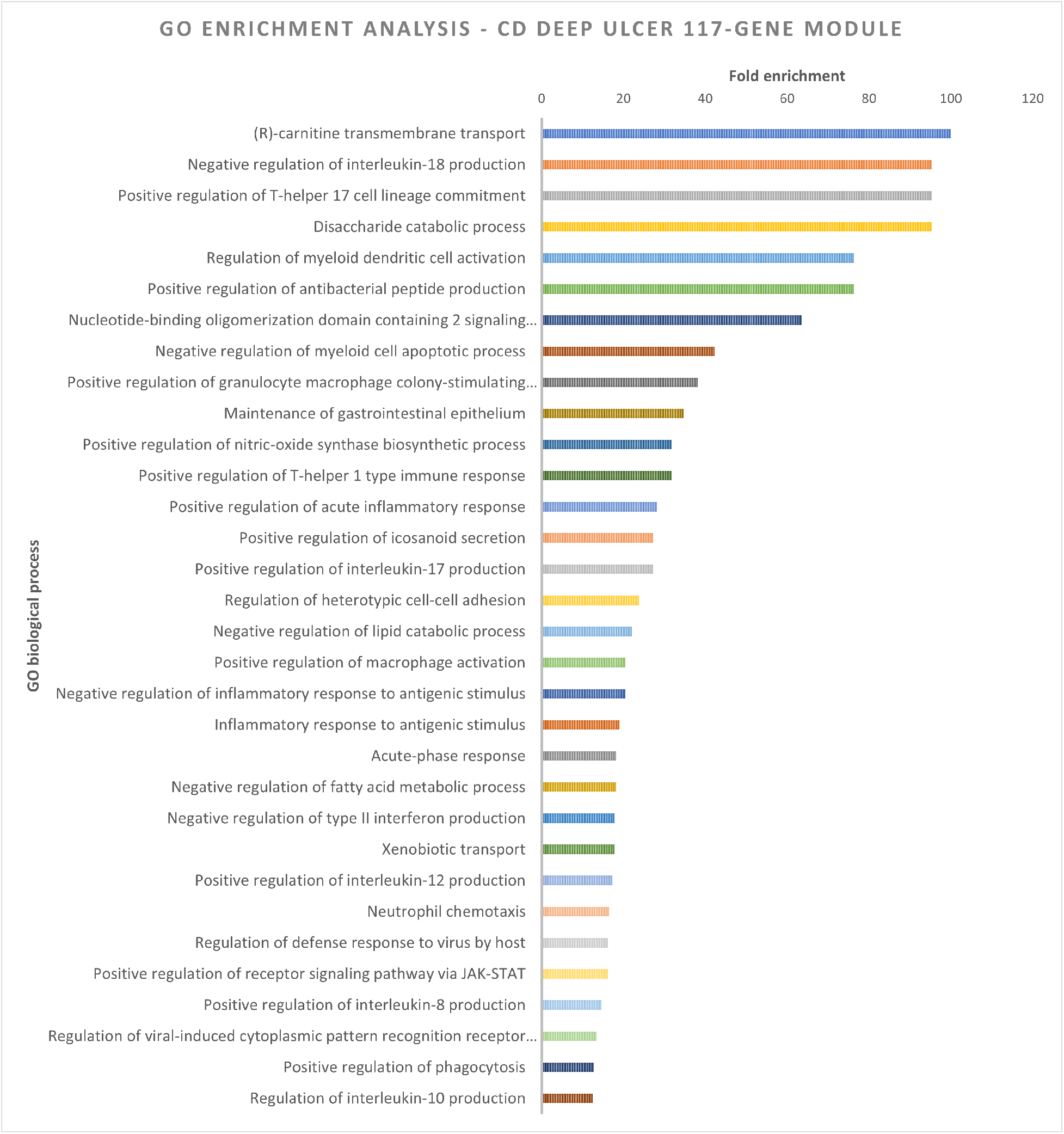
Gene Ontology enrichment analysis. Shown are most enriched biological processes associated with a 117-gene module which was found to be strongly associated with CD with deep ulcer. The GO processes tinclude transport mechanisms, regulation of reactive oxygen species, cell adhesion, and regulation of immune response to viruses and Gram-negative bacteria, which is in line with our current knowledge of IBD. Our results are statistically significant with a false discovery rate (FDR) of less than 0.05 and fold enrichment of up to or exceeding 100 for many of the processes. The most enriched processes are related to T-cell proliferation, transmembrane transport, signalling pathway activation, and production of antibacterial peptides. Full details are given in Supp. Material Section 1.1.4, and Tables 3 and 5.

The GO processes include transport mechanisms, regulation of reactive oxygen species, cell adhesion and regulation of immune response to viruses and Gram-negative bacteria: these are all implicated in IBD. We obtain statistically significant results with low FDR values under 0.05 and fold enrichment approaching or exceeding 100 for many of the processes. Some of the most enriched processes are involved in T-cell proliferation, transmembrane transport, signalling pathway activation and antibacterial peptide production.

This is consistent with our current understanding of IBD, which is characterised by a dysregulated immune response to pathogens [53]. We observe processes regulating the inflammatory response through IL-18 (fold enrichment over 100), IL-8 and IL-10 cytokines, including neutrophil migration, via NF-*κ*B and JAK-STAT signalling pathways [54]. These inflammatory processes are known to be heavily involved in IBD, which validates the large positive contribution of this module in Figure 5: this signifies greater confidence in predicting CD deep ulcers. Furthermore, this is the only module with enriched regulation of reactive oxygen species; this has been specifically associated with CD deep ulcers in the literature [21].

In Section 3.9.1 of the Supplementary Material we explain the GO analysis for other modules, demonstrating their significance in biological processes in IBD. One limitation is that some smaller gene modules do not lead to enriched processes. This is likely because there are not enough genes in the set to confidently infer the correct biological processes. This problem may be addressed by scaling up to larger datasets.

Our findings are consistent with literature, revealing biological processes including inflammatory response, response to cytokines, immune response to bacterial molecules, cell adhesion, leukocyte migration and extracellular matrix organisation [55, 56, 57, 58].

### 5.4 Class-contrastive explainability

Finally, we propose a class-contrastive method as an additional approach to cluster explanation. After identifying gene modules relevant to particular clusters (Section 5(d)), we can demonstrate their impact on disease subtype (CD deep ulcer, CD no ulcer and control) in a visual way. We again utilise the autoencoder for dimensionality reduction, as this leads to better classification performance in comparison to PCA (see Table 1 and Supp. Material Section 3.6).

The expression of each gene (normalised counts), across patients and disease subtypes, follows a Gaussian distribution (Figure 7). We observe that some genes are downregulated or upregulated in patients with CD compared to controls. For example, in Figure 7a, the distributions corresponding to “CD no ulcer” and “CD deep ulcer” are shifted higher compared to the control distribution: this shows upregulation of CXCL3, with higher mean expression levels. However, in Figure 7b, the distribution of the gene MEP1B in patients with CD is shifted lower, compared to controls: this shows downregulation of MEP1B compared to controls.

**Figure 7:**
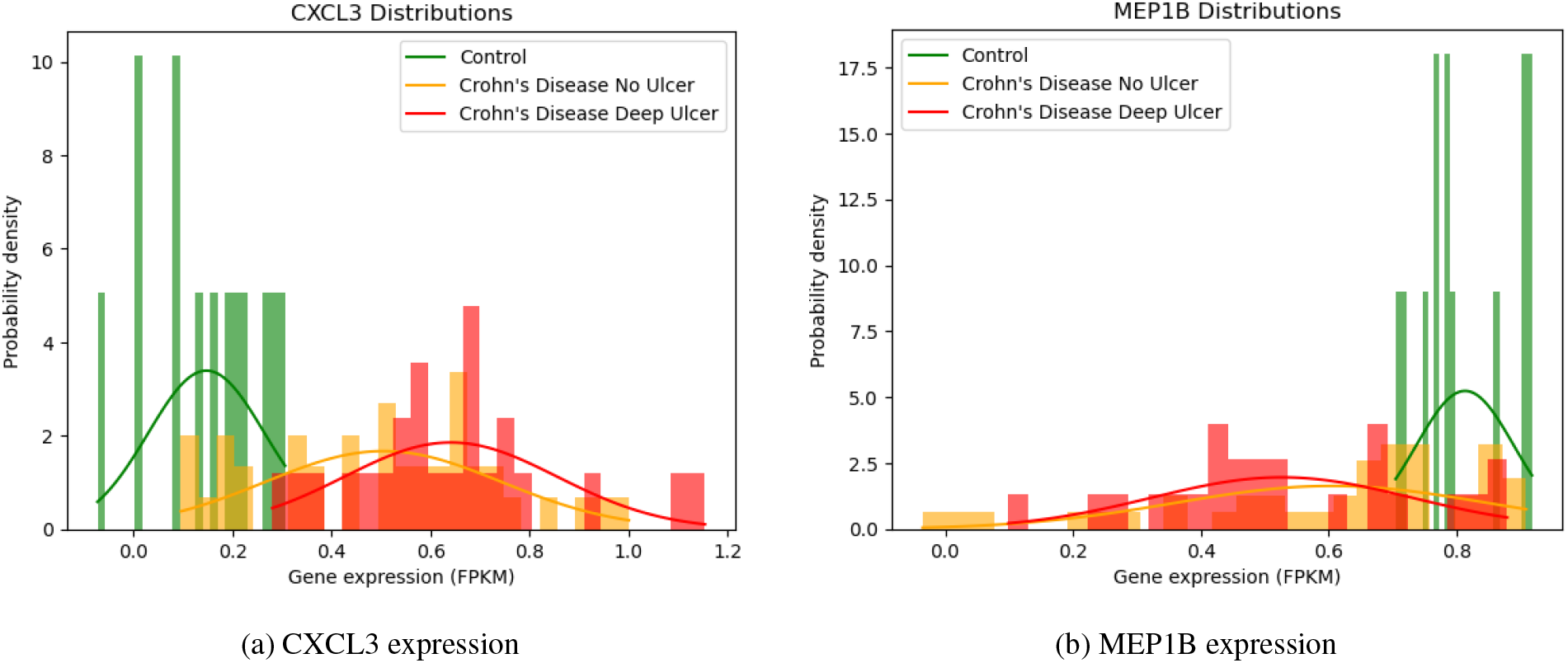
Gene expression distributions of CXCL3 (a) and MEP1B (b) across patients with CD deep ulcer, CD no ulcer and control. The gene expression can be approximated by normal distributions. Data from the RISK dataset [21, 22].

After selecting a patient with CD, we generate a class-contrastive explanation by modifying the expression of particular genes to make the genetic profile more similar to those of controls (patients without CD). This is achieved by changing the expression value of each gene to the mean expression value across control individuals (explained in Section 4(b)(iii) and Eq 6). We can discover the effect of various gene modules, such as those we identified, by modifying their expression in this way. We will demonstrate the process with Patient 46, who is a CD patient with a deep ulcer. Their initial position in the clustering model is shown in Figure 8.

**Figure 8:**
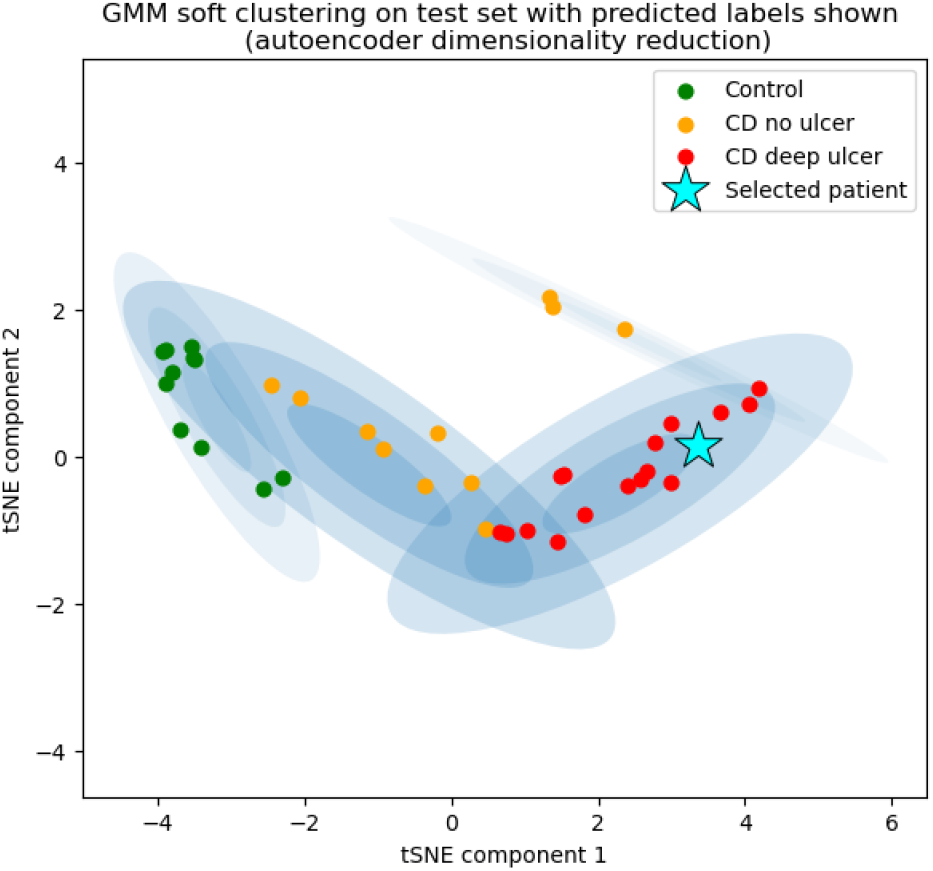
Initial position of Patient 46 (CD deep ulcer) within the clustering model.

We first select the 117-gene module, which was found to make the greatest positive contribution to CD deep ulcer predictions (Figure 5). We modify this set of genes for the patient (using the class-contrastive technique explained above) and refit the model with the same set of parameters. This results in the patient (Patient 46 with CD deep ulcer [a severe form of the disease], Figure 8) being assigned to the “CD no ulcer” cluster [a less severe form of the disease], as shown in Figure 9a. Note that the GMM can change slightly each time we refit the model due to stochasticity in the tSNE algorithm used for visualisation. However, the general structure of the model remains the same. Modifying these genes resulted in the model predicting a less severe form of the disease (CD without deep ulcer). This may suggest that this module contributes to a severe form of the disease (CD with deep ulcer).

**Figure 9:**
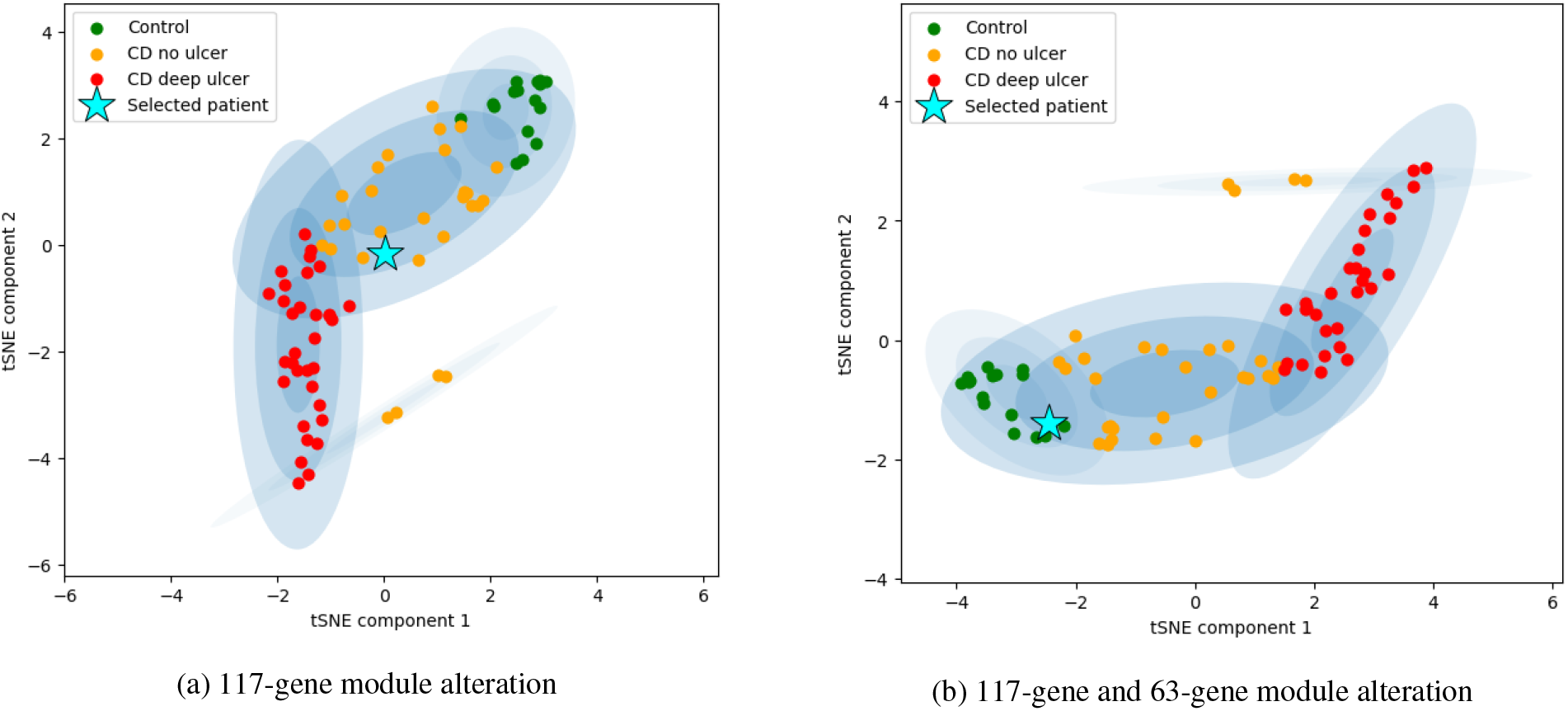
Visual explanations of the effect of modules on disease in a patient. Position of Patient 46 within clustering model after modifying 117-gene module (a) and both 117-gene and 63-gene modules (b), which together were found to account for all positive contribution to CD deep ulcer predictions (Section 5(d)). In (a) modifying the 117 genes using the class-contrastive technique results in Patient 46 (with CD deep ulcer [a severe form of the disease], Figure 8) being assigned to the “CD no ulcer” cluster [a less severe form of the disease]. In (b) modifying both the 117-gene and 63-gene modules using the class-contrastive technique results in Patient 46 moving from the CD deep ulcer cluster to the control cluster. This suggests that these modules may be involved in a severe form of CD that leads to deep ulcers.

Alternatively, we can select the 117-gene module and 63-gene module, which together were found to account for all positive contributions to CD deep ulcer predictions (Figure 5). Modifying these 180 genes and refitting the model results in Patient 46 being assigned to the control cluster (Figure 9b). As the patient has moved from the deep ulcer cluster directly to the control cluster, we can infer that some or all of these 180 genes play a role in a severe form of the disease. We confirmed the functional relevance of these modules by Gene Ontology enrichment analysis (please see Section 5(d)(i) and Supp. Material Section 3.9.1).

When this class-contrastive method is coupled with our gene module identification method, we can visually explore the effect of identified gene modules on the patient’s disease subtype. Although the GMM provides an excellent general representation for stratifying patients, we note that the classifier is not 100% accurate. Our work provides a proof-of-concept; scaling up to larger datasets in future work would likely further improve accuracy and F1-score.

The class-contrastive technique provides intuitive patient-specific explainability. As the identification of gene modules relied on our kernelSHAP extension, all patients and genes were taken into account (including correlations between genes).

## 6 Conclusions and discussion

### 6.1 Summary

We develop techniques to improve the interpretability of models for patient stratification based on genomic data. We adapted and applied machine learning techniques to transcriptomic data from patients with Crohn’s disease (CD), and identified genes and gene modules that are functionally relevant to the disease. We confirm these results using a range of peer-reviewed research, as well as Gene Ontology enrichment analysis. Our novel contributions are summarised below:

- Mixture-based patient stratification and classification into Crohn’s disease (CD) subtypes based on gene expression.
- Adaptation of kernelSHAP for inter-feature dependence; application to Gaussian Mixture Models (GMM) for identification and ranking of genes by disease subtype.
- Data integration technique and consensus clustering [3] to identify potential gene modules by disease subtype. Characterisation of gene modules and confirmation using Gene Ontology (GO) enrichment analysis.
- A class-contrastive technique to visually explain the impact of gene modules on disease subtype for each patient.

### 6.2 Limitations

Our work is not without limitations. Our techniques are not intended for de-novo risk gene identification from a large pool. Rather, given a set of genes and their expression values, we can identify genes and gene modules that influence disease subtype. We also occasionally identify genes, such as SELE, that are not known to have strong links to IBD. Further studies could disprove these as anomalies or confirm them as novel findings.

Our combination of auto-encoder and Gaussian Mixture Model (GMM) is an effective method to stratify patients; however, the method is not 100% accurate. Our work provides a proof-of-concept. Scaling up to larger datasets in future work would likely further improve accuracy and F1-score.

### 6.3 Conclusion

Compared to the state-of-the-art, our methods take gene correlations into account and identify risk genes and gene modules for each CD subtype. This involves adapting a widely used feature attribution algorithm called kernelSHAP, to incorporate inter-feature/gene dependence. Our novel mixture-based classifier uses a probabilistic model derived from our GMM, which captures complex relationships between patient expression profiles and disease subtype.

Our analysis results in many established IBD genes such as NOD2, IRGM, JAK2 and IL10, as well as novel uncharacterised findings like LOC100505851 and LOC100132831. We show the relevance of the identified gene modules and their role in disease by using GO enrichment analysis.

The effect of each gene module on disease subtype can be explained visually using our intuitive class-contrastive technique and gene module analysis. The explainability approach is model-agnostic and can potentially be applied to other diseases. These techniques have the potential for high impact in clinical decision making processes. Our approach may also be useful in other domains where explainability and feature correlations are important, such as financial risk analysis or perception in autonomous vehicle systems.

## Supporting information

supplementary section

## Data Availability

All data is available to download at: https://www.ncbi.nlm.nih.gov/geo/query/acc.cgi?acc=GSE57945

https://github.com/Sharday/Enhancing_patient_stratification_explainable_AI

## Acknowledgements

We would like to thank the paediatric study participants and the National Center for Biotechnology Information (NCBI) for their support in making the IBD RNA-Seq dataset freely accessible to the public.

## Ethics

No ethics approval was necessary. The study used only openly available human data that were originally located at: https://www.ncbi.nlm.nih.gov/geo/query/acc.cgi?acc=GSE57945

## Data accessibility

All data is available to download at: https://www.ncbi.nlm.nih.gov/geo/query/acc.cgi?acc=GSE57945 [22, 21].

## Code availability

All code is available from this repository: https://github.com/Sharday/Enhancing_patient_stratification_explainable_AI

## Author contributions

SO carried out the analysis and implementation, participated in the design of the study and wrote the manuscript. NL carried out the analysis and gave comments on the manuscript. SB carried out the analysis, participated in the design of the study and wrote the manuscript. All authors gave final approval for publication. SB directed the study.

## Funding

We would like to express our gratitude to the Cambridge Trust and Google DeepMind for their generous support through the DeepMind Cambridge Scholarship, awarded to SO for her postgraduate studies at the University of Cambridge. SB acknowledges funding from the Accelerate Programme for Scientific Discovery Research Fellowship. The funders had no role in study design, data collection and analysis, decision to publish, or preparation of the manuscript. The views expressed are those of the authors and not necessarily those of the funders.

### Algorithm 1 Post-processing algorithm for Gaussian Mixture Model (GMM) to classify disease subtype

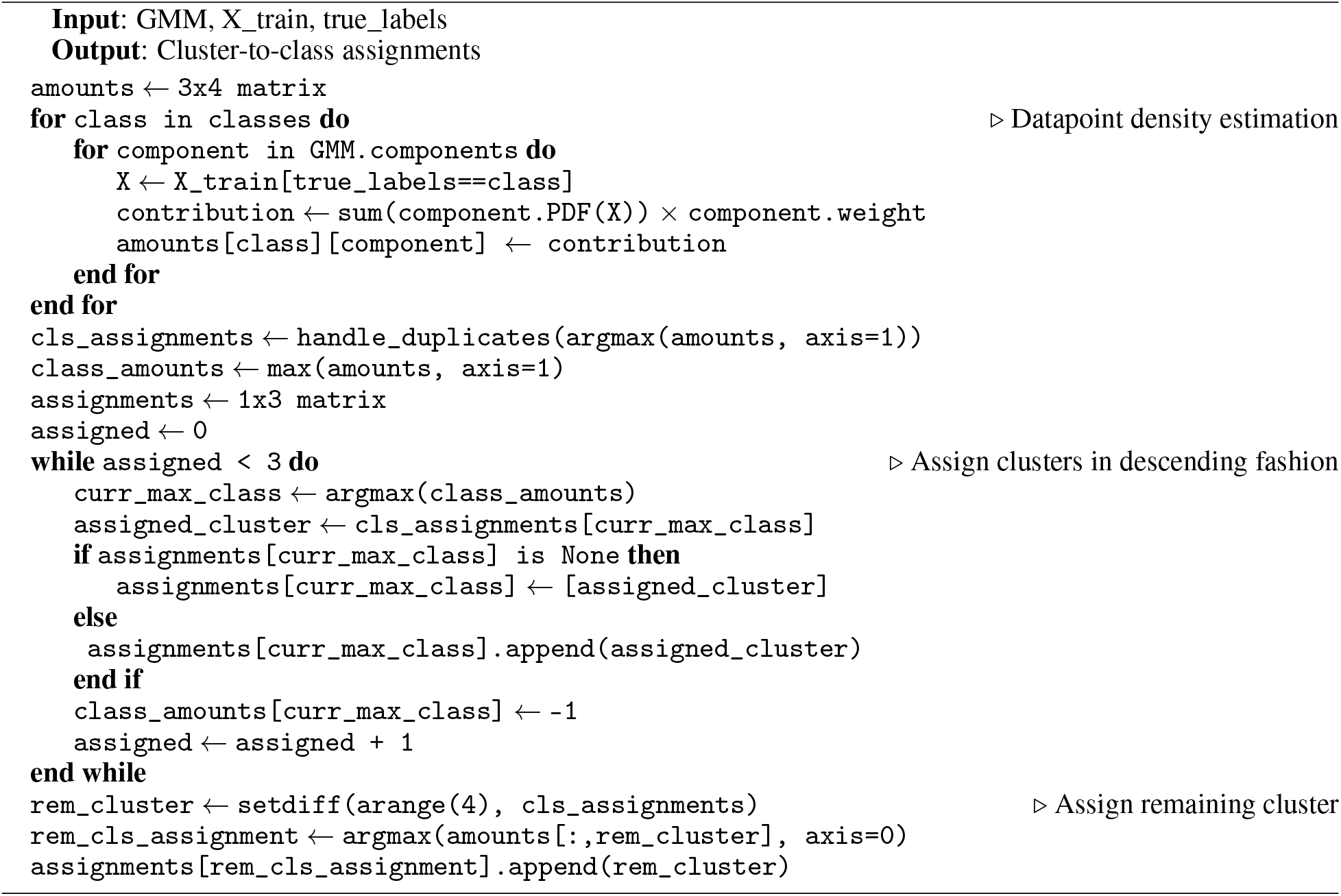

