## supplementary section for "Enhancing patient stratification and interpretability through class-contrastive and feature attribution techniques"

### Supplementary Material: Enhancing patient stratification and interpretability through class-contrastive and feature attribution techniques - unveiling potential therapeutic gene targets

**Sharday Olowu**
University of Cambridge
Cambridge, UK


**Neil Lawrence**
University of Cambridge
Cambridge, UK


**Soumya Banerjee**
University of Cambridge
Cambridge, UK


This document includes supplementary material pertaining to additional background, related work, further results and technical details of the study.

### 1 Background

This section explains the theoretical background relating to the machine learning approaches taken in this work, as well as the biological foundations associated with our main objective.

#### 1.1 Biological foundations

##### 1.1.1 DNA and expression mechanisms

The genome is defined as "the complete set of DNA in an organism" [1]. DNA (deoxyribonucleic acid) is a double-stranded molecule in the nucleus of cells, consisting of four nucleotide bases: adenine, thymine, guanine and cytosine. DNA is divided into sequences called genes, which code for proteins. Genotype describes the constitution of a gene, whereas phenotype refers to observable characteristics, as a result of genotype and interactions with the environment [1]. The expression of each gene is controlled through the regulation of transcription and translation processes [2], which produce proteins from genes.

During transcription of a gene, one DNA strand acts as a template. The enzymes DNA helicase and RNA polymerase separate the strands. As ribonucleotides attach to the complementary DNA bases, RNA polymerase catalyses the production of phosphodiester bonds between the ribonucleotides, forming the RNA transcript. The RNA transcripts are then translated into polypeptides and utilised in biological functions.

##### 1.1.2 RNA-Seq

The transcriptome is the complete set of RNA molecules expressed by the genome. The expression of each gene in the genome is controlled through the regulation of transcription and translation processes [2]. RNA-Seq is a Next-Generation Sequencing technology developed in the mid-2000s, which can be used to study gene expression in organisms [3].

During RNA-Seq, once RNA molecules have been extracted from the tissue, complementary DNA (cDNA) fragments are formed from the transcripts. This involves RNA fragmentation, reverse transcription, adapter ligation and PCR amplification. High-throughput sequencing is then used to obtain a short sequence read from each cDNA. These reads are aligned to a reference genome or transcriptome [3]. Abundance is estimated for each transcript, for example, as reads per kilobase of transcript per million mapped reads (RPKM) [4].

##### 1.1.3 Risk genes and gene modules

Risk genes can predispose an individual to developing certain disorders due to their involvement in known disease processes. Gene modules are sets of genes which have similar expression profiles and are involved in related biological processes, such as metabolic, immune and disease pathways [5]. By identifying and characterising particular gene modules, we can gain insight into their potential roles in disease pathways, leading us to effective therapeutic gene targets.

##### 1.1.4 Gene Ontology enrichment analysis

Gene Ontology (GO) terms refer to a specific biological process, molecular function or cellular component. Genes are annotated to "GO terms" to indicate their involvement in the corresponding processes. Through GO enrichment analysis [6], [7] of a particular gene set, we can discover GO terms that are over-represented in the sample of genes in comparison to the whole genome. We calculate a p-value as the probability of the number of genes in the input list being annotated to a certain GO term (sample frequency), given the number of genes annotated to this term in the reference genome (background frequency). False Discovery Rate (FDR) [8] can also be calculated from p-values. In this work, the GO enrichment analysis tool is used to discover the biological processes associated with detected gene modules.

#### 1.2 Differential expression analysis

Differential expression analysis is a common method currently used to identify potential risk genes. Gene expression data, such as RNA-seq or microarray data, is pre-processed and visualised. Genes that have significantly different expression levels in afflicted patients compared to healthy individuals are identified as genes that potentially contribute to the disease. For example, [9] and [10] compare treatment-naïve IBD patients to healthy controls in this manner, using volcano plots and heatmaps to visualise the data. They also analyse the genes further using functional enrichment analysis, to identify potential associated biological pathways. A similar differential analysis is performed in [11] and [12] for Ulcerative Colitis, identifying potential drivers of disease.

This section summarises some fundamental methods currently used in differential expression analysis, in which gene expression is compared between samples.

##### 1.2.1 Log-fold change

We can find the log-fold change in expression between two samples of tissues or individual cells. This is expressed as a ratio and can be calculated using:

$$log_2(x_2/x_1) \tag{1}$$

where $x_1$ is the normalised count (expression value) for sample 1 and $x_2$ is the normalised count for sample 2 [13].

This shows how much the gene expression differs between the samples on a logarithmic scale. We can identify whether genes are upregulated or downregulated in the second sample compared to the first sample. Gene expression data is often visualised using log-fold change values, for example in hierarchical clustering heatmaps and volcano plots. This helps us to interpret the behaviour of genes and identify outliers.

##### 1.2.2 Welch's t test

The Student's t test is a statistical test that can be used to determine whether a sample of data is significantly different to another sample of data by comparing their means [14].

We can compare the gene expression values of afflicted patients with those of healthy controls to identify significant genes. Welch's t test is an adaptation of the Student's t test. It is suitable for the given application as it is reliable even when the two groups have unequal variance and unequal sample sizes. Like the Student's t test, it assumes that the data in each group is normally distributed [15].

We first define the null hypothesis - that there is no significant difference between the mean values of the two groups [16]. We can compute the t statistic using Eq 2 [15]:

$$t' = \frac{\mu_1 - \mu_2}{\sqrt{\frac{s_1^2}{n_1} + \frac{s_2^2}{n_2}}} \tag{2}$$

where $\mu_i$, $n_i$ and $s_i^2$ are the mean, sample size and sample variance of group i. We then compute the degrees of freedom [15]:

$$v = \frac{\left(\frac{1}{n_1} + \frac{u}{n_2}\right)^2}{\frac{1}{n_1^2(n_1-1)} + \frac{u^2}{n_2^2(n_2-1)}} \tag{3}$$

where

$$u = \frac{s_2^2}{s_1^2} \tag{4}$$

We can use a t-distribution table or software library to find the p-value of the t statistic with the calculated degrees of freedom. The p-value is the probability that we would obtain a t statistic at least as large as what we calculated if the null hypothesis were true i.e. the probability that the results are due to chance [14]. A two-tailed test is appropriate if the mean of one sample may be higher or lower than the other; this results in a two-tailed p-value, which must be halved. If the resulting p-value is below a set threshold e.g. 0.05 or 0.01, we can reject the null hypothesis and state that there is a statistically significant difference between the means, at the given significance level [16].

##### 1.2.3 Volcano plots

Volcano plots are commonly used to visualise RNA-Seq data. We plot the magnitude of the fold-change in gene expression on the x axis, and the significance (p value) on the y axis, where these values are log-transformed. Using this plot, we can easily identify genes that are statistically significant and most differentially expressed in relation to controls. The most upregulated genes will be further to the right and the most downregulated genes further to the left. Genes further to the top have a greater significance [17]. In this work, we assess significance using Welch's t test.

##### 1.3 Dimensionality reduction

As gene expression data often has a large number of variables, the data is usually transformed into a lower dimensional space, while retaining important information, to ease downstream analysis.

###### 1.3.1 Principal Component Analysis

A widely used method for dimensionality reduction of gene expression data is Principal Component Analysis (PCA). This is a statistical technique used to project the data into a lower-dimensional subspace.

As explained in [18], if we have N datapoints $\mathbf{x}_N$, we can define a D-dimensional vector $\mathbf{u}_1$ to be a projection from D dimensions to 1, where the projection of our datapoints is $\mathbf{u}_1^T\mathbf{x}$. We can compute the sample covariance matrix of our datapoints $\mathbf{x}_n$ using:

$$S = \frac{1}{N}\sum_{n=1}^{N}(\mathbf{x}_n - \bar{\mathbf{x}})(\mathbf{x}_n - \bar{\mathbf{x}})^T \tag{5}$$

which summarises the variances and correlations between features. The variance of the projected data is given by $\mathbf{u}_1^T\mathbf{S}\mathbf{u}_1$. By differentiating with respect to $\mathbf{u}_1$ we find that $\mathbf{S}\mathbf{u}_1 = \lambda_1\mathbf{u}_1$, meaning that $\mathbf{u}_1$ is an eigenvector of $\mathbf{S}$ with eigenvalue $\lambda_1$. We can set $\mathbf{u}_1$ as the eigenvector with maximum variance; this is called the principal component. Each subsequent principal component is chosen such that it maximises variance and is orthogonal to all previous principal components. We can use a scree plot to show the amount of variance explained by each principal component.

The original datapoints can then be expressed as a linear combination of the principal components:

$$\mathbf{x}_n = \sum_{i=1}^{D}(\mathbf{x}_n^T\mathbf{u}_i)\mathbf{u}_i = \sum_{i=1}^{D}\alpha_{ni}\mathbf{u}_i \tag{6}$$

By selecting the first M principal components, we can use Eq 6 to project the data from D to M dimensions while maximising variance.

###### 1.3.2 Autoencoders

Autoencoders are a type of neural network that can learn to reconstruct input data using supervised deep learning. In general, they consist of encoder and decoder sections with a bottleneck layer in the middle. By training the network using backpropagation, the autoencoder can learn a feature representation of the input within the bottleneck layer, which is usually the smallest layer in the architecture [19]. It can therefore be used to represent data in a lower-dimensional subspace.

We can write the encoder section as a function $g$ that depends on the input $\mathbf{x_i}$:

$$\mathbf{h_i} = g(\mathbf{x_i}) \tag{7}$$

where $\mathbf{h_i} \in \mathbb{R}^q$ is the latent feature representation of the input [19]. The decoder section can be written as a function $f$ which maps the latent features to the output:

$$\widetilde{\mathbf{x}}_\mathbf{i} = f(\mathbf{h}_i) = f(g(\mathbf{x}_i)) \tag{8}$$

where $\widetilde{\mathbf{x}}_\mathbf{i} \in \mathbb{R}^n$. Therefore, we train the autoencoder to find $f(\cdot)$ and $g(\cdot)$ such that the difference between the input and output is minimised [19]. Since this is a regression problem, a common loss function to use is Mean Squared Error:

$$L_{MSE} = \frac{1}{M}\sum_{i=1}^{M}|\mathbf{x}_i - \tilde{\mathbf{x}}_i|^2 \tag{9}$$

where M is the number of datapoints in the training dataset, $\mathbf{x}_i$ is an input and $\tilde{\mathbf{x}}_i$ is the reconstructed version of the input. This quantifies the difference in the inputs and outputs using squared errors

[19]. Activation functions are also important for introducing non-linearity into the model, such as the rectified linear unit (ReLU) or sigmoid activation function [19].

We can use stochastic gradient descent to train the neural network, in which we repeatedly pass batches of input data through the network, known as a forward pass, calculate the loss and backpropagate the loss through the network to update the weights. One epoch is completed when all training inputs have passed through the network once i.e. one forward and one backward pass [18]. In the backward pass we use the chain rule to calculate the gradient of the loss function with respect to the weights. We can then update the weights of the network using a specified learning rate $\alpha$. Eventually, we converge to a local minimum of the cost function, resulting in a trained network [18]. Autoencoders are useful in a wide range of applications such as feature extraction, image compression, image denoising and dimensionality reduction [20].

##### 1.3.3 t-distributed Stochastic Neighbour Embedding (t-SNE)

A widely used technique for non-linear dimensionality reduction is t-distributed Stochastic Neighbour Embedding (t-SNE) [21]. Most commonly, it is used to reduce the data to 2 or 3 dimensions for visualisation.

We first compute conditional probabilities that represent the similarity between datapoints in high-dimensional space using:

$$p_{i|j} = \frac{exp(-||x_i - x_j||^2/2\sigma_i^2)}{\sum_{k \neq l} exp(-||x_k - x_l||^2/2\sigma_i^2)} \tag{10}$$

As van der Maaten and Hinton explain in [21], "The similarity of datapoint $x_j$ to datapoint $x_i$ is the conditional probability $p_{i|j}$ that $x_i$ would pick $x_j$ as its neighbour if neighbours were picked in proportion to their probability density under a Gaussian centred at $x_i$."

The joint probability of $i$ and $j$ can then be written as:

$$p_{i,j} = \frac{p_{i|j} + p_{j|i}}{2N} \tag{11}$$

where $N$ is the number of datapoints. The Gaussian kernel is set according to the density of the data points. We can learn a mapping from this space to the low-dimensional space by minimising the KL-divergence between the high-dimensional distribution $P$ and a low-dimensional distribution $Q$.

$$C = KL(P||Q) = \sum_i \sum_j p_{ij} log \frac{p_{ij}}{q_{ij}} \tag{12}$$

The $q_{i,j}$ distribution is computed in a similar way as $p_{i,j}$, however $q_{i|j}$ is calculated using the heavy-tailed student t distribution:

$$q_{i|j} = \frac{(1 + ||y_i - y_j||^2)^{-1}}{\sum_{k \neq j}(1 + ||y_k - y_j||^2)^{-1}} \tag{13}$$

With a heavier tail than the Gaussian distribution, points further from the reference point are given more probability mass. This helps to avoid the crowding problem, in which similar points in high-dimensional space are mapped to the same point in low-dimensional space. The optimisation is carried out using gradient descent. The final mapping therefore optimally represents the similarities between high dimensional points in low dimensional space, while preserving the structure of the data.

An important hyperparameter used in this algorithm is perplexity, $Perp(P_i) = 2^{H(P_i)}$. This can be interpreted as "the effective number of neighbours" taken into account when computing conditional probabilities $p_{i|j}$ in high dimensional space. Specifically, binary search is used to find the $\sigma_j$ value that results in a perplexity of the conditional distribution that is close to the target perplexity within a given tolerance. Perplexity can have a significant impact on the mapping, shifting emphasis in terms of the local and global structure of the data [22].

#### 1.4 Clustering algorithms

This section explains the clustering algorithms and evaluation metrics used in this work.

##### 1.4.1 K-Means clustering

K-Means is an iterative algorithm used to assign each datapoint $x_n$ to one of $k$ clusters. It aims to minimise the objective function (Eq 14), which represents the sum of squared distances from each datapoint to the centre of its assigned cluster $\mu_k$:

$$J = \sum_{n=1}^{K} \sum_{k=1}^{K} r_{nk} ||\mathbf{x}_n - \boldsymbol{\mu}_k||^2 \tag{14}$$

The variable $r_{nk}$ is used as an indicator, where $r_{nk} = 1$ if datapoint $x_n$ is assigned to cluster $k$ and 0 otherwise. The aim is to find the optimal $r_{nk}$ and $\mu_k$ that minimises $J$ [18]. The key points of the algorithm are as follows:

1. Select a number of $k$ clusters with which to perform clustering.
2. Randomly initialise the centroids (centres) for each cluster.
3. Assign each datapoint to the closest cluster, using a distance metric like Euclidean distance.
4. Assign new centroids for each cluster, by calculating the mean of the newly assigned datapoints.
5. Repeat steps 3 and 4 until convergence or the maximum number of iterations is reached.

The optimisation steps 3 and 4 correspond to the Expectation (E) and Maximisation (M) steps of the Expectation-Maximisation (EM) algorithm respectively. The EM algorithm is widely used in probabilistic models for finding maximum-likelihood estimates of parameters [18]. During the E step, we fix the cluster centres and assign each datapoint to the closest cluster using $r_{nk}$.

$$r_{nk} = \begin{cases} 1, & \text{if } k = argmin_j ||\mathbf{x}_n - \boldsymbol{\mu}_j||^2 \\ 0, & \text{otherwise} \end{cases} \tag{15}$$

During the M step, we fix the indicator variable and recalculate the cluster centroids by finding the mean of the datapoints in each cluster $k$:

$$\boldsymbol{\mu}_k = \frac{\sum_n r_{nk} \mathbf{x}_n}{\sum_n r_{nk}} \tag{16}$$

At completion each datapoint is assigned to one of the $k$ clusters [18].

##### 1.4.2 Gaussian Mixture Models

Another way to perform clustering is using Gaussian Mixture Models (GMMs). This method involves finding a probability distribution over clusters using a weighted average of Gaussian distributions [18].

The marginal distribution of a GMM can be expressed as $p(\mathbf{x}) = \sum_{\mathbf{z}} p(\mathbf{z}) p(\mathbf{x}|\mathbf{z}) = \sum_{k=1}^{K} \pi_k \mathcal{N}(\mathbf{x}|\boldsymbol{\mu}_k, \boldsymbol{\Sigma}_k)$. Each component $k$ is represented using a multivariate Gaussian distribution with mean vector $\mu_k$ and covariance matrix $\Sigma_k$. The mixing coefficients $\pi_k$ are associated with a K-dimensional random variable $z$ that indicates cluster membership, such that $z_k \in \{0, 1\}$ and $p(z_k = 1) = \pi_k$. We can sample from this distribution using ancestral sampling i.e. sample from $p(\mathbf{z})$ and then from $p(\mathbf{x}|\mathbf{z})$ [18].

We can also define the responsibility as $\gamma(z_k) \equiv p(z_k = 1|\mathbf{x})$ which describes the responsibility of component k for 'explaining' the given datapoint **x**. Given a set of datapoints, we can use maximum-likelihood estimation to find the parameters of the distribution to best model the data. We aim to maximise the log of the likelihood function:

$$\ln p(\mathbf{X}|\boldsymbol{\pi}, \boldsymbol{\mu}, \boldsymbol{\Sigma}) = \sum_{n=1}^{N} \ln \left\{ \sum_{k=1}^{K} \pi_k \mathcal{N}(\mathbf{x}_n|\boldsymbol{\mu}_k, \boldsymbol{\Sigma}_k) \right\} \tag{17}$$

There is no closed-form solution for this purpose for GMMs, so we use the iterative Expectation-Maximisation algorithm [18]. During the E step, we use the current parameter values of $\boldsymbol{\mu}_k$, $\boldsymbol{\Sigma}_k$ and $\boldsymbol{\pi}_k$ to calculate responsibilities using Eq 18:

$$\gamma(z_k) = \frac{\pi_k \mathcal{N}(\mathbf{x}_n|\boldsymbol{\mu}_k, \boldsymbol{\Sigma}_k)}{\sum_{j=1}^{K} \pi_j \mathcal{N}(\mathbf{x}_n|\boldsymbol{\mu}_j, \boldsymbol{\Sigma}_j)} \tag{18}$$

We use these calculated responsibilities to re-estimate the parameters during the M step using the following equations:

$$\boldsymbol{\mu}_k^{new} = \frac{1}{N_k}\sum_{n=1}^{N} \gamma(z_{nk})\mathbf{x}_n \tag{19}$$

$$\boldsymbol{\Sigma}_k^{new} = \frac{1}{N_k}\sum_{n=1}^{N} \gamma(z_{nk})(\mathbf{x}_n - \boldsymbol{\mu}_k^{new})(\mathbf{x}_n - \boldsymbol{\mu}_k^{new})^T \tag{20}$$

$$\pi_k^{new} = \frac{N_k}{N} \tag{21}$$

where $\gamma(z_{nk}) = p(z_k = 1|\mathbf{x}_n)$ and $N_k = \sum_{n=1}^{N} \gamma(z_{nk})$. The E step and M step are successively repeated until we find a local maximum for the log-likelihood [18]. The final parameters are then used to construct the GMM. This provides a soft clustering of the given datapoints.

The K-Means algorithm assumed that the clusters spread out evenly in all directions. By contrast, GMMs can handle non-spherical clusters, and the covariance can be tuned to form clusters that fit the shape of the data more closely [18].

##### 1.4.3 Consensus clustering

Consensus clustering is an approach that is gaining popularity as a more reliable alternative to vanilla clustering algorithms. It involves performing several partitionings of the same dataset and uses a function to find a consensus among these, resulting in a clustering that can outperform each individual partitioning in accuracy and stability [23], even in the presence of noise, outliers and sample variations [24].

After obtaining a set of basic partitionings, $\Pi = \{\pi_1, \pi_2, ..., \pi_r\}$, of the data objects, $\mathcal{X} = \{x_1, x_2, ..., x_n\}$, the goal of consensus clustering is to find a consensus partitioning $\pi$ such that we maximise:

$$\Gamma(\pi, \Pi) = \sum_{i=1}^{r} w_i U(\pi, \pi_i) \tag{22}$$

where $\Gamma : \mathbb{N}^n \times \mathbb{N}^{nr} \to \mathbb{R}$ is a consensus function, $U : \mathbb{N}^n \times \mathbb{N}^n \to \mathbb{R}$ is a utility function and $w_i \in \mathbb{R}_{++}$ is the weight specified for $\pi_i$ by the user [24].

In this work, we utilise the Weighted Ensemble Consensus of Random (WECR) K-Means algorithm [23] for the identification of potential gene modules.

##### 1.4.4 Hierarchical clustering

Hierarchical clustering takes either a top-down (divisive) or bottom-up (agglomerative) approach. In the bottom-up approach, each datapoint starts off as an individual cluster and at each step, pairs of clusters are merged according to a similarity metric and linkage method. The top-down approach starts with all datapoints as part of a single cluster and splits clusters recursively until we obtain individual datapoints [25]. The type of linkage, such as single, complete, average, Ward or centroid linkage [26], [27], describes how we define the distance between two clusters. In this work, hierarchical agglomerative clustering is employed with Euclidean distance, using average linkage. This means clusters with the lowest average distance are merged, where this distance is calculated as the average of distances between all possible pairs of datapoints. The final clustering can be visualised using a dendrogram, which shows the history of merges and the similarity between clusters [25].

##### 1.4.5 Evaluation metrics

There are many possible techniques that can be used to evaluate clustering results. They can be used to select optimal models in both individual and consensus clustering.

**Silhouette analysis** Through silhouette analysis, we can assess the separation distance between clusters using the Silhouette Coefficient. The Silhouette Coefficient $s$ for a sample $x_i$ in cluster $k$ can be calculated using Eq 23:

$$s(x_i) = \frac{b(x_i) - a(x_i)}{max(b(x_i), a(x_i))} \tag{23}$$

where $a(x_i)$ is the mean distance from the sample to all other points in the same cluster $k$, and $b(x_i)$ is the mean distance from the given sample to all points in the next closest cluster [28]. The silhouette value can vary between -1 and 1. Values close to 1 indicate well-separated clusters, whereas values close to 0 show that datapoints are near the decision boundary. As values become closer to -1 there is a greater probability that some points may be in the incorrect cluster, as it shows that $a(x_i)$ is greater than $b(x_i)$. Higher silhouette values indicate that the datapoints are more likely to be clustered correctly [28]. A silhouette plot [29] [30] can be used to visualise silhouette values across all samples in the dataset after clustering. The thickness of a bar shows the size of a cluster and we can see how the silhouette values vary across the samples within and between clusters. This allows us to assess the separability and quality of clustering [29].

**Davies-Bouldin Index** The Davies-Bouldin Index (DBI) evaluates the separation distance between pairs of clusters, which should be as large as possible, as well as within-cluster scatter, which should be as small as possible [31]. It is defined as:

$$DBI = \frac{1}{K} \sum_{i=1}^{K} \max_{i;j \neq i} \frac{S_i + S_j}{d_{i,j}} \tag{24}$$

where $K$ is the number of clusters and $d_{i,j}$ is the distance between clusters $i$ and $j$. The scatter within a cluster is given by $S_i = \frac{1}{|C_i|} \sum_{x_j \in C_i} ||x_j - v_i||$, where $C_i$ refers to cluster i, $x_j$ refers to a datapoint assigned to cluster i and $v_i$ is the centroid of cluster $i$. The DBI can range from 0 to infinity. Better clustering models are indicated by lower DBI values, as these indicate that the clusters are compact and separated well [31].

**Calinski-Harabasz Index** The Calinski-Harabasz Index (CH index) evaluates a clustering model using between-cluster variance, which measures separation distance between clusters, and within-cluster variance, showing how tightly packed each cluster is [32]. It is defined in Eq 25.

$$CH(K) = \frac{B(K)(N - K)}{W(K)(K - 1)} \tag{25}$$

$$B(K) = \sum_{k=1}^{K} a_k ||\overline{x_k} - \bar{x}||^2 \tag{26}$$

$$W(K) = \sum_{k=1}^{K} \sum_{C(j)=k} ||x_j - \overline{x_k}||^2 \tag{27}$$

Here, K is the number of clusters and N is the sample size. $B(K)$ is the between-cluster variance and should be as large as possible, whereas $W(K)$ is within-cluster variance and should be as small as possible. The CH index is the ratio between these measures, ranging from 0 to infinity. Higher CH index values indicate a greater quality of clustering as it shows that the clusters are better separated and more compact [32].

**Bayesian Information Criterion** The Bayesian Information Criterion (BIC) is metric used to select the best model from a collection of candidate models $M_k$ for $k \in \{k_1, ...k_L\}$. BIC is useful in clustering for choosing the model that fits the data best [33]. The BIC for model $M_k$ is defined as:

$$BIC = k \ln(n) - 2 \ln L(\hat{\Theta}_k | x) \tag{28}$$

where $n$ is the number of samples, $k$ is the number of parameters, and $\hat{\Theta}_k$ is the set of model parameters that maximises the likelihood of the data $L(\hat{\Theta}_k | x)$. The term $k \ln(n)$ penalises complex models with large numbers of parameters. The model with the minimal BIC value is said to be optimal as it has the best balance between model fit and complexity [33].

**Accuracy and F1-Score** After post-processing our KMeans and GMM clustering results, we obtain cluster assignments that correspond to disease phenotype predictions. As a result, we can assess classification performance using accuracy and F1-score.

These rely on four components (for binary classification) [34]:

- True Positives (TP): Number of samples correctly predicted as positive.
- False Positives (FP): Number of samples incorrectly predicted as positive.
- True Negatives (TN): Number of samples correctly predicted as negative.
- False Negatives (FN): Number of samples incorrectly predicted as negative.

where one class is assigned as "positive" and the other "negative".

Accuracy [35] can be calculated using:

$$accuracy = \frac{TP + TN}{TP + FP + TN + FN} \tag{29}$$

However, this measure can produce unreliable results when class sizes are imbalanced.

Therefore, accuracy is often used in conjunction with F1-score, which is the harmonic mean of precision and recall [34]:

$$Precision = \frac{TP}{TP + FP} \tag{30}$$

$$Recall = \frac{TP}{TP + FN} \tag{31}$$

$$F1\text{-}Score = \frac{2 \times Precision \times Recall}{Precision + Recall} \tag{32}$$

Recall is used to identify what proportion of the true positive instances were identified by the model, whereas precision shows the proportion of positive predictions that were correct. There is often a trade-off between precision and recall. F1-score combines these metrics and is robust to class imbalance. This work uses a weighted F1-score; this is a weighted sum of the class-wise F1-scores, in which the weights are proportional to the class size [34].

##### 1.5 Explainability

It is often difficult to determine how decisions are made in machine learning models, particularly when complexity is high. The ability to explain the reasoning behind machine learning model predictions is very important in sensitive fields such as law and healthcare. For example, it is critical for healthcare practitioners to know that any insights drawn or suggestions made are grounded in reality with sound judgement. Moreover, legal requirements are beginning to restrict the use of uninterpretable "black-box" models in sensitive domains due to a lack of transparency [36], [37]. This section describes two state-of-the-art methods for machine learning explainability: class-contrastive techniques and SHAPley Additive exPlanations (SHAP).

##### 1.5.1 Class-contrastive techniques

Class-contrastive reasoning is widespread in the social sciences. Studies in human cognition [38] have revealed that explanations are inherently contrastive; justifications for a belief or action are usually desired in contrast to another i.e. "Why P rather than Q?", where Q may be implied by the context [39].

This approach is now being applied in state-of-the-art machine learning research to reveal which factors led to a model's decision. The idea is to explain why the model made the given decision in contrast to another. For example, as demonstrated by Banerjee et al. in [40], a model may determine that a patient is at high risk of mortality due to their dementia and cardiovascular disease, whereas the risk would be much lower if they were not suffering from these diseases. By allowing us to distinguish predictions based on specific features, the transparency and interpretability of a model can be greatly improved.

##### 1.5.2 SHAPley Additive exPlanations (SHAP)

SHAPley Additive exPlanations (SHAP) [41] is a state-of-the-art method for explaining machine learning model predictions, using a game theory approach for feature attribution.

The classic SHAP method formulates the problem as a game in which each feature is a player and makes contributions to the outcome, which is the model prediction. A new model is trained for every possible coalition (subset) of features $S \subseteq F$. We can find the contribution of a feature by finding the prediction of a model trained with the feature included in the coalition $f_{S\cup\{i\}}(x_{S\cup\{i\}})$, and comparing this to the prediction of a model trained with the feature withheld $f_S(x_S)$. The Shapley value $\phi_i$ of a feature $i$ is a weighted average of the contribution of that feature across all possible coalitions:

$$\phi_i = \sum_{S\subseteq F\setminus\{i\}} \frac{|S|!(|F|-|S|-1)!}{|F|!}[f_{S\cup\{i\}}(x_{S\cup\{i\}}) - f_S(x_S)] \tag{33}$$

Because we have to train a new model for all possible subsets of features, the classic SHAP method can become computationally intractable when the model includes a significant number of features. Variants of this algorithm have been proposed to approximate the original method. Two notable variants are treeSHAP and kernelSHAP. TreeSHAP is the more efficient of the two, but can only be used for tree-based machine learning models such as decision trees or random forests [42]. By contrast, kernelSHAP is model-agnostic.

KernelSHAP provides an efficient approximation to SHAP values using weighted linear regression based on sampling. Rather than retraining a new model for each coalition, we marginalise the missing features out of the model. In Eq 34, we define a fidelity function $L$ that measures how unfaithful is a surrogate model $g$ in approximating the model $f$, in the feature subspace defined by $z'$. Here, we use $z' \in \{0,1\}^M$ to define the coalition of features, where $M$ is the number of input features. The features included in the coalition have a corresponding value of 1 and missing features are represented by a value of 0. We carry out a sum of the loss calculated over all models.

$$L(f, g, \pi_{x'}) = \sum_{z'\in Z} [f(h_x(z')) - g(z')]^2 \pi_{x'}(z') \tag{34}$$

We generate synthetic samples for each model, where each baseline sample $z$ is drawn from the same probability distribution as the input features. We can compute the model output $f(h_x(z'))$ as $E[f(z)|z_S] = E_{z_{\bar{S}}|z_S}[f(z)]$. However, feature independence is assumed so $f(h_x(z')) \approx E_{z_{\bar{S}}}[f(z)] \approx f([z_S, E[z_{\bar{S}}]])$. This means we simulate missing features using expectation values, to show that these features carry no information. We use $z'$ to represent a perturbed version of the sample $z$, where the included features take their value from the input instance we are analysing. The $h_x$ function is used to map the samples to a potentially higher-dimensional space.

The kernel weighting function (Eq 35) is used to penalise coalitions where the number of features is far from zero or M. When $|z'|$ is close to zero this demonstrates the independent effects of features, whereas when $|z'|$ is close to M, this shows how features interact with each other [43].

$$\pi_{x'}(z') = \frac{(M-1)}{(Mchoose|z'|)|z'|(M-|z'|)} \tag{35}$$

We then perform linear regression to minimise the fidelity function $L$. This gives rise to a linear equation, in which the resulting coefficients are the SHAP values of the corresponding features:

$$g(z') = \phi_0 + \sum_{i=1}^{M} \phi_i z'_i \tag{36}$$

The SHAP value represents the importance of a feature in terms of its influence on the model prediction. SHAP values are additive and can be summed, as shown, to approximate the output of the model for a given data instance.

### 2 Related work

The following is a summary and critique of existing literature related to the techniques employed in this work.

#### 2.1 Dimensionality reduction and cluster analysis

RNA-Seq datasets can be very high-dimensional due to the number of genes involved, further complicated by noise. It is therefore common practice to reduce the dimensionality using feature selection [44] or feature extraction [45] after data pre-processing, before performing further analysis. Feature selection can be more interpretable, as a subset of the original genes are selected for analysis, often using statistical tests. However, feature extraction can reduce the dataset to a very small number of variables by transforming the original features, while maximising variance. This is useful for larger datasets and can allow different types of patterns to be detected. A technique widely used for feature extraction is Principal Component Analysis (PCA). This has performed very well throughout the past few decades for analysing gene expression data [45]–[48]. However, PCA only captures linear relationships within the data, and as demonstrated in [46], it can fail to detect important biological information. This is more apparent with small sample sizes and when effect size is small i.e. phenotype is affected by very small changes in gene expression. It can also be difficult to determine which principal components contain relevant information [46].

Recently, deep learning models such as autoencoders have shown compelling performance in the analysis of high-dimensional single-cell RNA-Seq data [49]–[52]. Autoencoders can improve the signal-to-noise ratio [52] as the structure forces the model to learn an effective representation within a small number of latent variables in the bottleneck layer. Autoencoders also capture non-linear relationships within the data and can outperform PCA for dimensionality reduction and clustering, as demonstrated in [52]. This research has mostly focused on single-cell RNA-seq. We aim to reveal the strengths and weaknesses of the PCA and deep autoencoder techniques in the analysis of bulk RNA-Seq data.

Cluster analysis has been successful in discovering disease subtypes [53]–[56], cell types [57], [58] and drug development [59]–[61]. Apart from classical methods like KMeans and hierarchical clustering [62]–[66], other model-based and novel machine learning approaches are being applied for cluster analysis. For example, a multivariate Poisson-log normal mixture model is used in [67], a form of neuralised clustering proposed in [68] and genes are clustered using count-based correlations and dispersion estimation in [69]. However, in these works, clustering is usually carried out directly on genes to form groupings. We instead take the approach of grouping tissue samples based on the similarity of individual patient expression profiles, extracting gene modules at a later stage using a more involved process.

Although clustering is effective for exploratory analysis, it can be useful to anchor this to concrete data. Classifiers are usually trained separately, for example using Random Forests [70] or Support Vector Machines [71]. To our knowledge, there have been no works that adapt a mixture-based clustering model for classification of disease phenotype based on RNA-Seq data. We apply this method for interpretable analysis and seamless coupling to explainability techniques.

#### 2.2 Explainability

Explainable AI (XAI) is becoming increasingly important for sensitive applications. Two important approaches to XAI are feature attribution and class-contrastive techniques. Feature attribution methods are used to calculate the degree to which each feature contributes to the model prediction. These include Shapley Additive Predictions [41], LIME [72] and Anchors [73]. The class-contrastive approach uses counterfactual-based examples to explain why a data instance would be placed in one class over another, also utilising features.

Explanations can be local, to analyse predictions for particular data instances, or global, to explain the systematic behaviour of the model in general. Our work focuses on class-contrastive techniques and SHAP, including local and global explanations for the identification of gene targets.

##### 2.2.1 SHapley Additive exPlanations: applications

SHapley Additive exPlanations (SHAP) [41] is a state-of-the-art method for generating explanations via feature attributions. There are many implemented variants of SHAP, such as kernelExplainer, treeExplainer and gradientExplainer [74], which can offer faster approximations and versions of the algorithm tailored for specific types of models. It can therefore be used in a wide range of applications, and has been applied successfully for the analysis of gene expression data [75]–[79]. For example, Yap et al. demonstrate the utility of SHAP when applied to a tissue classifier in [75]. The SHAP GradientExplainer is suitable for neural networks and was used to find the individual contributions of genes to predictions. The most important genes identified by SHAP were congruent with those identified by differential expression analysis, and were associated with the expected biological processes when applying functional enrichment analysis. It can even be used to find the relative significance of regulatory pathways, as shown by Hayakawa et al. in their application of SHAP to a graph convolutional network classifier that predicts diffuse large B-cell lymphoma (DLBCL) subtypes [76].

More specifically related to this work, Yu et al. use a deep autoencoder in [77] to learn gene expression representations, applying treeExplainer SHAP to measure the contributions of genes to each of the latent variables. During functional enrichment analysis, the most important genes distinguished by SHAP led to the identification of many more enriched pathways than those genes identified by differential expression analysis. The use of an autoencoder to learn representations is similar to our work, but in [77], SHAP is applied directly to the hidden layer, which limits interpretability of the findings. In this work, we enhance interpretability in relation to disease phenotype, by applying SHAP to our mixture model. As explained in the main document, we use a novel approach to incorporate inter-feature dependence into kernelSHAP for more robust explanations.

#### 2.3 Gene module identification

The identification of gene modules is a crucial step in characterising the genetic component of disease.

Weighted Gene Co-expression Network Analysis (WGCNA) [80], [81] was proposed by Zhang et al. in 2005. This involves the use of a weighted gene co-expression network and hierarchical clustering. It has been applied in many works to identify potential gene modules and centralised hub genes as biomarkers for various diseases [82]–[84]. However, it can be sensitive to noise and the results can be highly dependent on the choice of parameters [5], such as the soft-thresholding parameter which controls correlations [80].

Other methods also tend to include a clustering aspect and/or network construction [85]–[87] to organise genes. Zhang et al. propose to combine two well-known algorithms in [85] to identify gene modules involved in hepatocellular carcinoma. The Newman algorithm is used to build a gene co-expression network, before applying the KMeans algorithm for secondary clustering. This approach optimises for the modularity of gene sets but does not attempt to quantify the contribution of each gene set to disease phenotype or progression. The computational complexity may also limit scalability to larger datasets. Our approach employs clustering but avoids the computational costs associated with network construction. Instead, we capture complex gene and sample relationships implicitly via the use of mixture modelling and a deep autoencoder that can infer both linear and non-linear relationships.

More specifically related to this work, in [68], Lu et al. propose an integrated deep learning framework that uses a deep autoencoder for dimensionality reduction of single-cell RNA-Seq data and reformulates the K-Means clustering procedure using a neural network. They use an adversarial approach to identify sets of genes that can explain differences between clusters. This is achieved by identifying perturbations of gene expression profiles that cause cells to move from one cluster to another. This yields lists of genes that can explain a cluster or pair of clusters. However, the underlying gene expression distributions are not taken into account. By comparison, our novel class-contrastive technique uses information about healthy expression profiles to inform perturbations. This improves efficiency and promotes more realistic cluster explanations.

The authors claim to account for gene dependencies in [68] by jointly handling the non-linear embedding and neuralised clustering. This not explained further and is validated only on synthetically generated dependent genes. Our approach explicitly accounts for inter-feature dependence by analysing the underlying data distributions and correlations between genes using data from real patients. Our approach also leads to more interpretable findings, as we use a probabilistic model derived from a GMM that captures relationships between phenotypes. Although [68] produces cluster-wise rankings on genes, it does not take account of how expression level can affect gene relevance, a useful aspect of our approach. It is also not possible for a sample to be associated with more than one cluster. Using a Gaussian Mixture Model, we provide a richer representation of sample relationships and a verifiable probabilistic model. The application of SHAP then provides specific gene contributions for each patient by phenotype, which can be combined for cluster-wise or global explanations.

### 3 Further results and technical details

#### 3.1 Differential expression analysis

As a preliminary analysis of the data selected from the RISK dataset [10], [88], we produced a volcano plot, shown in Figure 1, on the basis of Welch's t test [15]. Various thresholds can be used to select different subsets of genes, based on significance and/or extent of fold-change.

We then carried out hierarchical agglomerative clustering with average linkage and Euclidean distance on the entire sample i.e. 260 patients and 221 genes. The results are shown in Fig 2, generated using the seaborn library [89]. The darker the colour, the more downregulated the gene is and a lighter colour signifies greater upregulation. The clustering results in groups of patients with similar expression profiles as well as distinct groups of genes with similar expression patterns. These could potentially correspond to gene modules.

Using a volcano plot threshold of 1E-25 for significance level and 2.2 for absolute fold-change, we selected a sample of the most significant genes. The expression of these 87 genes is visualised in Figure 3, across a random subset of 30 patients. We can see that the correlations in gene expression patterns still hold, and that the extent of differential expression becomes greater as symptoms become more severe. For example, in general, Crohn's disease (CD) deep ulcer patients have the greatest degree of downregulation and upregulation of these genes, followed by CD no ulcer patients, followed by healthy controls. This signifies that these genes may contribute to the development of CD, and that the extent of differential expression may be implicated in symptom severity. However, some genes were not identified to be significant using the t test, but are linked to IBD in the literature, such as IRGM, HLA_DRB1 and IL10. This indicates that there may be other factors at play.

Differential gene expression analysis is informative in a broad sense, but can be too simplistic to accurately capture the nuances of the mechanisms underlying disease. When comparing only the expression of individual genes, it is difficult to draw specific conclusions. Other important factors are not considered, such as gene dependencies. Our work aims to address this by associating differential expression with disease phenotype in a more in-depth way, utilising and extending state-of-the-art machine learning explainability techniques.

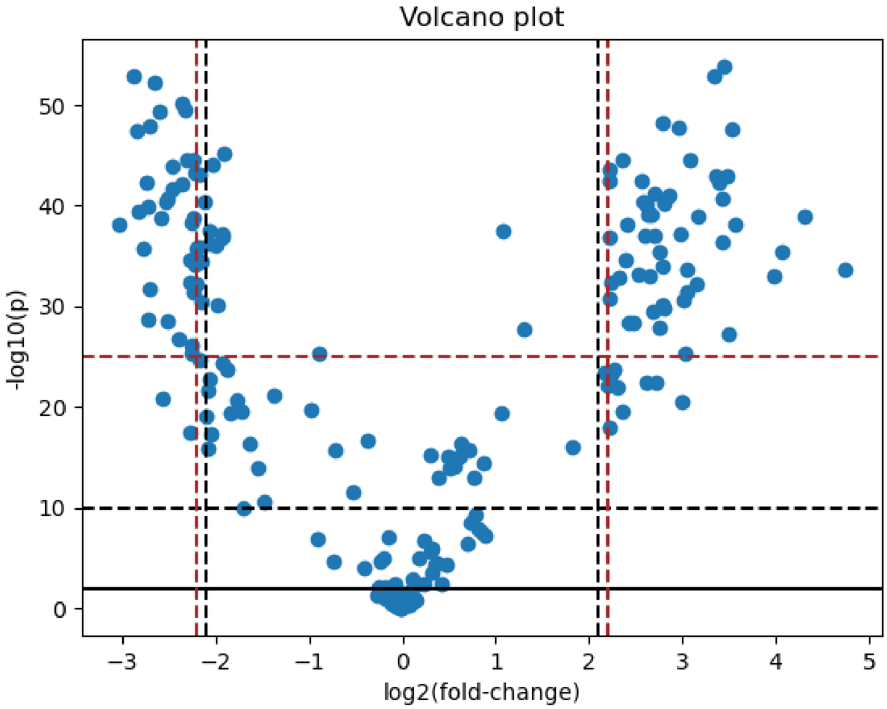

Figure 1: Volcano plot of gene sample.

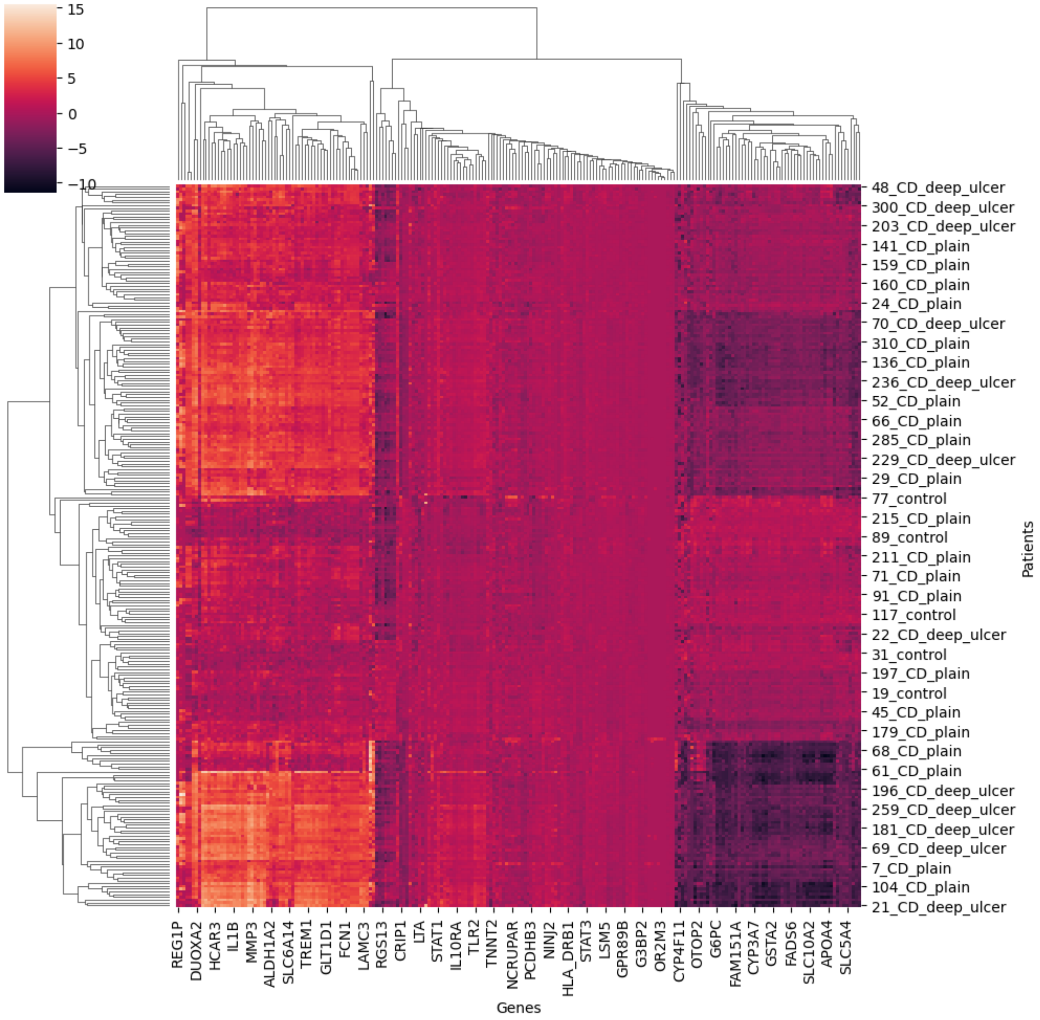

Figure 2: Hierarchical clustering heatmap including all 260 patients and all 221 genes.

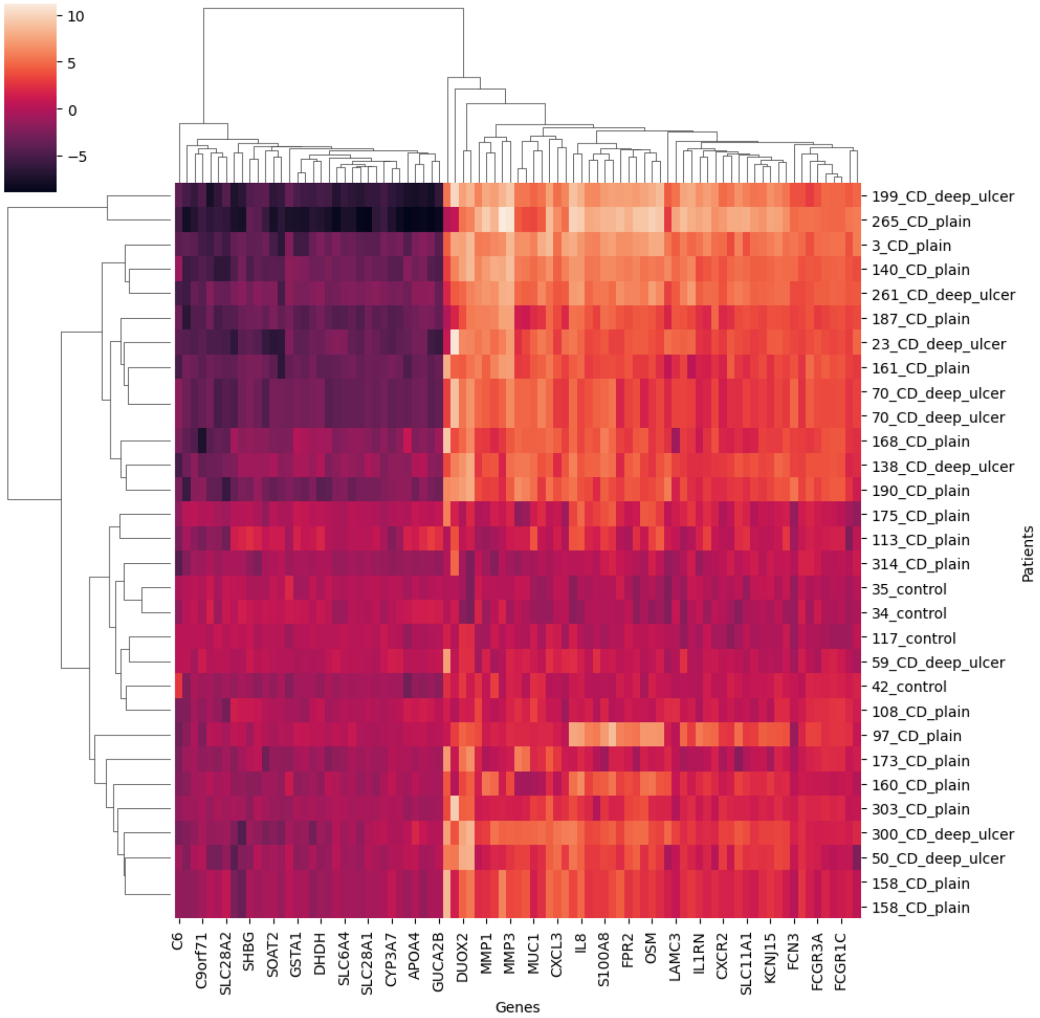

Figure 3: Hierarchical clustering heatmap including a random set of 30 patients and a selected subset of 87 genes informed by Welch's t test.

##### 3.2 Dimensionality reduction techniques

Autoencoders have been shown to effectively reduce dimensionality and noise in gene expression data. The structure of our autoencoder is detailed in Table 1. The training and validation loss curves for the final model are shown in Figure 4. We can see that the loss reduces and converges very quickly. The training and validation loss are almost identical from epoch 60 onwards which shows that the model is not overfitting to the data.

Table 1: Autoencoder architecture.

| Layer type | Output size | # Params |
|---|---|---|
| **Encoder** | | |
| Dense | 442 | 98124 |
| Batch Normalisation | 442 | 1768 |
| LeakyReLU | 442 | 0 |
| Dense | 221 | 97903 |
| Batch Normalisation | 221 | 884 |
| LeakyReLU | 221 | 0 |
| Dense | 32 | 7104 |
| **Decoder** | | |
| Dense | 221 | 7293 |
| Batch Normalisation | 221 | 884 |
| LeakyReLU | 221 | 0 |
| Dense | 442 | 98124 |
| Batch Normalisation | 442 | 1768 |
| LeakyReLU | 442 | 0 |
| Dense | 32 | 97903 |

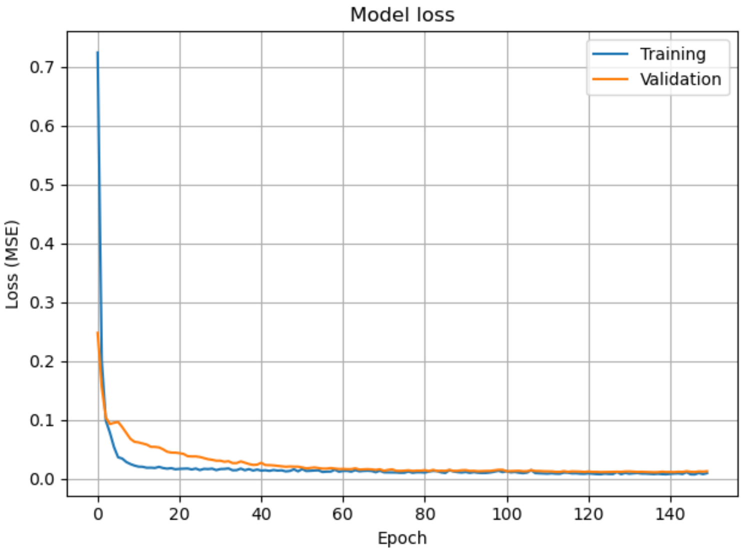

Figure 4: Autoencoder training and validation loss curves.

Figure 5 shows the proportion of variance explained by the number of principal components, when using PCA for dimensionality reduction. This figure was adapted from [90]. We reduce to 32 dimensions as this retains 91% of the variance [91]. We also reduce to 32 dimensions with the autoencoder for comparison purposes.

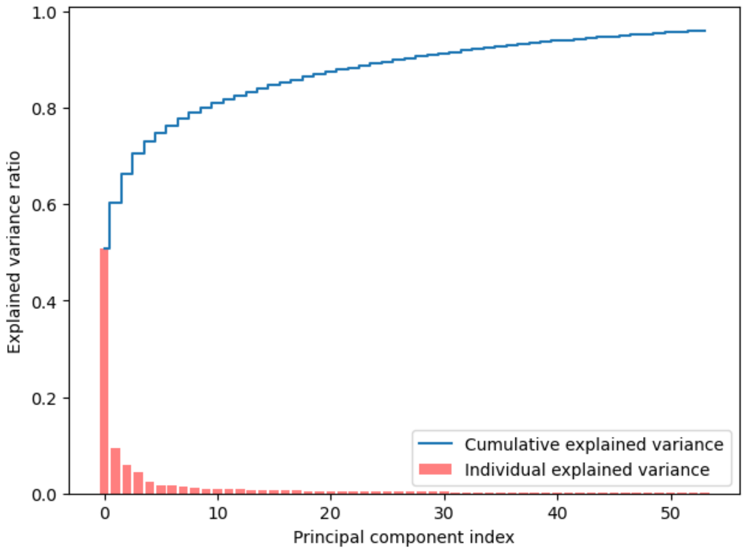

Figure 5: PCA scree plot, adapted from [90].

##### 3.3 Gaussian Mixture Model clustering

Below are some additional technical details and results relating to the training process of GMM clustering.

###### 3.3.1 PCA-based dimensionality reduction

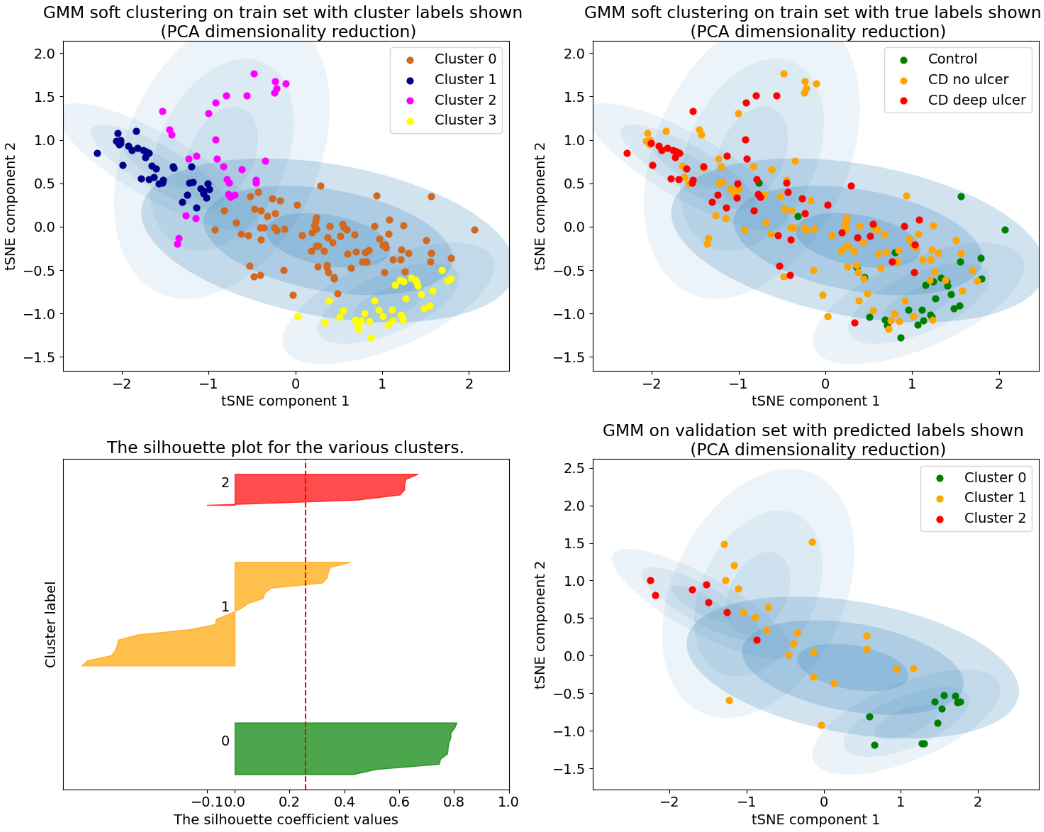

Figure 6: Sample of training results for GMM clustering after applying PCA and tSNE (perplexity=190) for dimensionality reduction. Results on the training set with four cluster labels shown (top left) and true labels shown (top right). Results on the validation set after post-processing shown on bottom right, alongside corresponding silhouette plot on bottom left. Clusters 0, 1 and 2 correspond to "control", "CD no ulcer" and "CD deep ulcer" clusters respectively.

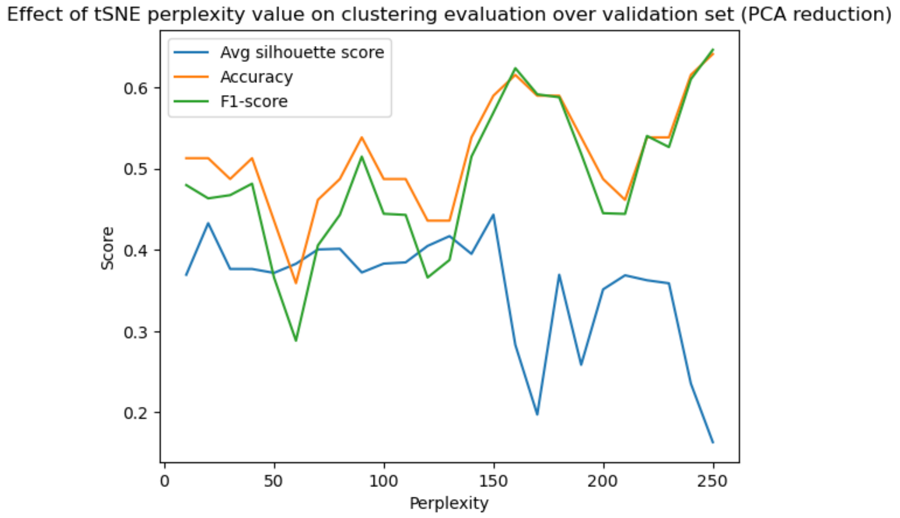

Figure 7: Effect of tSNE perplexity value on clustering and classification performance of GMM, with dimensionality reduced by PCA and tSNE, in terms of accuracy, F1-score and average silhouette score calculated on the validation set.

For two clustering models, PCA was used alongside tSNE for dimensionality reduction of the pre-processed RNA-Seq data. We first trained the GMM and KMeans models on the training set using various perplexity values for tSNE. For example, Figure 6 shows a sample of training results for the GMM when using a perplexity of 190 for tSNE, alongside PCA. We show the clustering on the training set with cluster labels (top left) and true labels (top right). After obtaining the initial clusters, we apply a post-processing step to classify disease phenotype, as explained in the main document. We show the post-processed clustering on the validation set with predicted labels on the bottom right, alongside the corresponding silhouette plot on the bottom left. In this example, the model somewhat distinguishes the classes but the clusters are not well-separated, as demonstrated in the silhouette plot. Most silhouette values are above 0.5 in clusters 0 and 2 but many have very negative values in cluster 1 so may be incorrectly assigned.

In Figure 7, we can see how dramatically the performance can vary, based on the perplexity chosen for tSNE. Perplexity determines the "effective number of neighbours" taken into account when calculating conditional probabilities that represent datapoint similarity. This can shift the focus between the local and global structure of the data. We tuned the perplexity by maximising the silhouette score (clustering quality), accuracy and F1-score (classification ability). For example, here we chose a perplexity of 150 as high scores were consistently achieved.

###### 3.3.2 Autoencoder-based dimensionality reduction

The same process was applied when the autoencoder was used for dimensionality reduction; please see the previous section for more details.

For the other two clustering models, our trained autoencoder was used alongside tSNE for dimensionality reduction of the pre-processed RNA-Seq data. We first trained the GMM and KMeans models on the training set using various perplexity values for the tSNE algorithm. For example, a sample of results for GMM clustering on the training and validation set are shown in Fig 8, when using a perplexity of 40 for tSNE.

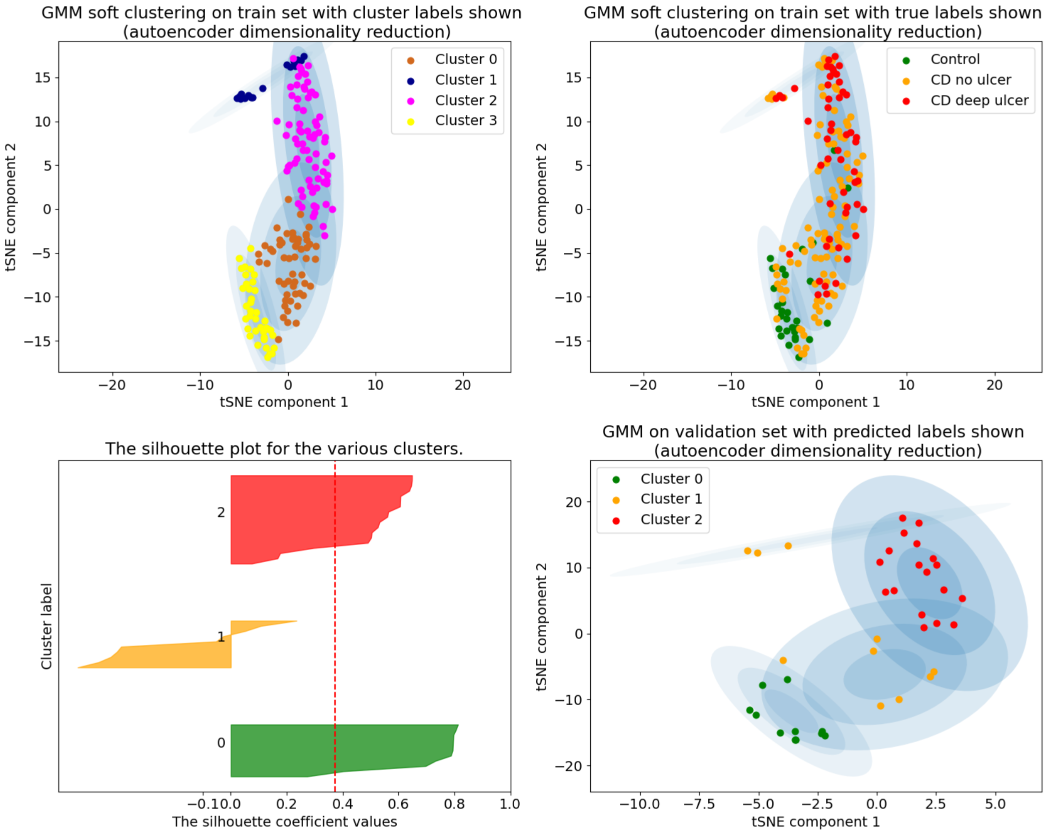

Figure 8: Sample of training results for GMM clustering after applying our autoencoder and tSNE (perplexity=40) for dimensionality reduction. Results on the training set with four cluster labels shown (top left) and true labels shown (top right). Results on the validation set after post-processing shown on bottom right, alongside corresponding silhouette plot on bottom left. Clusters 0, 1 and 2 correspond to "control", "CD no ulcer" and "CD deep ulcer" clusters respectively.

Fig 9 shows the effect of perplexity on classification and clustering performance as described above. The effect is markedly different to that of PCA (Fig 7) as the method of feature extraction for the autoencoder is much different to that of PCA. Here we chose a perplexity of 130 for deploying the model on the test set, as high validation scores are achieved.

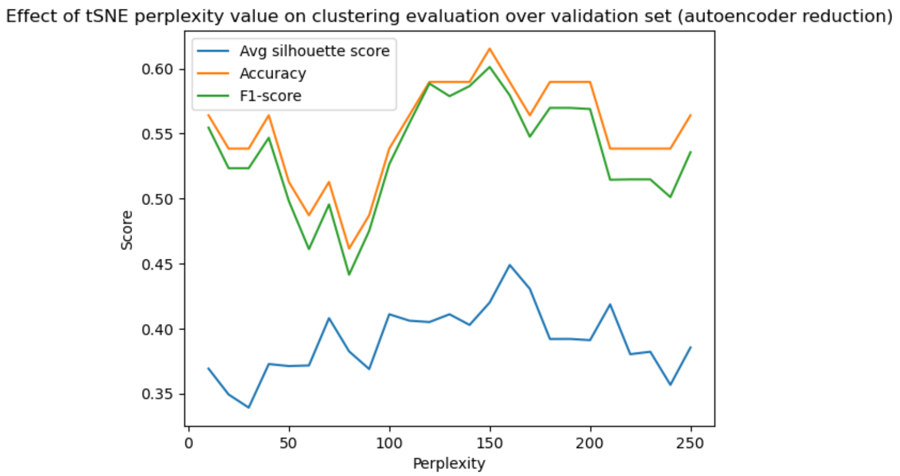

Figure 9: Effect of tSNE perplexity value on clustering and classification performance of GMM, with dimensionality reduced by our autoencoder and tSNE, in terms of accuracy, F1-score and average silhouette score calculated on the validation set.

##### 3.4 KMeans clustering

Figure 10 shows a visualisation of the final results for KMeans clustering on the test set.

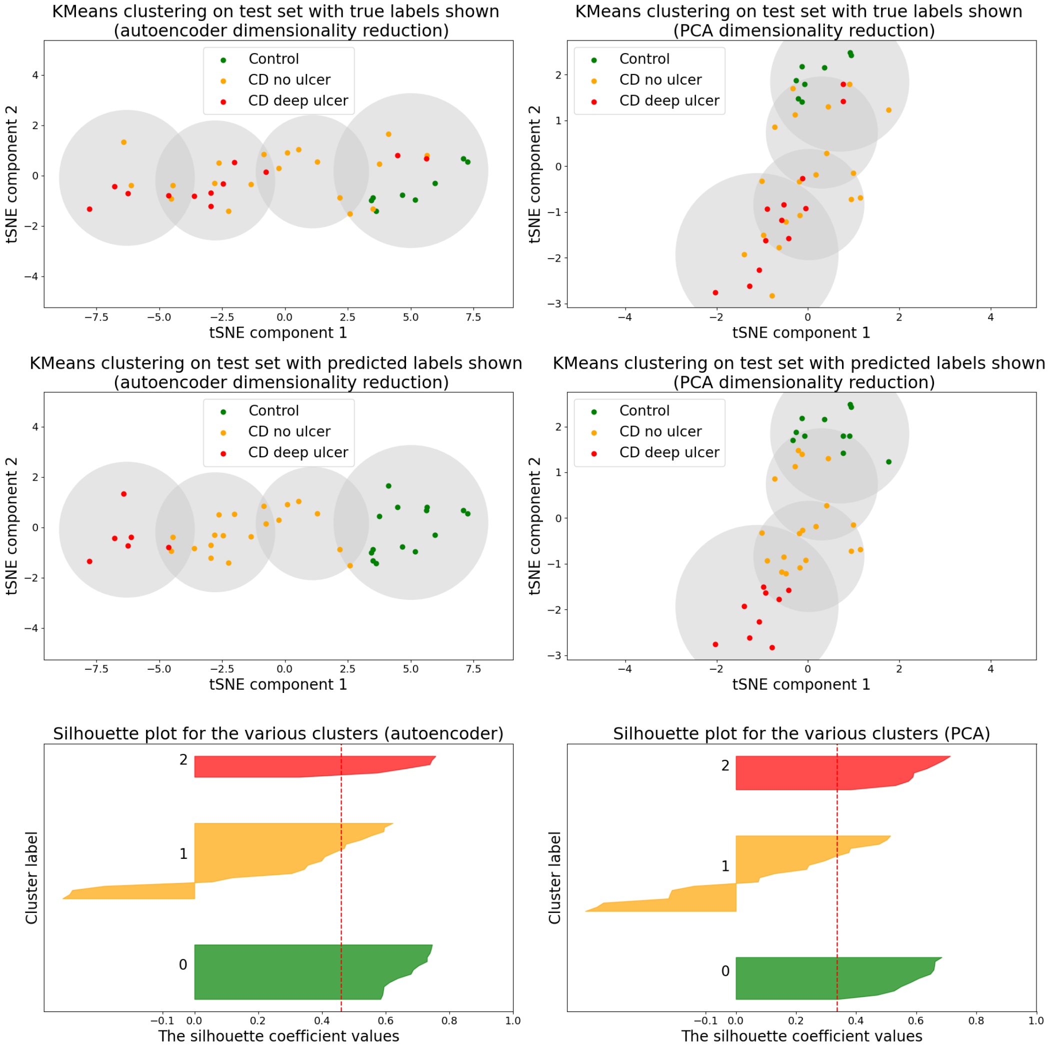

Figure 10: KMeans clustering model results after applying dimensionality reduction using autoencoder and tSNE (perplexity=90) (left) and PCA and tSNE (perplexity=160) (right). Deployed on the test set with true labels shown (top third) and predicted labels shown (middle third). Silhouette plots are shown for KMeans clusters after applying autoencoder-tSNE (left) and PCA-tSNE (right) methods, with clusters 0, 1 and 2 corresponding to “control”, “CD no ulcer” and “CD deep ulcer” respectively.

Below are some additional results relating to the training process of KMeans clustering. These correspond to those of the GMM in Section 3.3.

##### 3.4.1 PCA-based dimensionality reduction

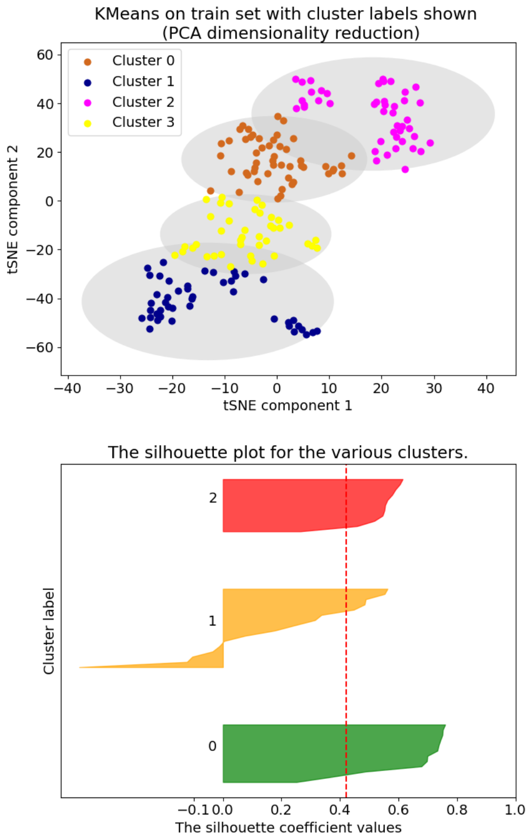

Figure 11: Sample of training results for KMeans clustering after applying PCA and tSNE (perplexity=190) for dimensionality reduction. Results on the training set with four cluster labels shown (top left) and true labels shown (top right). Results on the validation set after post-processing shown on bottom right, alongside corresponding silhouette plot on bottom left. Clusters 0, 1 and 2 correspond to “control”, “CD no ulcer” and “CD deep ulcer” clusters respectively.

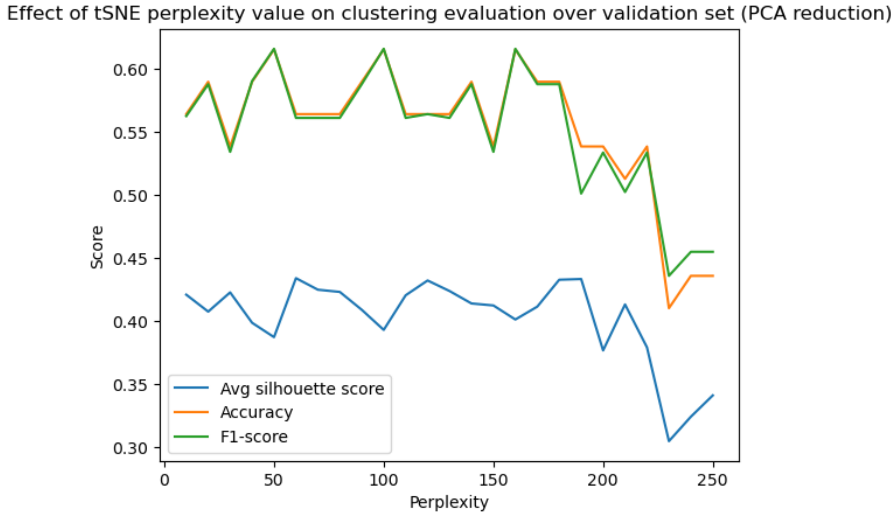

Figure 12: Effect of tSNE perplexity value on clustering and classification performance of KMeans, with dimensionality reduced by PCA and tSNE, in terms of accuracy, F1-score and average silhouette score calculated on the validation set.

##### 3.4.2 Autoencoder-based dimensionality reduction

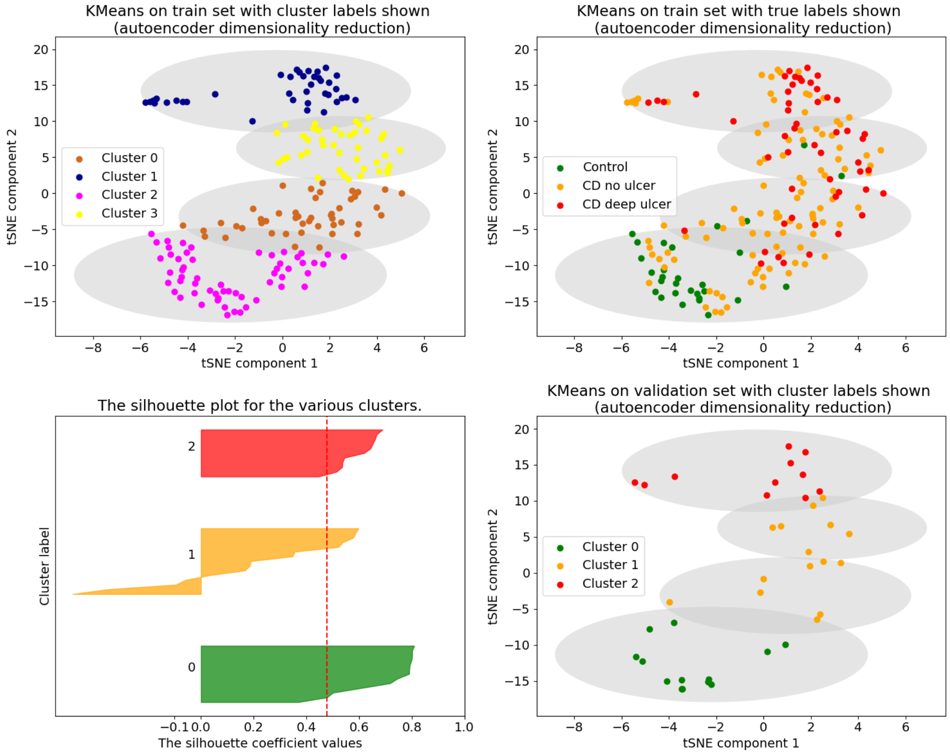

Figure 13: Sample of training results for KMeans clustering after applying our autoencoder and tSNE (perplexity=40) for dimensionality reduction. Results on the training set with four cluster labels shown (top left) and true labels shown (top right). Results on the validation set after post-processing shown on bottom right, alongside corresponding silhouette plot on bottom left. Clusters 0, 1 and 2 correspond to “control”, “CD no ulcer” and “CD deep ulcer” clusters respectively.

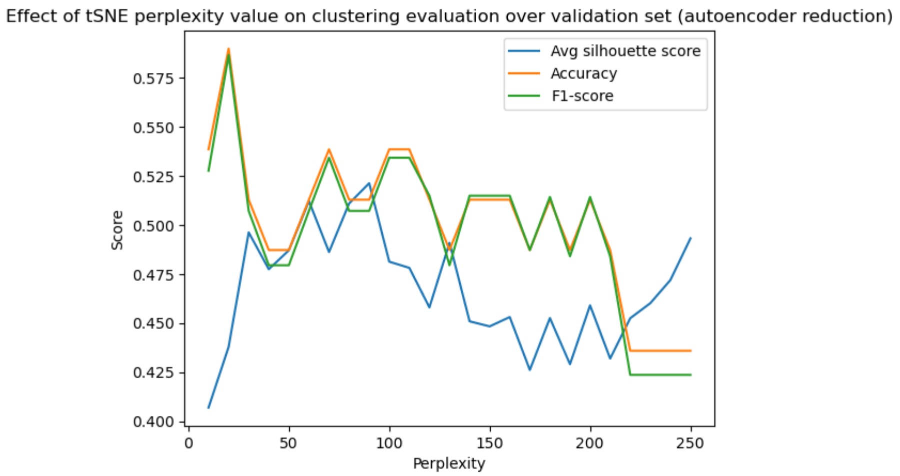

Figure 14: Effect of tSNE perplexity value on clustering and classification performance of KMeans, with dimensionality reduced by our autoencoder and tSNE, in terms of accuracy, F1-score and average silhouette score calculated on the validation set.

##### 3.5 Evaluation and comparison of KMeans and GMM

The final clustering and classification evaluation results are shown in Table 2. Despite the popularity of KMeans for RNA-Seq analysis in the literature, GMMs are demonstrably a much better method in the given context, achieving higher scores across the board for classification after post-processing, in the binary and multi-class settings. For example, accuracies and F1-scores are consistently above 90% in GMMs for binary classification but remain in the 80s with KMeans. The underlying distributions of the RNA-Seq data are Gaussian due to normalisation, making GMMs highly suitable. Because GMMs allow the covariance to be tuned, the mixture components become much better fitted to the data than the clusters in KMeans, which assumes spherical distributions. GMMs provides density estimation, which can be used to infer a more accurate phenotype in the post-processing step for classification. It also improves the visualisation of relationships between the expression profiles of different classes of patients. We can demonstrate the degree of association of each patient to each cluster, which in this case can indicate the severity of disease.

In general, the average silhouette scores are lower when using GMMs in comparison to KMeans, meaning the clusters are not as well-separated. This suggests that there may be a trade-off between classification and clustering quality. In the silhouette plots of Figure 10 and Figure 2 (main document), there are more negative silhouette scores in the “CD no ulcer” cluster for GMMs, showing greater uncertainty in this cluster. However, the average silhouette scores do not reach above 0.8 using either method. Classification performance is arguably more important than clustering quality in this context, since the phenotype classes are expected to be highly overlapping. When coupled with explainability techniques such as SHAP, an accurate classifier can lead to the identification of important risk genes and gene modules. In contrast to the current state-of-the-art, this process results in a verifiable and interpretable visual model, making it more applicable in clinical settings.

##### 3.6 Evaluation and comparison of dimensionality reduction methods

Table 2: Clustering and classification evaluation results for final GMM and KMeans models, using autoencoder and PCA dimensionality reduction methods. Results shown for binary classification (controls and all CD patients) and multi-class classification (control, CD no ulcer and CD deep ulcer).

| | | **Binary (control & CD)** | | **Multi-class (all labels)** | |
|---|---|---|---|---|---|
| | | Autoencoder | PCA | Autoencoder | PCA |
| **GMM** | Accuracy / % | 94.9 | 92.3 | 71.8 | 64.1 |
| | F1-Score / % | 96.7 | 94.9 | 71.5 | 62.6 |
| | Silh. score | 0.382 | 0.410 | 0.320 | 0.317 |
| **KMeans** | Accuracy / % | 84.6 | 82.1 | 64.1 | 59.0 |
| | F1-Score / % | 89.3 | 88.1 | 61.9 | 58.3 |
| | Silh. score | 0.556 | 0.409 | 0.469 | 0.334 |

We can see that using the autoencoder results in better performance overall compared to PCA when using GMMs. This is true for both binary and multi-class classification. For example, the accuracy and F1-score are 2.6% and 1.8% higher respectively when using the autoencoder for binary classification. For multi-class classification the difference is even larger at $\sim$8%. The clustering quality is more similar, with a negligible difference in silhouette scores in the multi-class setting and PCA achieving a higher average silhouette score by 0.028 in the binary setting.

When applying the KMeans algorithm, the autoencoder still leads to a better performance than PCA, although the differences are slightly less pronounced than with the GMMs. For example, the accuracy and F1-scores are 5.1% and 3.6% higher respectively for multi-class classification. When using KMeans, the clusters are not as well-fitted to the data due to the assumption of spherical distributions and lack of density estimation. This means the post-processing applied for classification is not as effective and performance is worse in comparison to the GMMs, regardless of dimensionality reduction technique.

PCA is a linear dimensionality reduction method that has been widely applied in RNA-Seq analysis [45]–[48]. However, it has been demonstrated that PCA can fail to detect important biological information, and has limitations when analysing small datasets or when effect size is small [46]. In complex contexts involving disease subtypes, autoencoders may be more suitable, due to their abilities in reducing noise and capturing linear and non-linear relationships. In future work, more advanced architectures can be explored, such as convolutional or variational autoencoders.

Regardless of dimensionality reduction method, the clusters overlap a lot by nature, as we are analysing subtypes of the same disease. This leads to similar performance in terms of clustering quality between methods. The GMM silhouette plots are nearly identical between PCA and autoencoder in Figure 2 (main document), but the autoencoder leads to slightly more negative scores in the "CD no ulcer" cluster and PCA leads to more negative scores in the "CD deep ulcer" cluster. In Figure 10 (KMeans), the cluster sizes are slightly different and PCA leads to a few more negative values in the "CD no ulcer" cluster. These slight differences are likely artefacts of the different methods of feature extraction employed by PCA and the autoencoder.

Overall, optimal performance is achieved by the GMM clustering model using the autoencoder and tSNE for dimensionality reduction. Coupled with SHAP for explainability, we can draw important insights about risk genes and gene modules which can be applied in clinical contexts.

#### 3.7 Cluster explainability using kernelSHAP with feature dependence

##### 3.7.1 Force plots

SHAP force plots show how features contributed to the model prediction for a given data instance. We show force plots for the "control" class and "CD deep ulcer" class for Patient 260, in Figures 15 and 16 respectively. The emboldened number shows the predicted probability of the patient being assigned to the given class. Genes in pink make a positive contribution towards this probability, and genes in blue a negative contribution.

When compared to the plots with feature independence (Fig 18 and 19) the contributions across the genes are slightly more equal in Figures 15 and 16. This may be because gene correlations are taken into account, so contributions are more evenly spread across correlated sets.

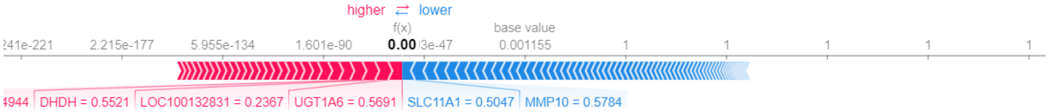

Figure 15: Force plot of Patient 260 for “control” class - dependent features.

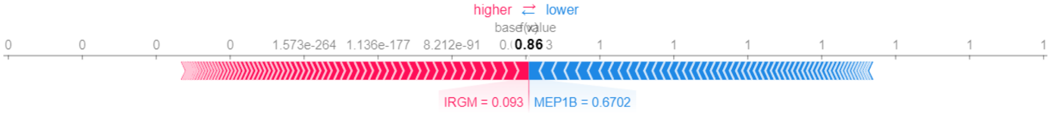

Figure 16: Force plot of Patient 260 for “CD deep ulcer” class - dependent features.

##### 3.7.2 Beeswarm plot

Figure 17 shows the beeswarm plot for CD deep ulcer with feature dependence included, which can be compared with the corresponding plot with feature independence in Figure 22. This shows the top 20 genes in terms of their influence on the “CD deep ulcer” class and the distributions of their impact on predictions. Each dot represents the SHAP value of that gene for a given patient, corresponding to the “CD deep ulcer” class. The colours also show how the expression value affects the impact on model predictions. The feature values are mixed fairly well across the distributions, likely due to the variants of each gene, which can encompass both causative and protective effects. Similarly to the summary plot, many established IBD genes are shown, such as NOD2, MEP1B, IRGM, JAK2, PTPN2, FOLH1 and the IL10s [92]–[98].

The genes IL10, IL10RA and IL10RB included in this plot are also known to be major risk factors. Interleukin-10 (IL-10) is an anti-inflammatory cytokine heavily involved in maintaining haemostasis in the intestinal tract. It does this by preventing pro-inflammatory cytokines like tumour necrosis factor (TNF) and IL-12 from being released. The IL-10 receptor contains $\alpha$ and $\beta$ subunits. IL-10RB codes for the $\beta$ subunits which are compatible with many different cytokines. When IL10 binds with IL10R, it activates Janus Kinase 1 (JAK1), Tyrosine kinase 2 (TYK2) and/or Signal Transducer and Activator of Transcription 3 (STAT3) signalling to prevent inflammation. Therefore, disruption of these genes often leads to the inflammatory symptoms of IBD [98]. When incorporating feature dependence, we demonstrably obtain more genes that affect the downstream processes involved in inflammation. This suggests that by taking gene correlations and dependencies into account, we can reach further to the root causes of the condition, leading to more effective therapeutic targets.

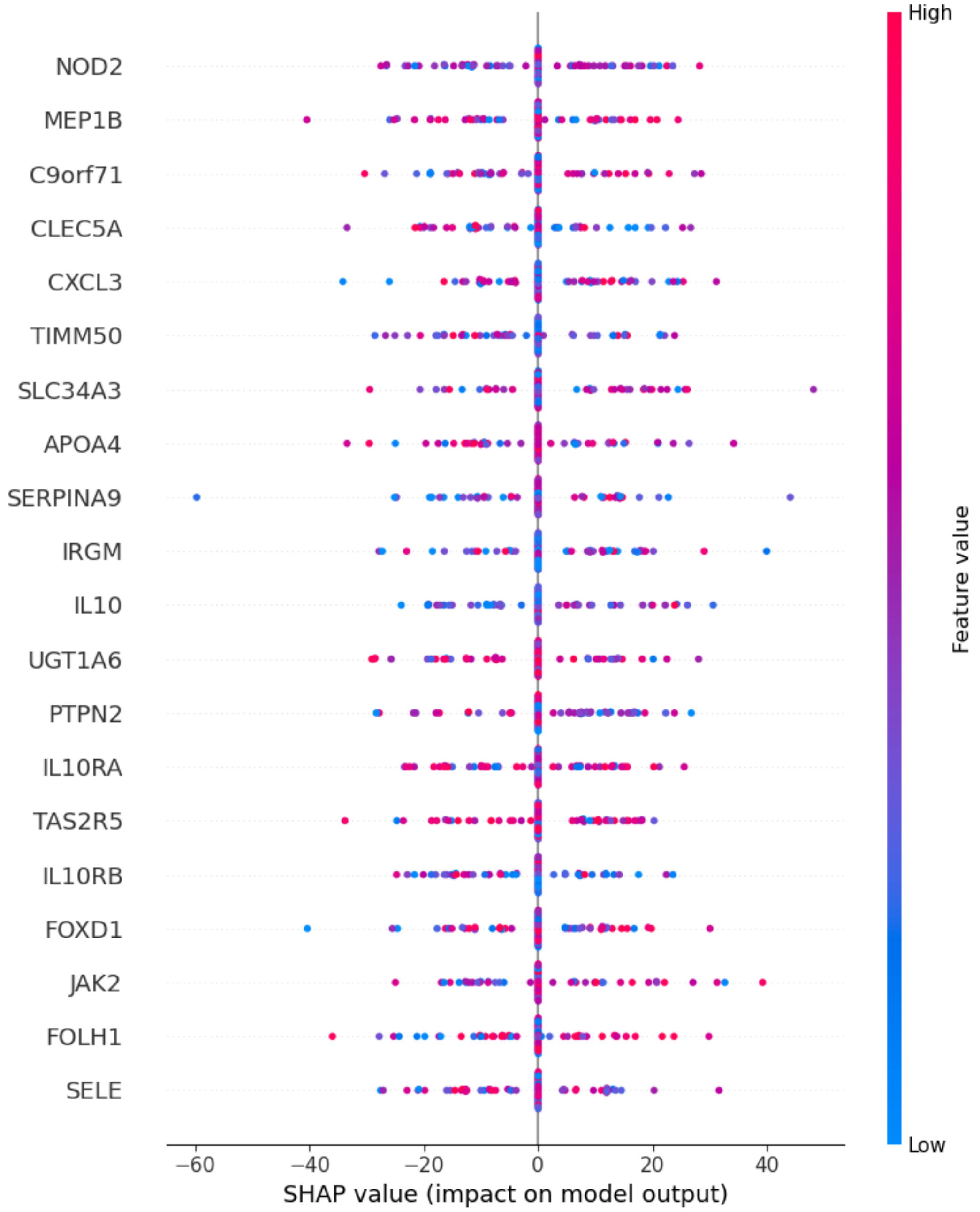

Figure 17: Beeswarm plot for “CD deep ulcer”, showing how expression value affects the impact of a gene on predictions for this class (feature dependence included). Genes are ranked by importance.

##### 3.8 Cluster explainability using original kernelSHAP

The following are results obtained when applying the original kernelSHAP algorithm to our GMM classifier. Features are assumed to act independently.

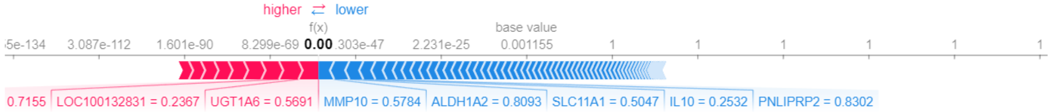

Figure 18: Force plot of Patient 260 for “control” class - independent features.

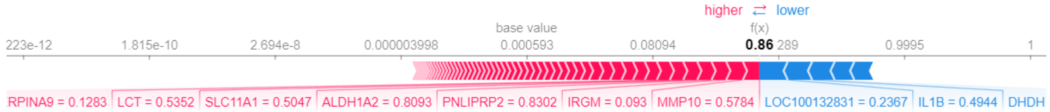

Figure 19: Force plot of Patient 260 for “CD deep ulcer” class - independent features.

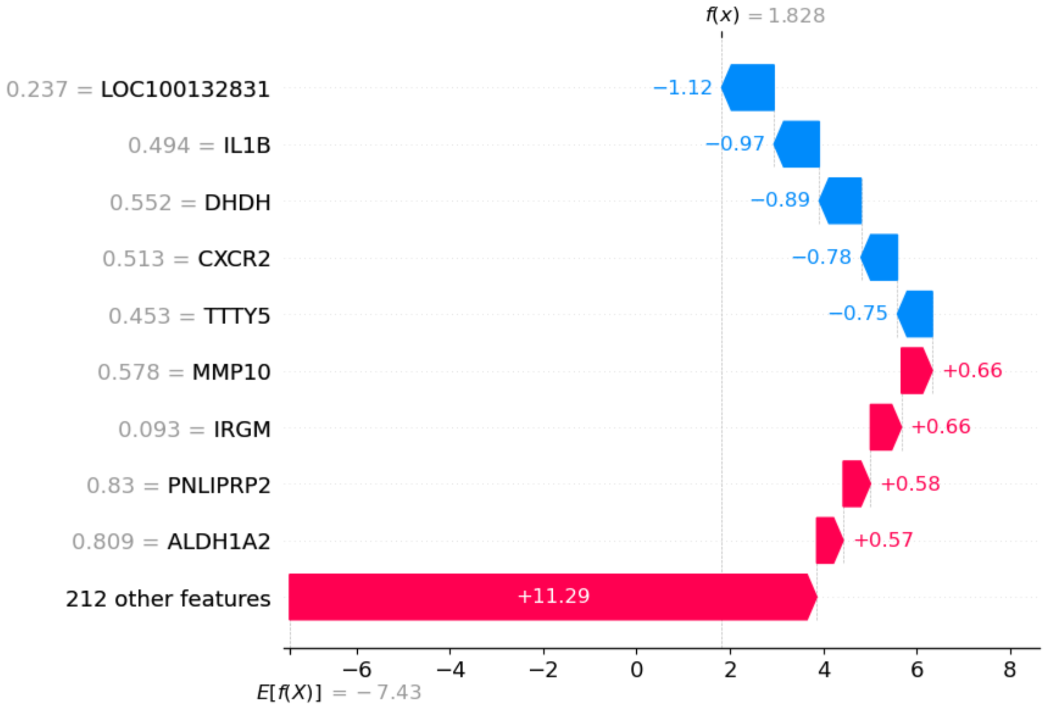

Figure 20: Waterfall plot of Patient 260 for “CD deep ulcer” class - independent features.

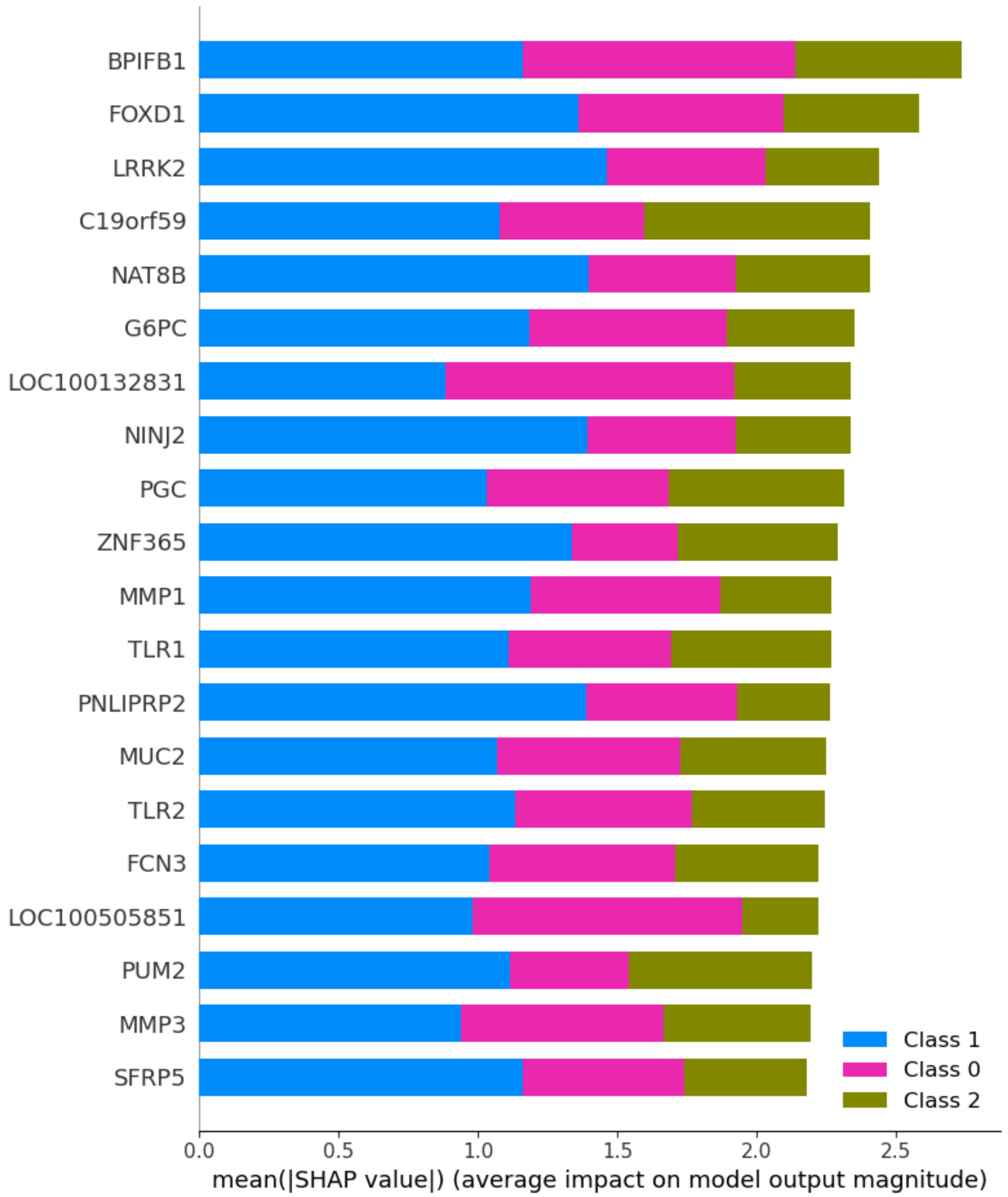

Figure 21: Summary plot showing top 20 genes in terms of their average impact on class predictions across all patients - independent features. Genes ranked by importance.

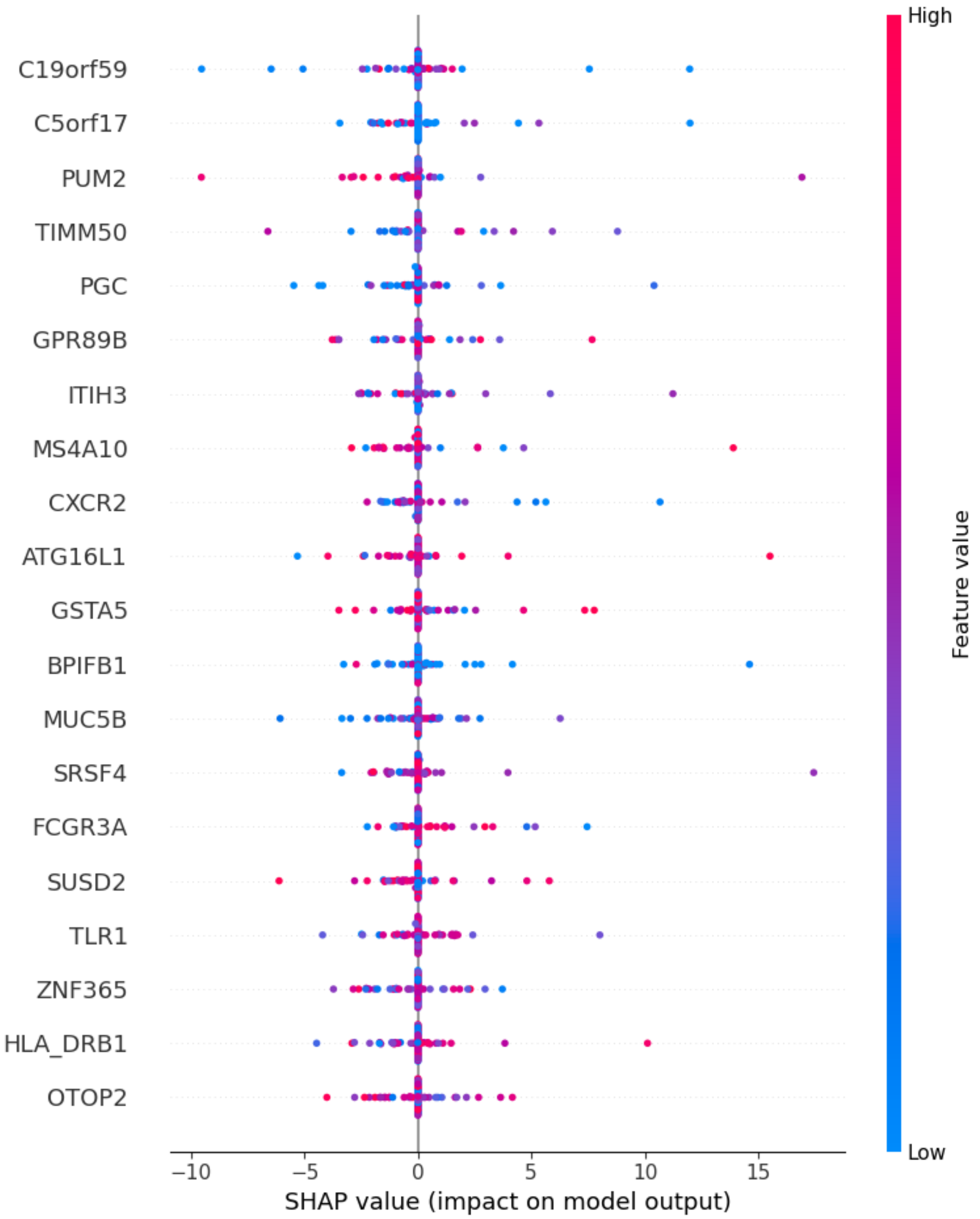

Figure 22: Beeswarm plot for "CD deep ulcer", showing how expression value affects the impact of a gene on predictions for this class (features independent). Genes are ranked by importance.

##### 3.9 Identification and characterisation of potential gene modules

The following are additional results obtained during the process of identifying and characterising potential gene modules. We include detailed results of Gene Ontology enrichment analysis.

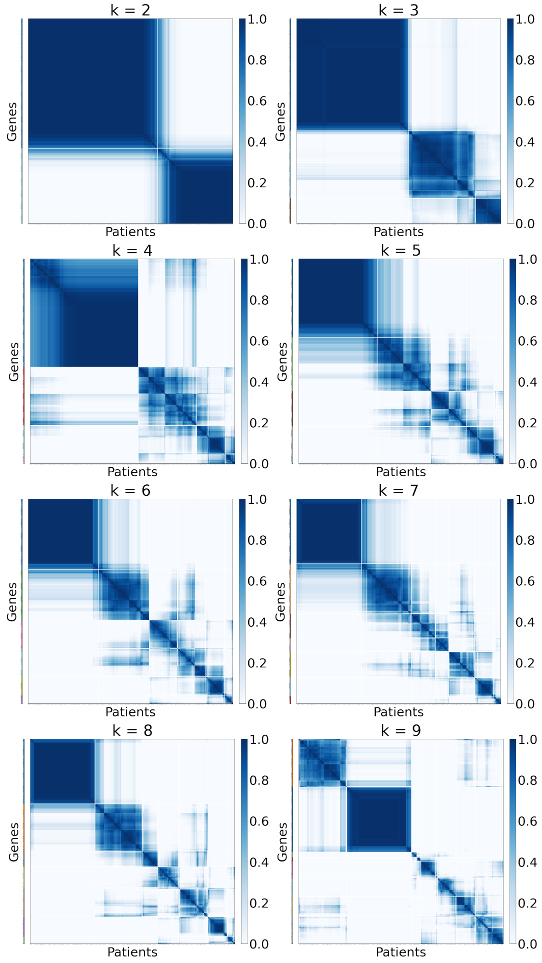

Figure 23: Results of WECR clustering [23], to identify "CD deep ulcer" gene modules, using various numbers of clusters $k$. Colour depth signifies extent of association.

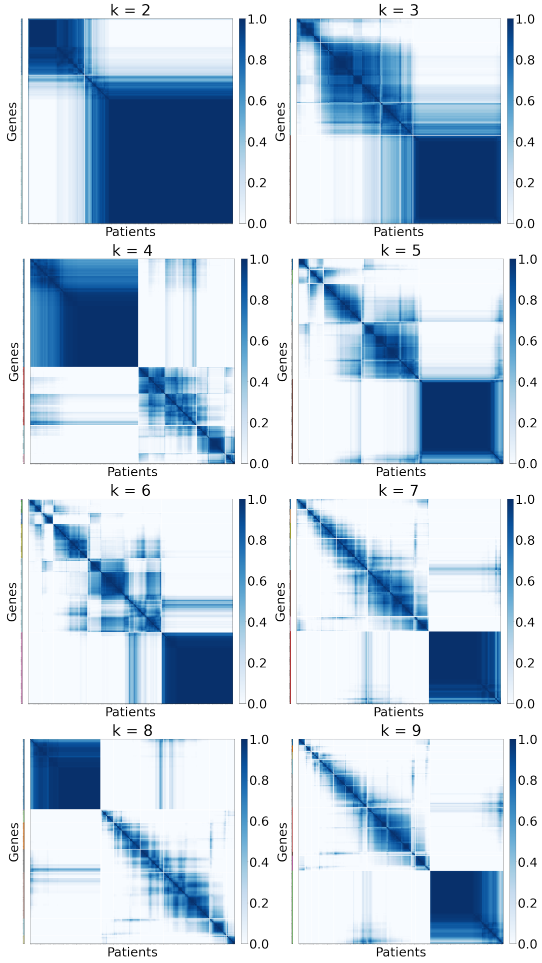

Figure 24: Results of WECR clustering [23], to identify "CD no ulcer" gene modules, using various numbers of clusters $k$. Colour depth signifies extent of association.

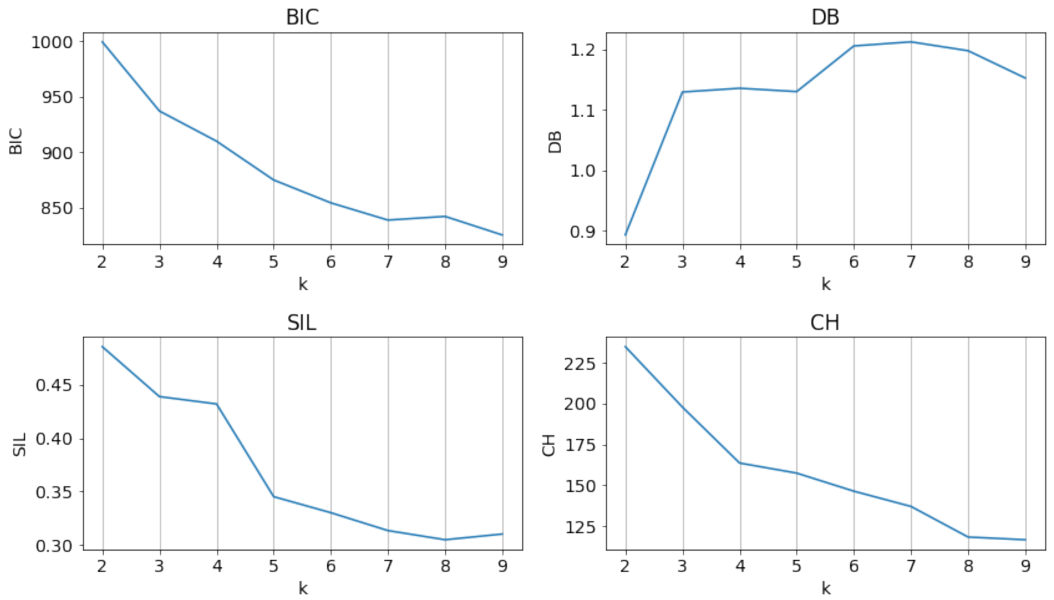

Figure 25: Plots for “CD deep ulcer” modules, to show how clustering evaluation metrics vary with the number of clusters $k$. Metrics: Bayesian Information Criterion (BIC), Davies-Bouldin (DB) Index, Silhouette Score (SIL) and Calinski-Harabasz (CH) Index.

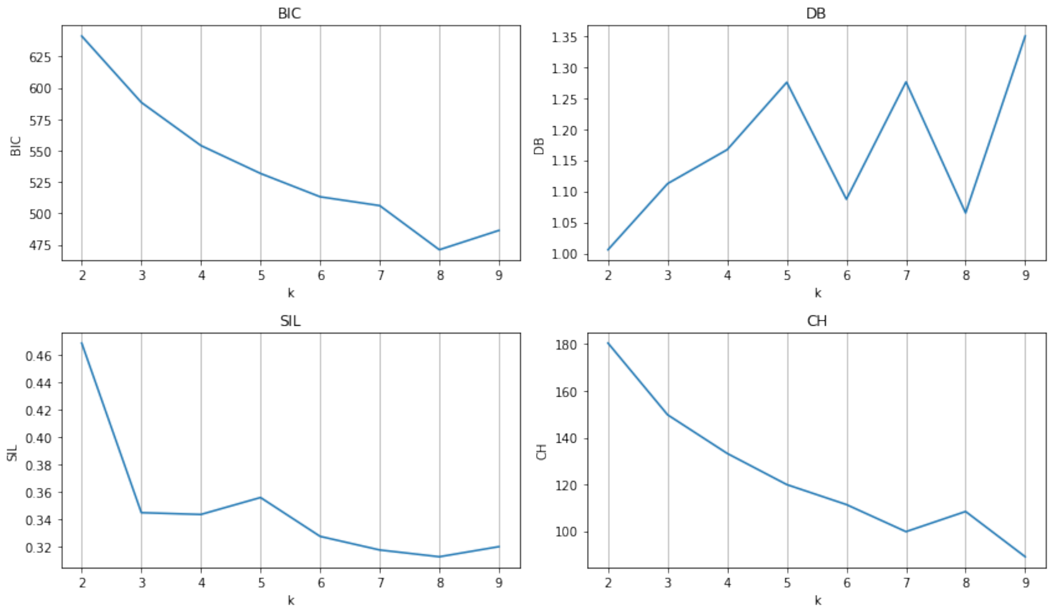

Figure 26: Plots for “CD no ulcer” modules, to show how clustering evaluation metrics vary with the number of clusters $k$. Metrics: Bayesian Information Criterion (BIC), Davies-Bouldin (DB) Index, Silhouette Score (SIL) and Calinski-Harabasz (CH) Index.

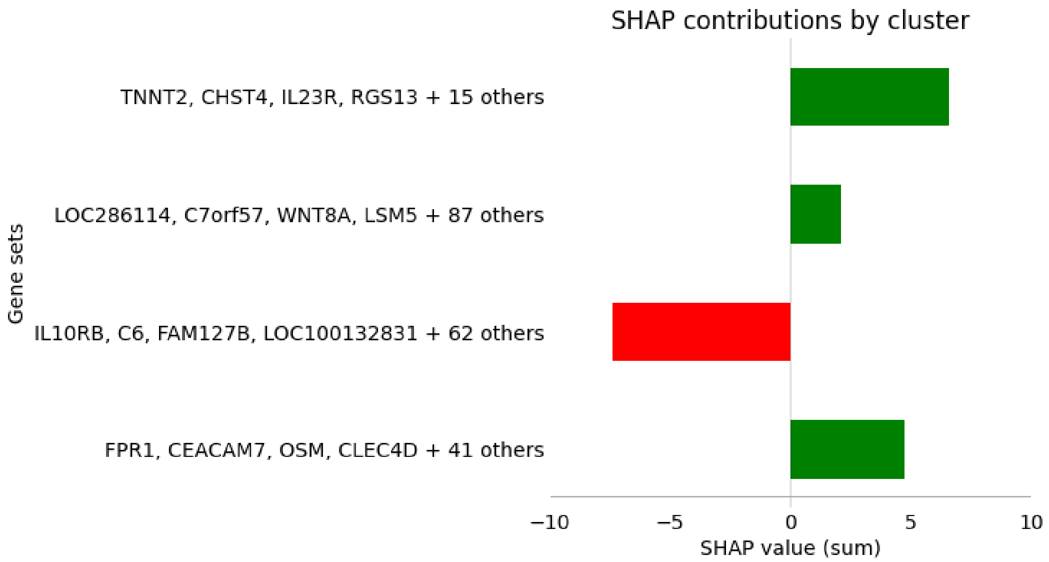

Figure 27: Final gene modules identified in association with CD symptoms without deep ulcer, alongside relative contributions determined using SHAP values.

##### 3.9.1 Gene Ontology enrichment analysis

Similarly to the 117-gene module analysed in the main document, GO analysis of the 63-gene module produced similar results, shown in Figure 28 and Table 6, with the most enriched processes relating to signalling pathways involved in the immune response to pathogens. However, here we also see the detection of slightly different molecules such as bacterial lipopeptides and signalling pathways involving TLR6 (Toll-like receptor 6) and TLR2 (Toll-like receptor 2). These can recognise a wide variety of pathogen-associated molecular patterns (PAMPs) such as lipoproteins and peptidoglycans [99], which extends recognition to Gram-positive bacteria. We also see processes relating to embryonic digestive tract development, extra-cellular matrix disassembly and tumour necrosis factor production, most of which are well-established in IBD [100], [101]. This suggests that the smaller module represents the additional and extended routes to disease symptoms.

The results for the 45-gene module are shown in Table 7 and Figure 29. We obtain similar results, such as neutrophil aggregation, with additional processes like autocrine signalling, immune response to fungus and use of the fc-gamma receptor signalling pathway. This suggests that the associations of “CD no ulcer” are more wide-ranging than “CD deep ulcer”; for example, fc-gamma receptors can recognise many different types of immunoglobulins [102].

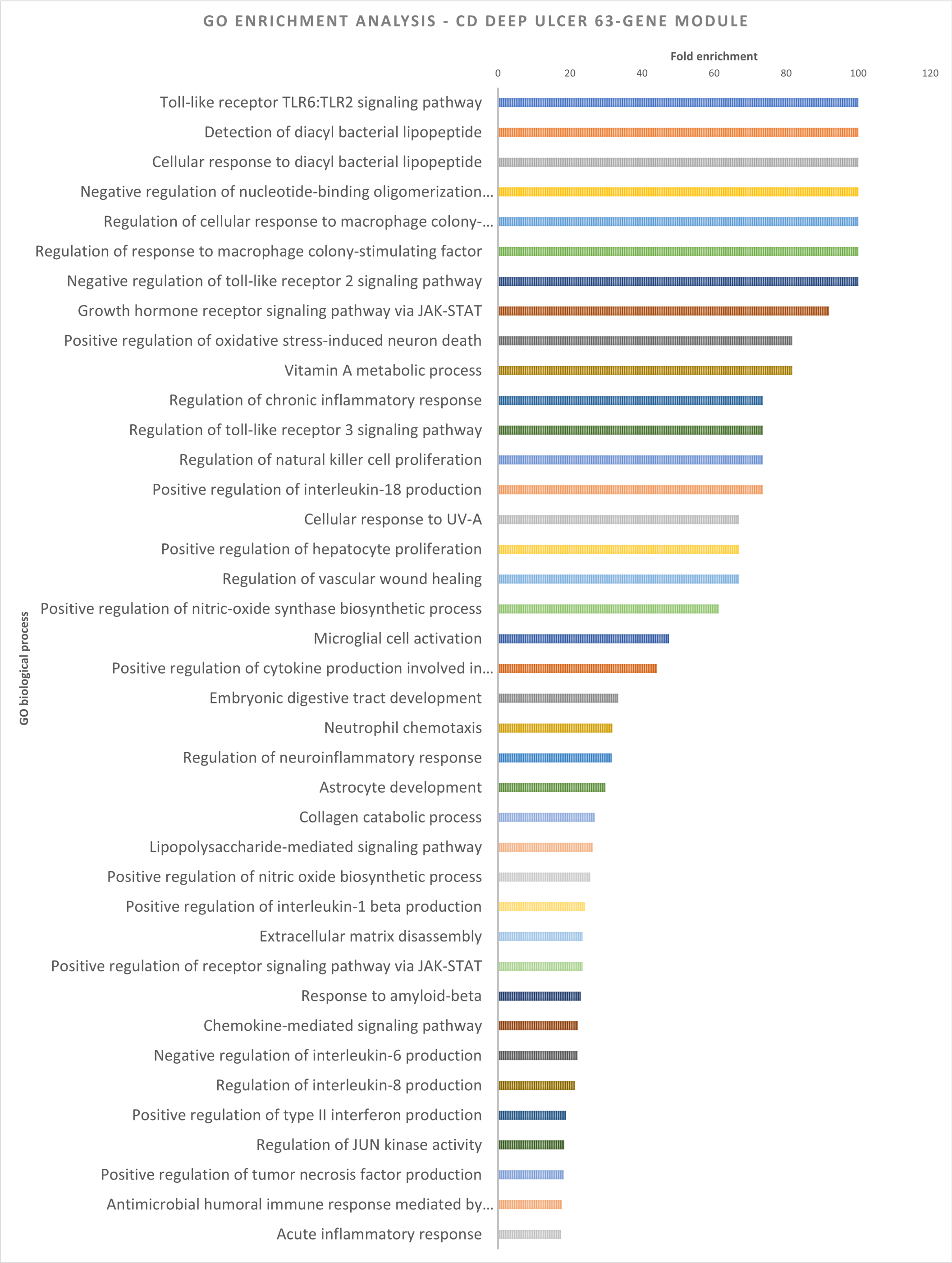

Figure 28: Gene Ontology enrichment analysis [6], [7], [103]: most enriched biological processes associated with 63-gene module identified for CD with deep ulcer. Full details given in Table 6.

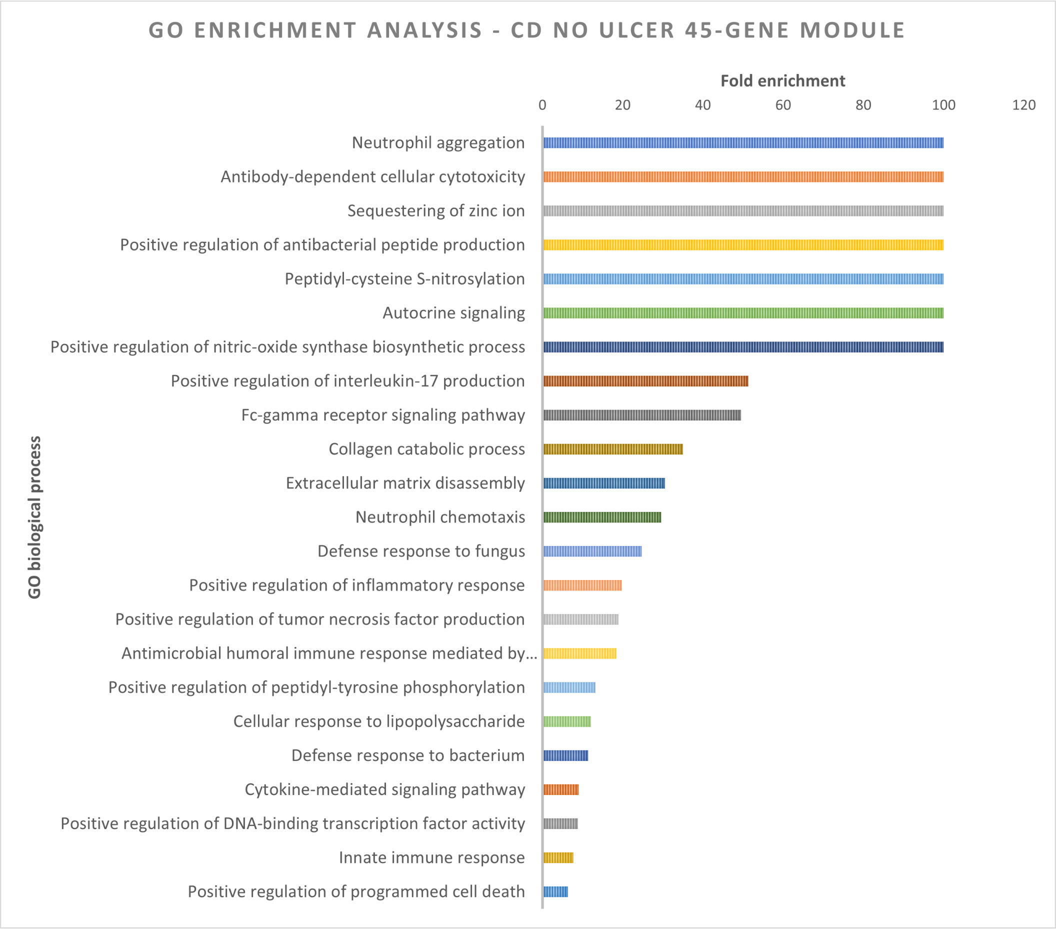

Figure 29: Gene Ontology enrichment analysis [6], [7], [103]: most enriched biological processes associated with 45-gene module identified in this work for CD without deep ulcer. Full details given in Table 7.

Table 3: CD deep ulcer gene module memberships, corresponding to Figure 5 (main document).

| Module A 117 genes | Module B 63 genes | Module C 10 genes | Module D 31 genes |
|---|---|---|---|
| IRGM | CXCL3 | TYK2 | C9orf71 |
| MIF | S100A12 | LCN2 | MEP1B |
| TIMM50 | FAM92A3 | CYP3A4 | C6 |
| WNT8A | IL10RB | APOB | HMGCS2 |
| GCM2 | LSM5 | KIAA1683 | SHBG |
| LOC283299 | LAMC3 | LOC100288778 | G3BP2 |
| C7orf57 | GRAMD1A | GUSBP11 | CYP4F11 |
| CEACAM7 | HLA_B | TLR1 | FOXD1 |
| LOC339166 | IL8 | ICAM5 | AGXT2 |
| REG1P | RNF24 | LTA | PNLIPRP2 |
| LOC286114 | FXYD5 | | APOA1 |
| LOC100505851 | TNFAIP2 | | GSTA1 |
| PPP1R17 | MMP7 | | TAS2R5 |
| NCRUPAR | FAM127B | | PTPN21 |
| PGC | FCN3 | | CUBN |
| table continues | | | |

| continue table | | | |
|---|---|---|---|
| Module A | Module B | Module C | Module D |
| SELE | STAT3 | | TCF7L2 |
| CRP | DUOX2 | | G6PC |
| PROK2 | MUC5B | | DHDH |
| FPR1 | TNNT2 | | CDHR1 |
| LOC147646 | FCGR3B | | ATG16L1 |
| HCAR3 | JAK2 | | SLC5A12 |
| NAT8 | PTPN2 | | SLC34A3 |
| FPR2 | FCGR1B | | NAT8B |
| CXCL9 | KCNJ15 | | GSTA5 |
| TREM1 | FCGR3A | | SLC10A2 |
| CLEC5A | LRRK2 | | SLC28A1 |
| BPIFB1 | TNFAIP3 | | APOA4 |
| FRMD1 | SERPINA9 | | FABP6 |
| OTOP2 | CXCL5 | | SLC13A1 |
| SUSD2 | TCN1 | | GSTA2 |
| OSM | HCAR2 | | SFRP5 |
| IL10RA | ZNF365 | | |
| LCT | DACT3 | | |
| FOLH1B | CXCR1 | | |
| NOD2 | TNFAIP6 | | |
| FCGR1A | ALDH1A2 | | |
| FCRL3 | HLA_DRB5 | | |
| EFNB1 | FOLH1 | | |
| HSD11B1 | S100A8 | | |
| APOC3 | TLR2 | | |
| CSF3 | TLR6 | | |
| XPNPEP2 | SAA2 | | |
| CHI3L1 | GLT1D1 | | |
| CXCR2 | WLS | | |
| REG1A | FCRL4 | | |
| IL1RN | SLC11A1 | | |
| LOC392364 | MMP1 | | |
| IL10 | PTPN22 | | |
| CXCL6 | MMP3 | | |
| FCN1 | FCGR1C | | |
| LYPD1 | FLJ35424 | | |
| SLC6A4 | SRRD | | |
| C16orf78 | LRAT | | |
| RGS13 | SHISA2 | | |
| C19orf59 | SLC23A3 | | |
| LEPREL1 | HLA_DRB1 | | |
| PCDHB3 | CXCL11 | | |
| CPO | TNF | | |
| TCF4 | FADS6 | | |
| PUM2 | AQP9 | | |
| SLC28A2 | ACYP2 | | |
| SLC22A4 | FAM151A | | |
| FDCSP | FGF11 | | |
| CHST4 | | | |
| LATS2 | | | |
| DUOXA2 | | | |
| HSPA7 | | | |
| LOC100506115 | | | |
| FHIT | | | |
| table continues | | | |

| continue table | | | |
|---|---|---|---|
| Module A | Module B | Module C | Module D |
| CYP3A7 | | | |
| EGFL6 | | | |
| MGAM | | | |
| SLC22A5 | | | |
| CLVS1 | | | |
| OR2M3 | | | |
| LYPLAL1 | | | |
| IL1B | | | |
| CRIP1 | | | |
| IL12B | | | |
| MUC2 | | | |
| TTTY5 | | | |
| CLDN8 | | | |
| SLC6A14 | | | |
| AADAC | | | |
| STAT1 | | | |
| SLC5A4 | | | |
| C5orf17 | | | |
| MUC1 | | | |
| TM4SF19 | | | |
| SAA1 | | | |
| GUCA2B | | | |
| LOC100132831 | | | |
| SRSF4 | | | |
| FANCF | | | |
| ITIH3 | | | |
| CNTFR | | | |
| IL23R | | | |
| MS4A10 | | | |
| NINJ2 | | | |
| MMP10 | | | |
| CLEC4D | | | |
| SLC5A11 | | | |
| OTOP3 | | | |
| CYP4F2 | | | |
| TLR4 | | | |
| FMO1 | | | |
| CNR1 | | | |
| ABCC2 | | | |
| CD300E | | | |
| S100A9 | | | |
| TM4SF4 | | | |
| UGT1A6 | | | |
| KLHL4 | | | |
| GPR89B | | | |
| TPMT | | | |
| SOAT2 | | | |
| DLG5 | | | |

Table 4: CD no ulcer gene module memberships, corresponding to Figure 27.

| Module A<br>19 genes | Module B<br>91 genes | Module C<br>66 genes | Module D<br>45 genes |
|---|---|---|---|
| TNNT2 | LOC286114 | IL10RB | FPR1 |
| CHST4 | C7orf57 | C6 | CEACAM7 |
| IL23R | WNT8A | FAM127B | OSM |
| RGS13 | LSM5 | LOC100132831 | CLEC4D |
| NAT8B | BPIFB1 | TCF7L2 | NOD2 |
| DACT3 | LOC147646 | FANCF | FCGR3A |
| CXCL6 | IL10 | CRIP1 | NINJ2 |
| AGXT2 | TM4SF19 | LOC283299 | PGC |
| APOC3 | IRGM | FABP6 | TCN1 |
| SLC5A12 | C16orf78 | ZNF365 | CXCL9 |
| TLR4 | NCRUPAR | FLJ35424 | MMP10 |
| SLC22A4 | CLEC5A | SLC22A5 | GRAMD1A |
| LCT | C5orf17 | SERPINA9 | LRRK2 |
| ABCC2 | LOC100505851 | GSTA1 | FCGR1B |
| LTA | OR2M3 | SLC13A1 | LOC100506115 |
| TLR1 | SLC11A1 | FCRL4 | S100A9 |
| HLA_DRB1 | LEPREL1 | HMGCS2 | PROK2 |
| FAM151A | CRP | MS4A10 | SLC6A14 |
| ATG16L1 | LAMC3 | GUCA2B | HLA_DRB5 |
| | CHI3L1 | SLC10A2 | HCAR2 |
| | LOC339166 | TYK2 | IL1RN |
| | PTPN2 | ACYP2 | IL10RA |
| | DUOX2 | CNR1 | MMP3 |
| | HSPA7 | TCF4 | SAA2 |
| | CXCR2 | CNTFR | LCN2 |
| | PPP1R17 | MGAM | ICAM5 |
| | REG1P | LYPLAL1 | C19orf59 |
| | MMP1 | CYP4F11 | FCGR1A |
| | PUM2 | SHBG | CSF3 |
| | FCRL3 | TLR6 | FOLH1B |
| | CLDN8 | TAS2R5 | MUC5B |
| | HCAR3 | PTPN21 | EGFL6 |
| | CYP3A7 | LRAT | CXCL5 |
| | MUC2 | UGT1A6 | ALDH1A2 |
| | AADAC | GSTA2 | FCGR3B |
| | FHIT | SLC6A4 | JAK2 |
| | SHISA2 | SRSF4 | STAT1 |
| | SELE | WLS | LATS2 |
| | EFNB1 | FRMD1 | TNFAIP2 |
| | RNF24 | C9orf71 | MMP7 |
| | PCDHB3 | FPR2 | TREM1 |
| | FAM92A3 | OTOP3 | CD300E |
| | CXCR1 | G3BP2 | S100A8 |
| | FCN1 | FMO1 | FCN3 |
| | CLVS1 | CXCL3 | AQP9 |
| | TIMM50 | APOA1 | |
| | G6PC | ITIH3 | |
| | FXYD5 | KLHL4 | |
| | DHDH | HLA_B | |
| | FCGR1C | IL8 | |
| | STAT3 | FADS6 | |
| table continues | | | |

| Module A | Module B | Module C | Module D |
|---|---|---|---|
| | SLC34A3 | MEP1B | |
| | APOB | APOA4 | |
| | TNFAIP3 | CUBN | |
| | TPMT | CXCL11 | |
| | LOC100288778 | KIAA1683 | |
| | CYP3A4 | DLG5 | |
| | TTTY5 | SFRP5 | |
| | SLC5A4 | REG1A | |
| | FDCSP | NAT8 | |
| | GSTA5 | SLC28A1 | |
| | S100A12 | CYP4F2 | |
| | LOC392364 | SLC23A3 | |
| | SRRD | SOAT2 | |
| | KCNJ15 | FGF11 | |
| | DUOXA2 | GUSBP11 | |
| | TNFAIP6 | | |
| | LYPD1 | | |
| | TNF | | |
| | PTPN22 | | |
| | TLR2 | | |
| | SLC28A2 | | |
| | TM4SF4 | | |
| | OTOP2 | | |
| | SUSD2 | | |
| | GLT1D1 | | |
| | IL12B | | |
| | CDHR1 | | |
| | MIF | | |
| | MUC1 | | |
| | GCM2 | | |
| | SLC5A11 | | |
| | IL1B | | |
| | FOXD1 | | |
| | FOLH1 | | |
| | HSD11B1 | | |
| | XPNPEP2 | | |
| | CPO | | |
| | PNLIPRP2 | | |
| | SAA1 | | |
| | GPR89B | | |

Table 5: GO enrichment analysis results for CD deep ulcer module A (117-gene module).

| GO biological process | Homo sapiens (REF) | Gene module | Fold Enrich. | Raw P value | FDR |
|---|---|---|---|---|---|
| (R)-carnitine transmembrane transport | 3 | 2 | >100 | 2.67E-04 | 3.02E-02 |
| Negative regulation of interleukin-18 production | 4 | 2 | 95.32 | 3.99E-04 | 3.99E-02 |
| Positive regulation of T-helper 17 cell lineage commitment | 4 | 2 | 95.32 | 3.99E-04 | 3.97E-02 |

Continued on next page

Table 5: GO enrichment analysis results for CD deep ulcer module A (117-gene module). (Continued)

| | | | | | |
|---|---|---|---|---|---|
| Disaccharide catabolic process | 4 | 2 | 95.32 | 3.99E-04 | 3.94E-02 |
| Regulation of myeloid dendritic cell activation | 5 | 2 | 76.26 | 5.57E-04 | 4.91E-02 |
| Positive regulation of antibacterial peptide production | 5 | 2 | 76.26 | 5.57E-04 | 4.88E-02 |
| Nucleotide-binding oligomerization domain containing 2 signalling pathway | 9 | 3 | 63.55 | 2.94E-05 | 5.39E-03 |
| Negative regulation of myeloid cell apoptotic process | 18 | 4 | 42.36 | 4.77E-06 | 1.31E-03 |
| Positive regulation of granulocyte macrophage colony-stimulating factor production | 15 | 3 | 38.13 | 1.06E-04 | 1.50E-02 |
| Maintenance of gastrointestinal epithelium | 22 | 4 | 34.66 | 9.59E-06 | 2.17E-03 |
| Positive regulation of nitric-oxide synthase biosynthetic process | 18 | 3 | 31.77 | 1.72E-04 | 2.16E-02 |
| Positive regulation of T-helper 1 type immune response | 18 | 3 | 31.77 | 1.72E-04 | 2.14E-02 |
| Positive regulation of acute inflammatory response | 27 | 4 | 28.24 | 1.98E-05 | 3.96E-03 |
| Positive regulation of icosanoid secretion | 21 | 3 | 27.23 | 2.58E-04 | 2.99E-02 |
| Positive regulation of interleukin-17 production | 28 | 4 | 27.23 | 2.25E-05 | 4.39E-03 |
| Regulation of heterotypic cell-cell adhesion | 24 | 3 | 23.83 | 3.69E-04 | 3.79E-02 |
| Negative regulation of lipid catabolic process | 26 | 3 | 22 | 4.57E-04 | 4.38E-02 |
| Positive regulation of macrophage activation | 28 | 3 | 20.43 | 5.58E-04 | 4.87E-02 |
| Negative regulation of inflammatory response to antigenic stimulus | 28 | 3 | 20.43 | 5.58E-04 | 4.84E-02 |
| Inflammatory response to antigenic stimulus | 40 | 4 | 19.06 | 8.10E-05 | 1.23E-02 |
| Acute-phase response | 42 | 4 | 18.16 | 9.66E-05 | 1.42E-02 |
| Negative regulation of fatty acid metabolic process | 42 | 4 | 18.16 | 9.66E-05 | 1.41E-02 |
| Negative regulation of type II interferon production | 43 | 4 | 17.73 | 1.05E-04 | 1.51E-02 |
| Xenobiotic transport | 43 | 4 | 17.73 | 1.05E-04 | 1.49E-02 |

Continued on next page

Table 5: GO enrichment analysis results for CD deep ulcer module A (117-gene module). (Continued)

| | | | | | |
|---|---|---|---|---|---|
| Positive regulation of interleukin-12 production | 44 | 4 | 17.33 | 1.14E-04 | 1.55E-02 |
| Neutrophil chemotaxis | 81 | 7 | 16.47 | 3.88E-07 | 2.16E-04 |
| Regulation of defence response to virus by host | 47 | 4 | 16.22 | 1.45E-04 | 1.95E-02 |
| Positive regulation of receptor signalling pathway via JAK-STAT | 47 | 4 | 16.22 | 1.45E-04 | 1.94E-02 |
| Positive regulation of interleukin-8 production | 65 | 5 | 14.66 | 3.26E-05 | 5.84E-03 |
| Regulation of viral-induced cytoplasmic pattern recognition receptor signalling pathway | 57 | 4 | 13.38 | 2.91E-04 | 3.15E-02 |
| Positive regulation of phagocytosis | 75 | 5 | 12.71 | 6.21E-05 | 9.79E-03 |
| Regulation of interleukin-10 production | 61 | 4 | 12.5 | 3.72E-04 | 3.77E-02 |
| Xenobiotic metabolic process | 125 | 8 | 12.2 | 4.83E-07 | 2.51E-04 |
| Regulation of chemokine production | 95 | 6 | 12.04 | 1.49E-05 | 3.23E-03 |
| Regulation of B cell proliferation | 66 | 4 | 11.55 | 4.93E-04 | 4.64E-02 |
| Positive regulation of B cell activation | 85 | 5 | 11.21 | 1.09E-04 | 1.50E-02 |
| Positive regulation of reactive oxygen species metabolic process | 69 | 4 | 11.05 | 5.79E-04 | 4.96E-02 |
| Positive regulation of NIK/NF-$\kappa$B signalling | 69 | 4 | 11.05 | 5.79E-04 | 4.93E-02 |
| Defence response to Gram-negative bacterium | 94 | 5 | 10.14 | 1.71E-04 | 2.18E-02 |
| Antimicrobial humoral immune response mediated by antimicrobial peptide | 104 | 5 | 9.17 | 2.68E-04 | 2.98E-02 |
| Positive regulation of lymphocyte proliferation | 146 | 7 | 9.14 | 1.57E-05 | 3.31E-03 |
| Cellular response to lipopolysaccharide | 199 | 9 | 8.62 | 1.46E-06 | 5.41E-04 |
| Regulation of cytokine production involved in immune response | 121 | 5 | 7.88 | 5.22E-04 | 4.73E-02 |
| Positive regulation of peptidyl-tyrosine phosphorylation | 181 | 7 | 7.37 | 5.87E-05 | 9.55E-03 |
| Sodium ion transport | 183 | 6 | 6.25 | 4.74E-04 | 4.48E-02 |

Continued on next page

Table 5: GO enrichment analysis results for CD deep ulcer module A (117-gene module). (Continued)

| | | | | | |
|---|---|---|---|---|---|
| Regulation of lymphocyte mediated immunity | 185 | 6 | 6.18 | 5.01E-04 | 4.65E-02 |
| Regulation of T cell proliferation | 186 | 6 | 6.15 | 5.15E-04 | 4.70E-02 |
| Cytokine-mediated signalling pathway | 372 | 11 | 5.64 | 5.23E-06 | 1.38E-03 |
| Positive regulation of protein kinase activity | 365 | 9 | 4.7 | 1.51E-04 | 1.99E-02 |
| Innate immune response | 752 | 16 | 4.06 | 2.33E-06 | 7.57E-04 |
| Negative regulation of cell communication | 1360 | 18 | 2.52 | 2.66E-04 | 3.05E-02 |
| Negative regulation of signalling | 1361 | 18 | 2.52 | 2.68E-04 | 2.97E-02 |

Table 6: GO enrichment analysis results for CD deep ulcer module B (63-gene module).

| **GO biological process** | **Homo sapiens (REF)** | **Gene module** | **Fold Enrich.** | **Raw P value** | **FDR** |
|---|---|---|---|---|---|
| Toll-like receptor TLR6:TLR2 signalling pathway | 2 | 2 | >100 | 4.32E-05 | 7.17E-03 |
| Detection of diacyl bacterial lipopeptide | 2 | 2 | >100 | 4.32E-05 | 7.10E-03 |
| Cellular response to diacyl bacterial lipopeptide | 4 | 2 | >100 | 1.08E-04 | 1.45E-02 |
| Negative regulation of nucleotide-binding oligomerization domain containing 2 signalling pathway | 4 | 2 | >100 | 1.08E-04 | 1.42E-02 |
| Regulation of cellular response to macrophage colony-stimulating factor stimulus | 6 | 2 | >100 | 2.00E-04 | 2.28E-02 |
| Regulation of response to macrophage colony-stimulating factor | 6 | 2 | >100 | 2.00E-04 | 2.26E-02 |
| Negative regulation of toll-like receptor 2 signalling pathway | 7 | 2 | >100 | 2.57E-04 | 2.71E-02 |
| Growth hormone receptor signalling pathway via JAK-STAT | 8 | 2 | 91.92 | 3.21E-04 | 3.11E-02 |
| Positive regulation of oxidative stress-induced neuron death | 9 | 2 | 81.7 | 3.91E-04 | 3.53E-02 |
| Vitamin A metabolic process | 9 | 2 | 81.7 | 3.91E-04 | 3.51E-02 |
| Regulation of chronic inflammatory response | 10 | 2 | 73.53 | 4.69E-04 | 4.06E-02 |

Continued on next page

Table 6: GO enrichment analysis results for CD deep ulcer module B (63-gene module). (Continued)

| | | | | | |
|---|---|---|---|---|---|
| Regulation of toll-like receptor 3 signalling pathway | 10 | 2 | 73.53 | 4.69E-04 | 4.04E-02 |
| Regulation of natural killer cell proliferation | 15 | 3 | 73.53 | 1.50E-05 | 2.92E-03 |
| Positive regulation of interleukin-18 production | 10 | 2 | 73.53 | 4.69E-04 | 4.02E-02 |
| Cellular response to UV-A | 11 | 2 | 66.85 | 5.53E-04 | 4.40E-02 |
| Positive regulation of hepatocyte proliferation | 11 | 2 | 66.85 | 5.53E-04 | 4.38E-02 |
| Regulation of vascular wound healing | 11 | 2 | 66.85 | 5.53E-04 | 4.34E-02 |
| Positive regulation of nitric-oxide synthase biosynthetic process | 18 | 3 | 61.28 | 2.43E-05 | 4.46E-03 |
| Microglial cell activation | 31 | 4 | 47.44 | 2.39E-06 | 7.60E-04 |
| Positive regulation of cytokine production involved in inflammatory response | 25 | 3 | 44.12 | 5.90E-05 | 8.94E-03 |
| Embryonic digestive tract development | 33 | 3 | 33.42 | 1.27E-04 | 1.62E-02 |
| Neutrophil chemotaxis | 81 | 7 | 31.77 | 3.93E-09 | 4.09E-06 |
| Regulation of neuroinflammatory response | 35 | 3 | 31.51 | 1.49E-04 | 1.82E-02 |
| Astrocyte development | 37 | 3 | 29.81 | 1.74E-04 | 2.04E-02 |
| Collagen catabolic process | 41 | 3 | 26.9 | 2.31E-04 | 2.53E-02 |
| Lipopolysaccharide-mediated signalling pathway | 42 | 3 | 26.26 | 2.47E-04 | 2.63E-02 |
| Positive regulation of nitric oxide biosynthetic process | 43 | 3 | 25.65 | 2.64E-04 | 2.71E-02 |
| Positive regulation of interleukin-1 beta production | 61 | 4 | 24.11 | 2.91E-05 | 5.21E-03 |
| Extracellular matrix disassembly | 47 | 3 | 23.47 | 3.38E-04 | 3.18E-02 |
| Positive regulation of receptor signalling pathway via JAK-STAT | 47 | 3 | 23.47 | 3.38E-04 | 3.16E-02 |
| Response to amyloid-beta | 48 | 3 | 22.98 | 3.59E-04 | 3.28E-02 |
| Chemokine-mediated signalling pathway | 83 | 5 | 22.15 | 4.04E-06 | 1.11E-03 |
| Negative regulation of interleukin-6 production | 50 | 3 | 22.06 | 4.02E-04 | 3.57E-02 |
| Regulation of interleukin-8 production | 86 | 5 | 21.38 | 4.76E-06 | 1.24E-03 |
| Positive regulation of type II interferon production | 78 | 4 | 18.85 | 7.26E-05 | 1.04E-02 |

Continued on next page

Table 6: GO enrichment analysis results for CD deep ulcer module B (63-gene module). (Continued)

| | | | | | |
|---|---|---|---|---|---|
| Regulation of JUN kinase activity | 60 | 3 | 18.38 | 6.69E-04 | 5.00E-02 |
| Positive regulation of tumour necrosis factor production | 101 | 5 | 18.2 | 1.01E-05 | 2.15E-03 |
| Antimicrobial humoral immune response mediated by antimicrobial peptide | 104 | 5 | 17.68 | 1.15E-05 | 2.37E-03 |
| Acute inflammatory response | 84 | 4 | 17.51 | 9.56E-05 | 1.32E-02 |
| Killing of cells of another organism | 94 | 4 | 15.65 | 1.45E-04 | 1.78E-02 |
| Positive regulation of inflammatory response | 145 | 6 | 15.21 | 3.33E-06 | 1.02E-03 |
| Positive regulation of interleukin-6 production | 99 | 4 | 14.85 | 1.76E-04 | 2.05E-02 |
| Regulation of phagocytosis | 103 | 4 | 14.28 | 2.04E-04 | 2.27E-02 |
| Positive regulation of NF-kappaB transcription factor activity | 160 | 6 | 13.79 | 5.75E-06 | 1.40E-03 |
| Positive regulation of MAP kinase activity | 117 | 4 | 12.57 | 3.26E-04 | 3.10E-02 |
| Response to type II interferon | 131 | 4 | 11.23 | 4.94E-04 | 4.12E-02 |
| Intracellular receptor signalling pathway | 166 | 5 | 11.07 | 9.98E-05 | 1.35E-02 |
| Positive regulation of apoptotic signalling pathway | 135 | 4 | 10.89 | 5.51E-04 | 4.41E-02 |
| Calcium-mediated signalling | 137 | 4 | 10.73 | 5.81E-04 | 4.49E-02 |
| Positive regulation of protein-containing complex assembly | 201 | 5 | 9.15 | 2.38E-04 | 2.54E-02 |
| Defence response to bacterium | 294 | 6 | 7.5 | 1.58E-04 | 1.91E-02 |
| Positive regulation of protein transport | 316 | 6 | 6.98 | 2.32E-04 | 2.51E-02 |
| Regulation of Wnt signalling pathway | 334 | 6 | 6.6 | 3.10E-04 | 3.06E-02 |
| Negative regulation of catabolic process | 341 | 6 | 6.47 | 3.45E-04 | 3.21E-02 |
| Response to virus | 367 | 6 | 6.01 | 5.06E-04 | 4.20E-02 |
| Positive regulation of leukocyte activation | 379 | 6 | 5.82 | 5.98E-04 | 4.60E-02 |
| Negative regulation of cell population proliferation | 708 | 8 | 4.15 | 6.56E-04 | 4.92E-02 |
| Response to organic cyclic compound | 871 | 9 | 3.8 | 5.57E-04 | 4.35E-02 |
| Regulation of locomotion | 1034 | 10 | 3.56 | 4.46E-04 | 3.91E-02 |

Table 7: GO enrichment analysis results for CD no ulcer module D (45-gene module).

| GO biological process | Homo sapiens (REF) | Gene module | Fold Enrich. | Raw P value | FDR |
|---|---|---|---|---|---|
| Neutrophil aggregation | 2 | 2 | >100 | 2.54E-05 | 8.09E-03 |
| Antibody-dependent cellular cytotoxicity | 4 | 2 | >100 | 6.33E-05 | 1.57E-02 |
| Sequestering of zinc ion | 4 | 2 | >100 | 6.33E-05 | 1.54E-02 |
| Positive regulation of antibacterial peptide production | 5 | 2 | >100 | 8.85E-05 | 1.95E-02 |
| Peptidyl-cysteine S-nitrosylation | 6 | 2 | >100 | 1.18E-04 | 2.27E-02 |
| Autocrine signalling | 7 | 2 | >100 | 1.51E-04 | 2.75E-02 |
| Positive regulation of nitric-oxide synthase biosynthetic process | 18 | 4 | >100 | 1.16E-07 | 1.14E-04 |
| Positive regulation of interleukin-17 production | 28 | 3 | 51.3 | 3.64E-05 | 1.03E-02 |
| Fc-gamma receptor signalling pathway | 29 | 3 | 49.53 | 4.01E-05 | 1.12E-02 |
| Collagen catabolic process | 41 | 3 | 35.04 | 1.05E-04 | 2.16E-02 |
| Extracellular matrix disassembly | 47 | 3 | 30.56 | 1.54E-04 | 2.74E-02 |
| Neutrophil chemotaxis | 81 | 5 | 29.56 | 9.51E-07 | 6.45E-04 |
| Defence response to fungus | 58 | 3 | 24.77 | 2.79E-04 | 4.49E-02 |
| Positive regulation of inflammatory response | 145 | 6 | 19.81 | 6.78E-07 | 4.81E-04 |
| Positive regulation of tumour necrosis factor production | 101 | 4 | 18.96 | 6.71E-05 | 1.56E-02 |
| Antimicrobial humoral immune response mediated by antimicrobial peptide | 104 | 4 | 18.42 | 7.49E-05 | 1.72E-02 |
| Positive regulation of peptidyl-tyrosine phosphorylation | 181 | 5 | 13.23 | 4.11E-05 | 1.13E-02 |
| Cellular response to lipopolysaccharide | 199 | 5 | 12.03 | 6.38E-05 | 1.53E-02 |
| Defence response to bacterium | 294 | 7 | 11.4 | 2.70E-06 | 1.31E-03 |
| Cytokine-mediated signalling pathway | 372 | 7 | 9.01 | 1.22E-05 | 4.22E-03 |
| Positive regulation of DNA-binding transcription factor activity | 272 | 5 | 8.8 | 2.67E-04 | 4.34E-02 |
| Innate immune response | 752 | 12 | 7.64 | 3.25E-08 | 4.23E-05 |
| Positive regulation of programmed cell death | 527 | 7 | 6.36 | 1.07E-04 | 2.15E-02 |

[19] Umberto Michelucci. *An Introduction to Autoencoders*. 2022. arXiv: 2201.03898 [cs.LG].

[38] Tim Miller. *Explanation in Artificial Intelligence: Insights from the Social Sciences*. 2018. arXiv: 1706.07269 [cs.AI].

[39] Tim Miller. "Contrastive explanation: a structural-model approach". In: *The Knowledge Engineering Review* 36 (2021), e14. DOI: 10.1017/S0269888921000102.

[40] Soumya Banerjee, Pietro Lio, Peter B. Jones, et al. "A class-contrastive human-interpretable machine learning approach to predict mortality in severe mental illness". In: *npj Schizophrenia* 7 (1 Dec. 2021), p. 60. ISSN: 2334-265X. DOI: 10.1038/s41537-021-00191-y.

[41] Scott Lundberg and Su-In Lee. *A Unified Approach to Interpreting Model Predictions*. 2017. arXiv: 1705.07874 [cs.AI].

[72] Marco Tulio Ribeiro, Sameer Singh, and Carlos Guestrin. *"Why Should I Trust You?": Explaining the Predictions of Any Classifier*. 2016. arXiv: 1602.04938 [cs.LG].
